# Wastewater metagenomic sequencing quantitatively tracks transmission dynamics for human pathogenic viruses

**DOI:** 10.64898/2026.11.13.26368067

**Authors:** Elana M. G. Chan, Lennart Justen, Alessandro Zulli, Michael R. McLaren, Rebecca Y. Linfield, Leon S. Moskatel, Charles Whittaker

## Abstract

Wastewater metagenomic sequencing theoretically characterizes all nucleic acids in a sample, but whether sequencing reads reflect transmission dynamics of circulating viruses in the contributing community is unknown. We evaluated if wastewater metagenomic sequencing data can be used to reconstruct transmission dynamics of endemic, routinely circulating viruses. Using deep, untargeted wastewater metagenomic sequencing data from four locations in the United States, we generated quantitative estimates of transmission dynamics (time-varying growth rate and effective reproduction number) within a Bayesian framework for widely monitored viruses. Across locations and viruses, estimates were statistically concordant with those derived from clinical surveillance and polymerase chain reaction (PCR)-based wastewater monitoring data. Furthermore, we estimated growth rate from wastewater metagenomic sequencing data for numerous viruses not otherwise widely monitored, demonstrating the data expands the targets that can feasibly be monitored in parallel. Wastewater metagenomic sequencing can support both novel pathogen detection and routine surveillance of endemic pathogens.

## Introduction

The COVID-19 pandemic significantly expanded the use of wastewater monitoring for infectious disease surveillance. Before the pandemic, wastewater monitoring was primarily used for binary pathogen detection and notably to support polio eradication efforts.^1^ During the pandemic, studies demonstrated that wastewater concentrations of SARS-CoV-2 RNA reflect COVID-19 incidence,^2,3^ bringing mainstream attention to its use for quantitatively tracking disease transmission dynamics. Wastewater monitoring has since expanded to a myriad of pathogens—including other respiratory viruses,^4^ enteric viruses,^5^ fungi,^6^ bacteria,^7,8^ and arboviruses^9,10^—along with other purposes, such as tracking variants,^11,12^ identifying trends,^13^ evaluating policies and nonpharmaceutical interventions,^14,15^ and inferring seasonal characteristics and drivers.^16–20^

Presently, most wastewater monitoring efforts use quantitative polymerase chain reaction (PCR)-based methods to detect prespecified, known viruses.^21^ These “targeted” methods have shown significant utility for detecting and quantitatively tracking known pathogens. However, because assay design requires prior knowledge of the target genome sequence, targeted approaches are neither suited to detect novel or rapidly evolving pathogens nor feasibly monitor the full diversity of viruses in wastewater. By contrast, metagenomic sequencing characterizes nucleic acids across a sample without needing to prespecify targets. Shotgun metagenomic sequencing methods, referred to here as “untargeted” wastewater monitoring, involves sequencing all genomic material present in a wastewater sample. Nucleic acids from human pathogenic viruses comprise a small fraction of this material, however, so sensitive and consistent detection requires substantial sequencing depth or high total sequencing reads per sample.^22^ Nonetheless, declining sequencing costs have increased feasibility for scaling wastewater metagenomic sequencing with most attention focusing on its capability to detect novel or unexpected pathogens.

In addition to its ability to detect novel or unexpected pathogens, wastewater metagenomic sequencing generates billions of reads of expected, endemic targets, meaning the data are also positioned to provide broader public health value. Virus-specific normalized read abundance from untargeted wastewater monitoring data have been shown to temporally correlate with targeted wastewater monitoring and clinical surveillance data for human pathogenic viruses,^23,24^ suggesting that wastewater metagenomic sequencing may be used to infer quantitative transmission dynamics of both widely-surveilled (e.g., SARS-CoV-2) and under-surveilled (e.g., rhinovirus) pathogens. If so, wastewater metagenomic sequencing could serve multiple public health functions, including early detection of emerging, unexpected, or hitherto unseen threats and routine, quantitative monitoring of multiple endemic pathogens in parallel.

Whether virus-specific metagenomic read abundance can reliably reconstruct underlying transmission dynamics remains unknown. Two quantitative statistics widely used to monitor transmission dynamics are the instantaneous epidemic growth rate (*r_t_*) and effective reproduction number (*R_t_*).^25,26^ Both *r_t_* and *R_t_*have been estimated from wastewater monitoring data, although only using data collected via targeted approaches.^27,28^ To our knowledge, no studies have evaluated whether untargeted wastewater monitoring data may be used to reliably infer transmission dynamics (as represented by *r_t_* and *R_t_*). In this study, we aimed to assess the use of untargeted wastewater monitoring data for quantitatively tracking pathogen transmission dynamics. Using data from four distinct sites in the United States, we estimated *r_t_* and *R_t_* using untargeted wastewater monitoring, targeted wastewater monitoring, and clinical surveillance data for widely-monitored viruses. We then evaluated concordance among the time-varying estimates of pathogen transmission derived from each data source. Secondarily, we estimated *r_t_* for viruses only with available untargeted wastewater monitoring data to infer the transmission dynamics of less-monitored viruses. We found that wastewater metagenomic sequencing tracks transmission dynamics concordant with other data sources while expanding the viral targets we can reliably monitor.

## Results

We obtained clinical surveillance, targeted wastewater monitoring, and untargeted wastewater monitoring data for various viruses from four sites in the United States (**Fig. 1**): Boise, Idaho (November 2024–March 2026); Boston, Massachusetts (September 2024–March 2026); Riverside, California (December 2024–March 2026); and Ottumwa, Iowa (February 2025–March 2026). Refer to the Methods for a complete overview of data sources. We obtained weekly totals of disease diagnoses by International Classification of Diseases, 10th revision (ICD-10) code (**Table S1–2**) from Epic Cosmos within the county or state of sites.^29^ For influenza A and B virus (IAV/IBV), we instead obtained weekly totals of positive lab results. Targeted and untargeted wastewater monitoring data were obtained from wastewater treatment plants as described by Boehm et al.^21^ and Justen et al.^23,24^, respectively. We used viral nucleic acid concentrations normalized by pepper mild mottle virus (PMMoV) RNA concentrations collected three times per week for targeted wastewater monitoring data; these data were only available through 15 September 2025. For untargeted wastewater monitoring data, we used weekly virus clade counts normalized by PMMoV clade counts that were scaled to account for sequencing depth. Among viruses with multiple data sources, surveillance time series were overall positively correlated across locations during weeks with concurrent data (**Table S3**). Therefore, we proceeded to estimate *r_t_* and *R_t_* for viruses using each data source and assess concordance among the derived estimates. We derived *r_t_* by smoothing time series data using a generalized additive model (GAM), implemented in a Bayesian framework using the mgcv R package,^30^ and then calculating the logarithmic derivative of smoothed data.^26^ To derive *R_t_*, we used the renewal equation model in a Bayesian framework^31^ as described by the R package EpiNow2.^32^ Refer to the Methods for model specifications, but both approaches derive daily estimates.

**Fig. 1.**
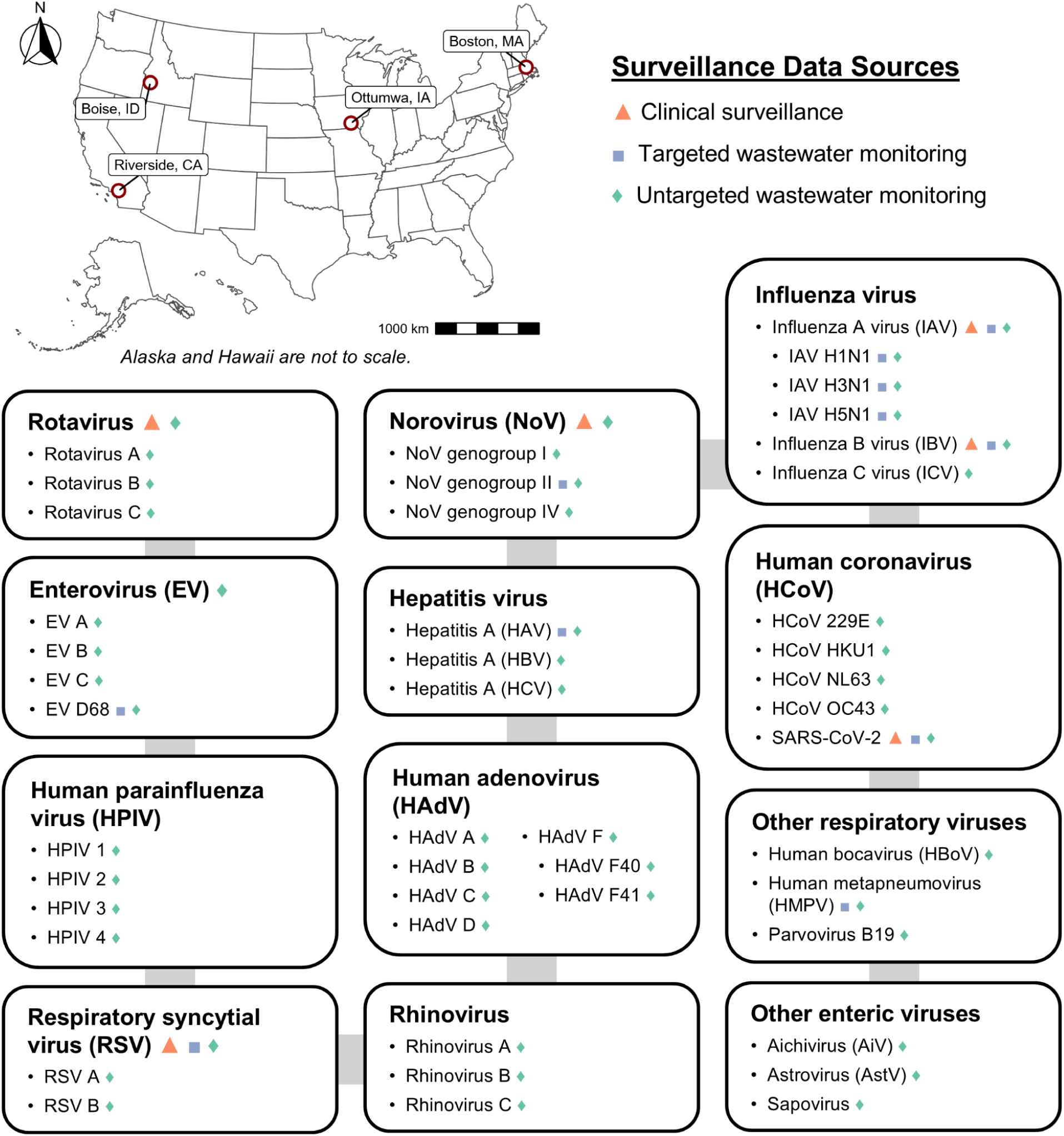
Map of study sites and selected human pathogenic viruses. Surveillance data were obtained for viruses from four locations in the United States. Symbols and colors denote the surveillance data source(s) obtained for viruses at each site (orange triangles = Epic Cosmos clinical surveillance, purple squares = targeted wastewater monitoring, green diamonds = untargeted wastewater monitoring). Note that targeted wastewater monitoring data are published in Boehm et al.^21^ for rotavirus, HAdV F, and parvovirus B19. However, the duration of routine targeted wastewater monitoring was less than six months during the period when untargeted wastewater monitoring was ongoing from the study sites, so we did not include targeted wastewater monitoring data for these viruses in our study.

### Transmission dynamics derived from wastewater metagenomic sequencing are concordant with other data sources

We estimated *R_t_* for SARS-CoV-2, IAV, IBV, respiratory syncytial virus (RSV), norovirus (NoV), and rotavirus. The renewal equation links observed surveillance signals to latent infections in the population via a generative model of transmission and infection-to-reporting and infection-to-shedding delays for clinical and wastewater data sources, respectively. These viruses had surveillance data from multiple sources, and we obtained sufficient data to inform prior model assumptions—particularly regarding fecal shedding dynamics (**Table S4**). In contrast, calculating *r_t_* is not based on a model of disease transmission, so we estimated *r_t_* for all viruses. For each wastewater data source, we only derived *r_t_* and *R_t_* if at least 75% of samples had positive detections.

An example of SARS-CoV-2 surveillance data and derived *R_t_*estimates are shown in **Fig. 2** for each location. Durations of surveillance data varied across sites and data sources, but all had ongoing surveillance during summer 2025 with SARS-CoV-2 transmission increasing during this period (*R_t_* > 1). Averaging across data sources, peak *R_t_* occurred between 29 June (Ottumwa) and 6 Aug (Boston). Among sites, the peak *R_t_* derived from untargeted wastewater monitoring data overall occurred within 2 weeks of that derived from other data. Between untargeted wastewater monitoring and clinical data, peak *R_t_* estimates occurred within 12 days for Boise, Boston, and Riverside and 24 days apart for Ottumwa. Between untargeted and targeted wastewater monitoring data, peak *R_t_* estimates occurred within 14 days for Boise, Ottumwa, and Riverside and 63 days apart for Boston. The magnitude of peak *R_t_*values derived from untargeted wastewater monitoring data differed, on average, 0.04 (range: 0.01–0.12) from those derived from clinical data and 0.04 (range: 0.02–0.06) from those derived from targeted wastewater monitoring data. Surveillance data with GAM smoothing alongside *r_t_*estimates are shown in **Fig. S1**. Derived *r_t_* estimates similarly indicated increasing SARS-CoV-2 transmission during summer 2025 across data sources (*r_t_* > 0), although average peak *r_t_* dates (range: 13 July–12 Aug) occurred later than average peak *R_t_* dates.

**Fig. 2.**
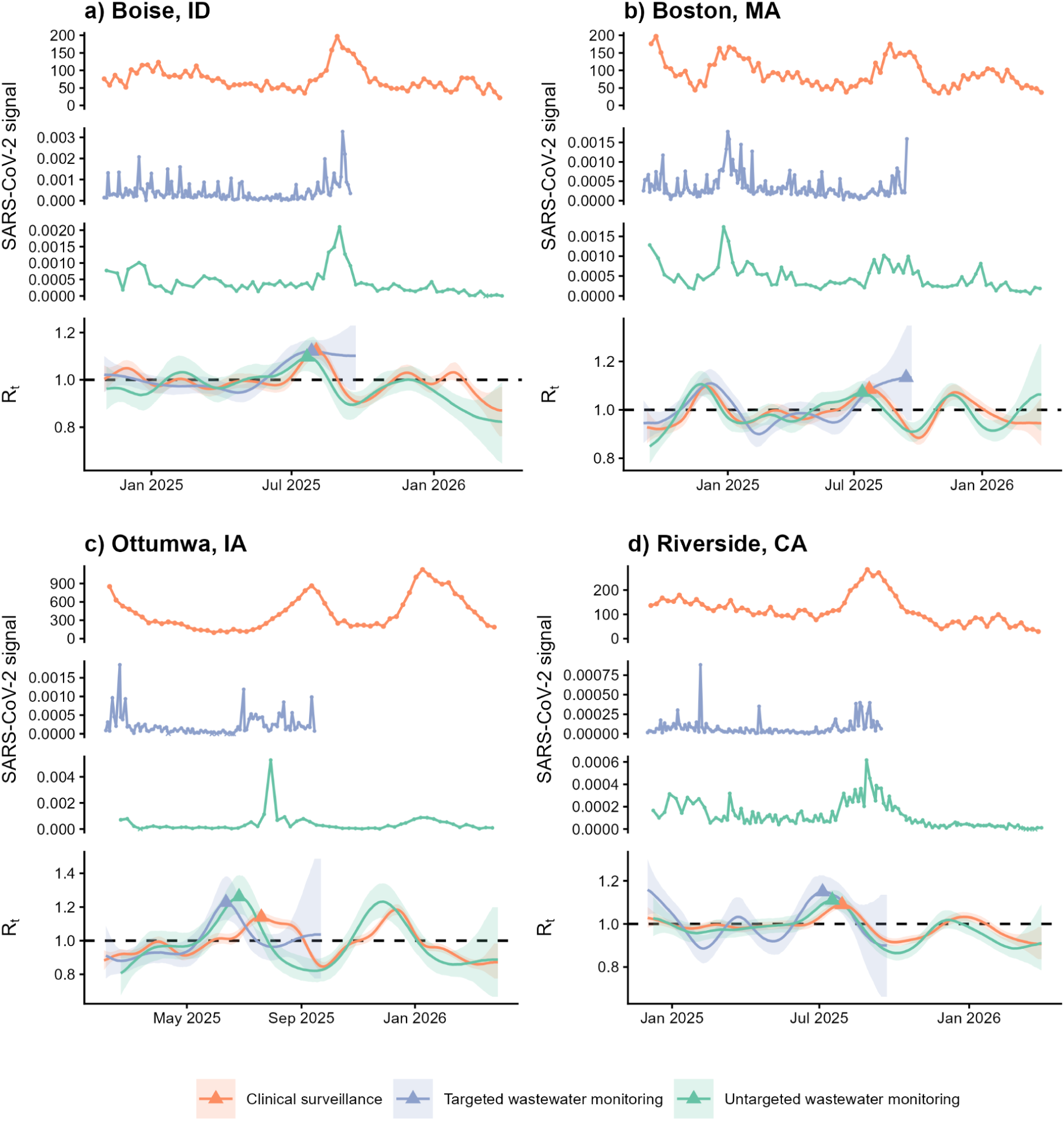
SARS-CoV-2 surveillance data and *R_t_* estimates for all locations. Time series data from each data source alongside *R_t_* estimates are shown for (**a**) Boise, Idaho; (**b**) Boston, Massachusetts; (**c**) Ottumwa, Iowa; and (**d**) Riverside, California. “Signal” indicates counts of COVID-19 diagnoses from Epic Cosmos for clinical data (orange), PMMoV-normalized concentrations of SARS-CoV-2 RNA for targeted wastewater data (purple), and PMMoV-normalized SARS-CoV-2 clade counts scaled to account for sequencing depth for untargeted wastewater data (green). For wastewater monitoring data sources, x’s denote non-detect samples; outlier measurements are not shown for visualization. On the *R_t_* panels, lines represent median estimates with 90% CrIs shaded; upward triangles denote the peak *R_t_* estimate derived from each data source during summer 2025. Abbreviations: CrI = credible interval, PMMoV = pepper mild mottle virus.

Derived *R_t_* estimates for SARS-CoV-2, alongside the other viruses for which *R_t_* was derived, are shown in **Fig. 3a** for Boston. For NoV, we used clinical data irrespective of genogroup and wastewater monitoring data specific to genogroup II as only genogroup II had data from both wastewater sources and is the predominant genogroup.^33^ All data sources had ongoing surveillance during the 2024–25 fall–spring season, and increasing transmission occurred for each virus (*R_t_* > 1). In general, the peak *R_t_* derived from untargeted wastewater monitoring data mostly occurred within 2 weeks of that derived from other data for each virus (**Fig. 3b**). Excluding IBV, peak *R_t_* estimates derived from untargeted wastewater monitoring occurred, on average, 8 days (range: 0–17 days) apart from those derived from clinical data and 10 days (range: 5–14 days) apart from those derived from targeted wastewater monitoring data. For IBV, the peak *R_t_* derived from untargeted wastewater monitoring data occurred over a month later than the peak derived from both other data. **Fig. 3c** displays the magnitude of peak *R_t_* estimates by data source and virus. For IBV and RSV, the peak *R_t_* magnitude derived from untargeted wastewater monitoring data was over 0.10 below the peak derived from other data. For other viruses, peak *R_t_* magnitudes derived from untargeted wastewater monitoring data only differed, on average, 0.04 (range: 0.01–0.08) from those derived from clinical data and 0.02 (range: <0.01–0.06) from those derived from targeted wastewater monitoring data. Refer to **Figs. S2–12** for surveillance data alongside *R_t_* and *r_t_* estimates for these viruses and other locations. As with SARS-CoV-2, derived *r_t_* estimates mostly followed similar dynamics as derived *R_t_* dynamics, although peak *r_t_* dates generally occurred later than peak *R_t_* dates.

**Fig. 3.**
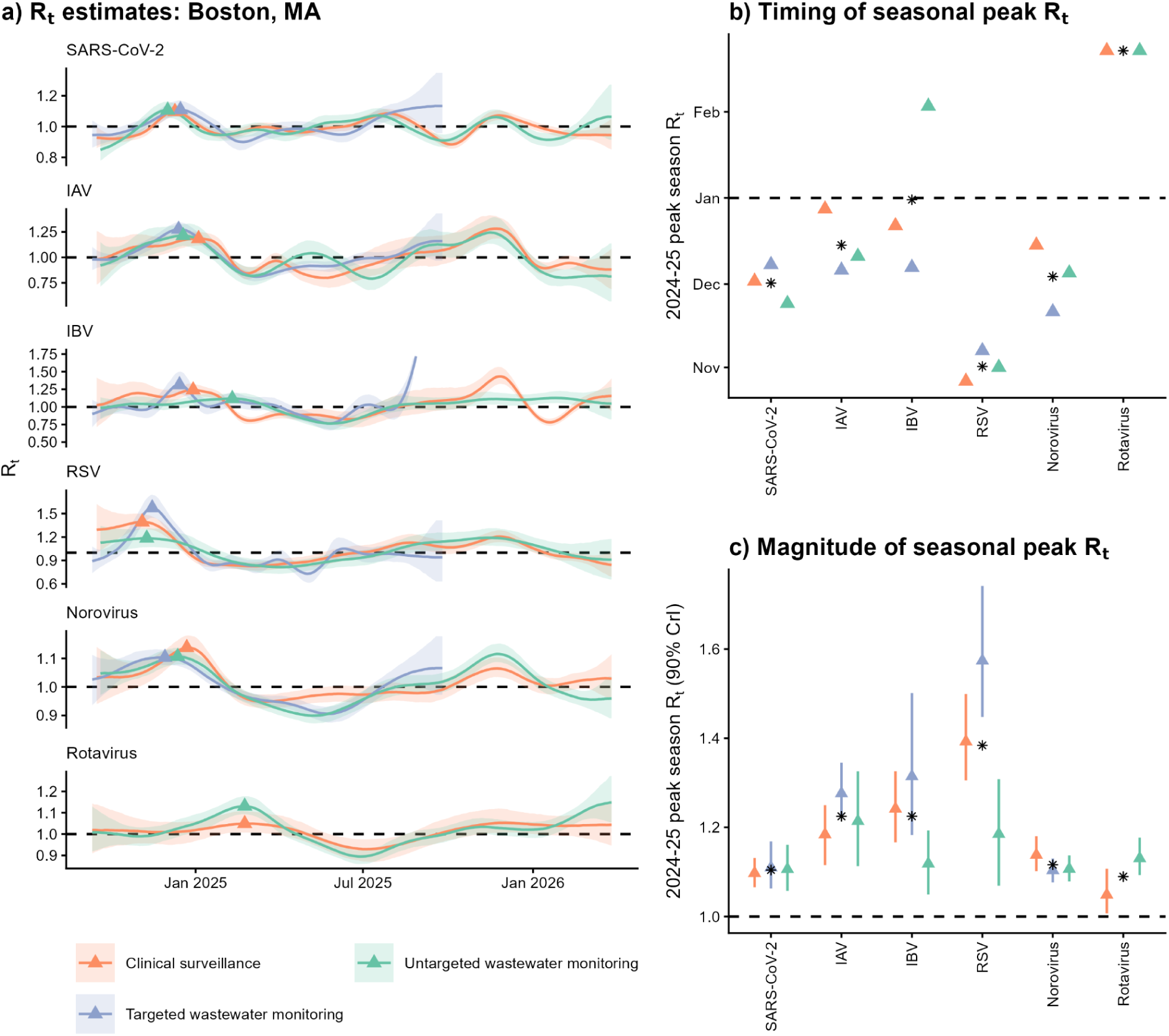
SARS-CoV-2, IAV, IBV, RSV, norovirus, and rotavirus *R_t_* estimates for Boston, Massachusetts. (**a**) *R_t_* estimates are shown for each virus as derived from each data source (orange = Epic Cosmos clinical surveillance, purple = targeted wastewater monitoring, green = untargeted wastewater monitoring). Lines represent median estimates with 90% CrIs shaded; upward triangles denote the peak *R_t_*estimate observed from each data source during the 2024–25 fall–spring season. The y-axis for IBV was adjusted in panel a such that extreme estimates are not displayed for visualization purposes only; refer to **Fig. S5** for this figure without adjustment. For norovirus, clinical data include all genogroups whereas wastewater monitoring data sources are specific to genogroup II. (**b**) Timing of peak *R_t_* estimates during the 2024–25 season for each virus and data source; black stars denote average dates across data sources. (**c**) Magnitude of peak *R_t_* estimates during the 2024–25 season for each virus and data source; error bars denote 90% CrIs and black stars denote average values across data sources. Abbreviations: CrI = credible interval, IAV = influenza A virus, IBV = influenza B virus, RSV = respiratory syncytial virus.

To statistically assess whether derived estimates were concordant among data sources across the entire period with concurrent surveillance, we calculated three complementary metrics. Mean absolute error (MAE) quantifies numerical agreement, agreement on binary (AOB) assesses agreement on epidemic direction, and Kendall’s tau correlation evaluates temporal association (**Fig. 4a**). We calculated each metric between all pairs of posterior draws between data source *a* and *b*, generating observed posterior distributions. We further used Bayesian permutation analysis to generate null posterior distributions which allowed us to evaluate whether observed concordance between two data sources was statistically significant. Refer to the Methods for complete details, but permutation *p* values < 0.05 suggested significant concordance.

**Fig. 4.**
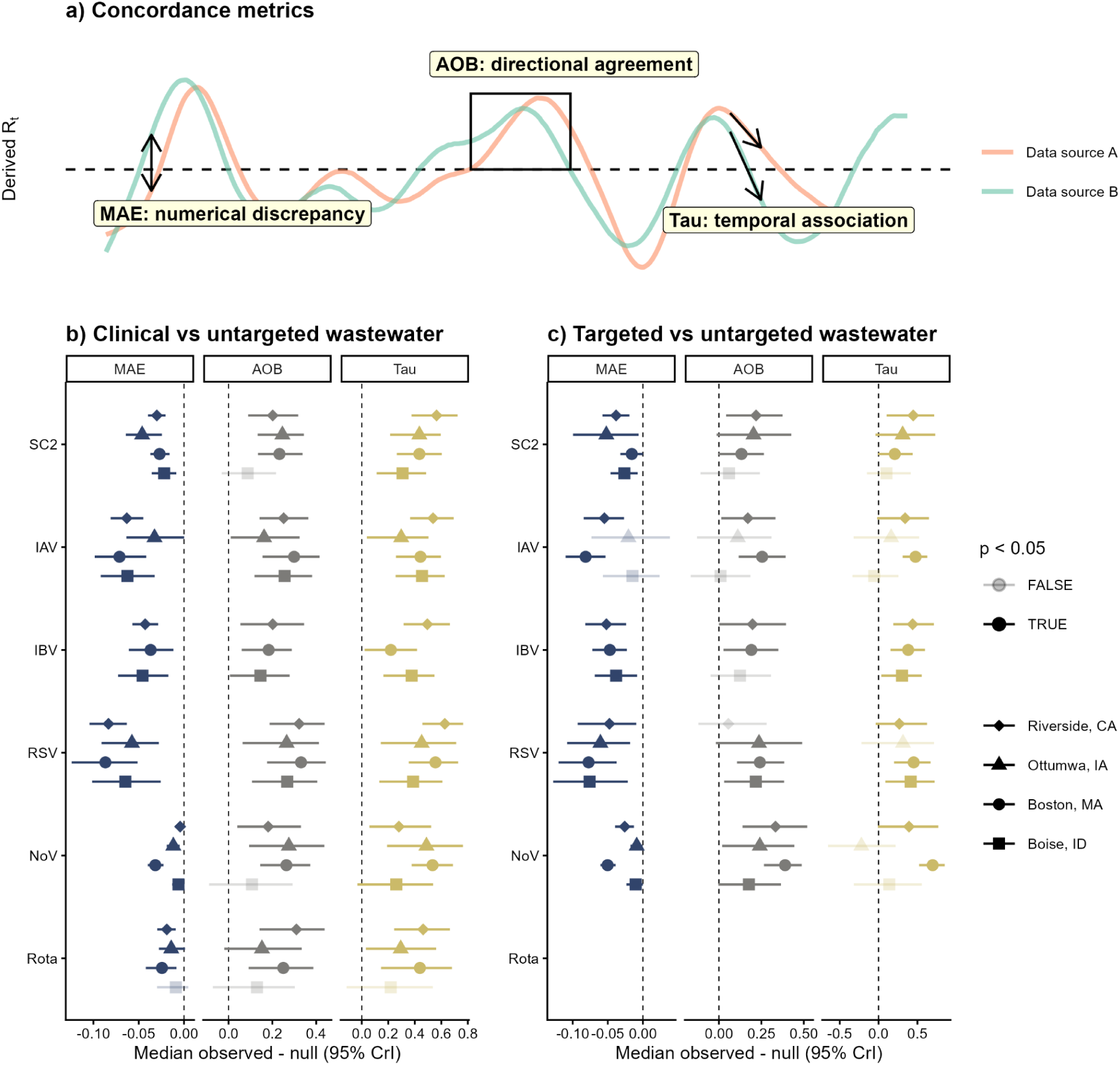
Concordance between *R_t_*estimates derived from untargeted wastewater monitoring versus other data sources. (**a**) Schematic depicting the three complementary measures of concordance. Difference between the observed and null posterior distributions of *R_t_* estimates derived from (**b**) Epic Cosmos clinical surveillance versus untargeted wastewater monitoring and (**c**) targeted versus untargeted wastewater monitoring data. Symbols denote median differences for unique locations with error bars representing 95% CrIs; darker hues indicate significantly similar derived *R_t_* estimates. For NoV, clinical data include all genogroups whereas wastewater monitoring data sources are specific to genogroup II. For Ottumwa, *R_t_* estimates were not derived from untargeted wastewater monitoring data for IBV as >75% of samples did not have a positive detection. Here, *p* values represent Bayesian permutation probability values, with *p* < 0.05 indicating significant concordance. Abbreviations: AOB = agreement on binary, CrI = credible interval, IAV = influenza A virus, IBV = influenza B virus, MAE = mean absolute error, NoV = norovirus, Rota = rotavirus, RSV = respiratory syncytial virus, SC2 = SARS-CoV-2.

Overall, *R_t_* estimates derived from untargeted wastewater monitoring data were significantly concordant across all metrics when compared with those derived from the other data sources (**Table S5**). We conducted a total of 69 tests comparing *R_t_* estimates derived from untargeted wastewater monitoring and clinical data; 64 tests indicated significant concordance (**Fig. 4b**). *R_t_* estimates derived from these data sources were significantly concordant based on all metrics for all viruses and sites with the exception of Boise. For Boise, derived *R_t_* estimates were not significantly concordant based on AOB for SARS-CoV-2 and NoV and based on all metrics for rotavirus. We conducted a total of 57 tests comparing *R_t_* estimates derived from untargeted and targeted wastewater monitoring data; 44 tests indicated significant concordance (**Fig. 4c**). *R_t_* estimates derived from these data sources were only significantly concordant for Boston based on all metrics for all viruses. For Riverside, derived *R_t_*estimates were not significantly concordant based on AOB for RSV. For Ottumwa and Boise, derived *R_t_* estimates were not significantly concordant based on all metrics for IBV and 1–2 metrics for other viruses.

We also assessed concordance among derived *r_t_* estimates using the same approach (**Fig. S13**, **Table S6**). We additionally included hepatitis A virus (HAV), IAV H1N1, and human metapneumovirus (HMPV) in the concordance assessment as these viruses had sufficient time series data for us to derive *r_t_*from more than one data source (**Figs. S14–16**). Overall, *r_t_*estimates derived from untargeted wastewater monitoring data were similarly significantly concordant across all metrics when compared with those derived from other data sources. We conducted a total of 69 tests comparing *r_t_* estimates derived from untargeted wastewater monitoring and clinical data; 65 tests indicated significant concordance. We conducted a total of 66 tests comparing *r_t_* estimates derived from untargeted and targeted wastewater monitoring data; 50 tests indicated significant concordance.

Moreover, we derived *R_t_* estimates for SARS-CoV-2, IAV, and RSV using alternative fecal shedding distributions to evaluate the sensitivity of the *R_t_* modeling framework to fecal shedding dynamics (**Table S7**). We then reassessed concordance between *R_t_* estimates derived from untargeted wastewater monitoring and other data sources (**Table S5**). Overall, concordance remained significant when using alternative shedding distributions. For example, **Fig. S17** displays concordance results for Boston—the location with the longest duration of concurrent surveillance across data sources—using each fecal shedding distribution. *R_t_* estimates derived from untargeted wastewater monitoring data remained significantly concordant with those derived from other data sources based on all three metrics for all shedding distributions considered except for one SARS-CoV-2 shedding distribution.

### Longitudinal wastewater metagenomic sequencing offers insight into the transmission dynamics of less-monitored pathogens

For most viruses, surveillance data were only available from untargeted wastewater monitoring data. Given transmission dynamics derived from untargeted wastewater monitoring data were overall concordant with those derived from clinical and targeted wastewater monitoring data for the viruses with multiple surveillance sources, we derived *r_t_* estimates from untargeted wastewater monitoring data alone for remaining viruses. Refer to **Figs. S18–29** for untargeted wastewater monitoring data and derived *r_t_*estimates for each virus and site, but a summary for Boston is shown in **Fig. 5**.

**Fig. 5.**
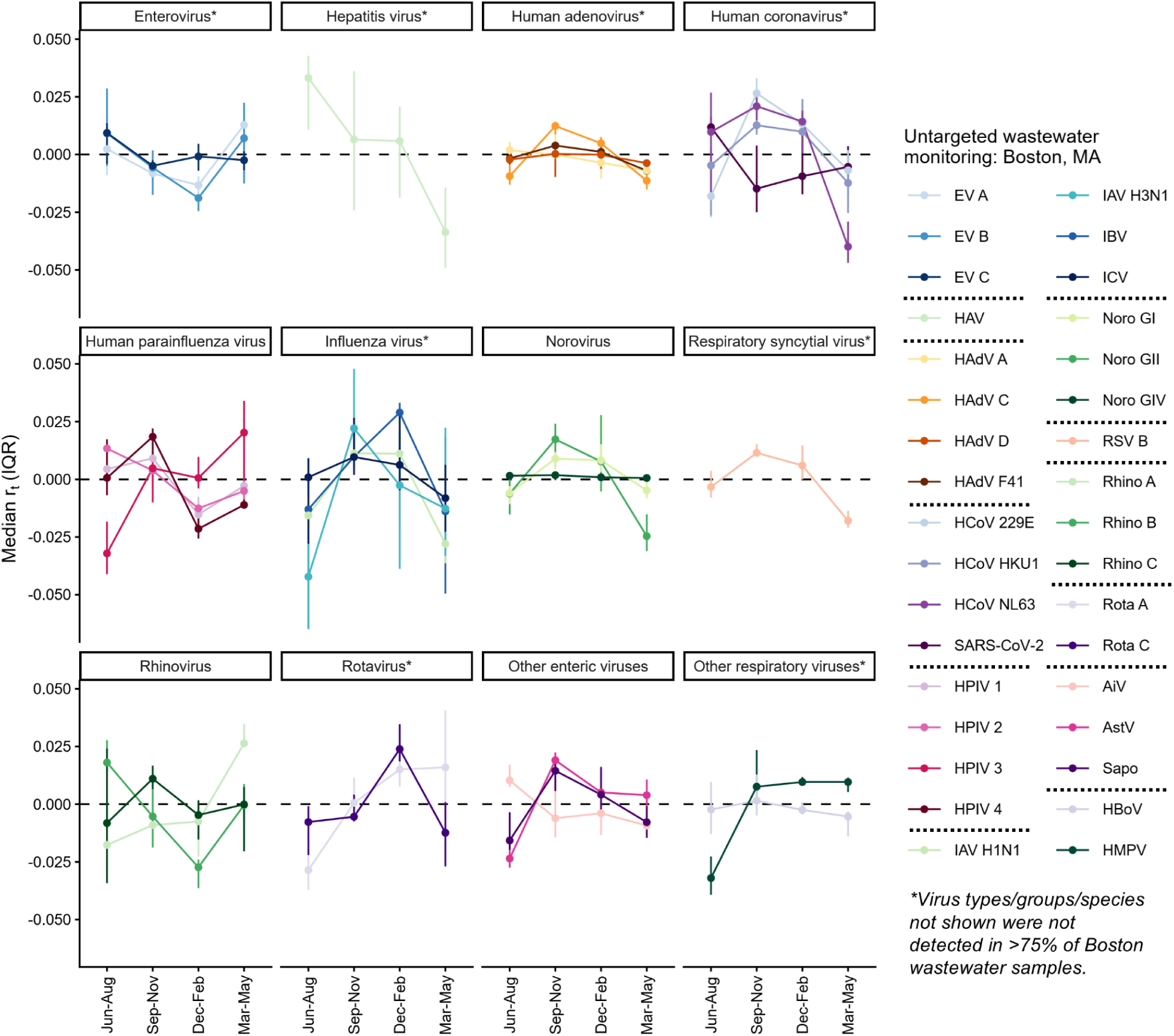
*r_t_* estimates for all viruses derived from untargeted wastewater monitoring data for Boston, Massachusetts. Median *r_t_*estimates by season (summer = Jun–Aug, fall = Sep–Nov, winter = Dec–Feb, spring = Mar–May) are shown for all viruses detected in at least 75% of untargeted wastewater samples; error bars represent the IQR. Abbreviations: AiV = aichivirus, AstV = astrovirus, EV = enterovirus, HAdV = human adenovirus, HAV = hepatitis A virus, HBoV = human bocavirus, HCoV = human coronavirus, HMPV = human metapneumovirus, HPIV = human parainfluenza virus, IAV/IBV/ICV = influenza A/B/C virus, IQR = interquartile range, Noro = norovirus, Rota = rotavirus, Rhino = rhinovirus, RSV = respiratory syncytial virus, Sapo = sapovirus.

In **Fig. 5**, we temporally aggregated *r_t_* estimates by season (Jun–Aug: summer, Sept–Nov: fall, Dec–Feb: winter, Mar–May: spring) to summarize relative differences in *r_t_* estimates among viruses. Not all viruses may be intrinsically seasonal, or the seasonal cutoffs used may obfuscate peaks that occur during seasonal transitions; refer to **Figs. S18–29** for disaggregated *r_t_* time series. Nonetheless, **Fig. 5** highlights broad differences in *r_t_* estimates across calendar time and viruses. For instance, enterovirus (EV) D68 was rarely detected in Boston wastewater, but EV A–C were detected with *r_t_*generally elevated in spring–summer. Only HAV was consistently detected among hepatitis viruses; HBV was rarely detected and HCV was never detected. Aside from HAdV B, all HAdV groups were detected. For HAdV F, HAdV F40 was rarely detected and HAdV F41 was commonly detected. Relative to other viruses, HAdV *r_t_* estimates did not vary much in magnitude and exhibited minimal seasonal differences. HCoV OC43 was rarely detected but all other HCoVs were detected with derived *r_t_* estimates generally highest in fall. In contrast, *r_t_* estimates for SARS-CoV-2 were generally lowest in fall and highest in summer. HPIV 1, 2, and 4 all exhibited similar seasonal patterns with *r_t_* estimates typically > 0 in summer–fall and < 0 in winter–spring. HPIV 3 *r_t_* estimates were highest in spring and lowest in summer. For IAV, IAV H1N1 and IAV H3N1 displayed similar patterns, with *r_t_* highest in fall; IAV H5N1 was rarely detected. Generally, IBV *r_t_* estimates were highest in winter and ICV *r_t_*estimates were highest in fall—although the magnitude of ICV *r_t_*estimates fluctuated the least, comparatively. All NoV genogroups were detected. Derived *r_t_* estimates for NoV GIV displayed negligible differences across seasons whereas *r_t_* estimates for NoV GI and NoV GII were typically elevated in fall with magnitudes being higher for NoV GII compared to NoV GI. During the study period, RSV B was more common than RSV A; RSV B *r_t_* estimates were > 0 in fall–winter. Rhinovirus A–C were all present in wastewater with each species exhibiting unique patterns. Derived *r_t_* estimates were highest in spring for rhinovirus A, summer for rhinovirus B, and fall for rhinovirus C. Rotavirus B was never detected, but both rotavirus A and C were detected with the highest *r_t_* estimates occurring in winter. Among the other enteric viruses, astrovirus and sapovirus *r_t_* estimates exhibited similar temporal patterns, with estimates most elevated in fall. Aichivirus *r_t_* estimates were instead highest in summer. Lastly, parvovirus B19 was rarely detected but the other two respiratory viruses, human bocavirus and HMPV, were more often detected. HMPV *r_t_*estimates were generally < 0 in summer but elevated in the remaining seasons. Comparatively, bocavirus *r_t_* estimates did not fluctuate much across seasons.

## Discussion

Wastewater metagenomic sequencing—or untargeted wastewater monitoring—is particularly well-suited to detect rare, albeit potentially consequential, novel and reemerging threats. Yet the data simultaneously offer routine public health value as most wastewater metagenomic reads belonging to viruses are of circulating, endemic pathogens. We demonstrated that untargeted wastewater monitoring may be used to quantitatively track transmission dynamics of routinely detected viruses while expanding the range of targets that may be reliably monitored. Estimates of growth rate (*r_t_*) and effective reproduction number (*R_t_*) derived from untargeted wastewater monitoring data were significantly concordant with those derived from clinical data and targeted, PCR-based wastewater monitoring data. Moreover, untargeted wastewater monitoring data covered more viral targets than other data sources, allowing transmission dynamics to be inferred for a wider number of targets.

Across distinct locations and viruses, we provided evidence that untargeted wastewater monitoring can recapitulate *R_t_* and *r_t_*as derived from clinical and targeted wastewater monitoring. The timing and magnitude of peak *R_t_* and *r_t_* estimates derived from untargeted wastewater monitoring data were overall relatively similar as when derived from other data sources during periods of increasing disease transmission. In addition, the relative timing of observed peak *R_t_*values derived from untargeted wastewater monitoring data during the 2024–25 season aligned with established temporal patterns based on historic clinical and targeted wastewater monitoring data. RSV typically peaks first, followed by IAV then IBV which we observed.^19,34^ The seasonality of SARS-CoV-2 is not as established but in past years has had a wintertime peak coinciding with RSV and influenza;^16,19^ we observed a peak in SARS-CoV-2 *R_t_* between that of RSV and influenza. NoV activity is typically elevated in winter and rotavirus in winter–spring,^35–38^ and we observed a wintertime peak in *R_t_* for NoV before rotavirus.

Our Bayesian permutation analysis then provided statistical evidence that the observed similarities in *R_t_* and *r_t_*estimates derived from untargeted wastewater monitoring and other data sources were significant. Other studies have calculated metrics similar to MAE, AOB, and correlation to assess similarity between two derived time series, although only between point estimates.^27,39,40^ By using permutation within a Bayesian framework, we not only quantified the numerical discrepancy, agreement on epidemic direction, and temporal association between two time series but further provided statistical evidence that the observed metrics were significantly different than had there been no relationship between the two time series.

Nonetheless, there are shortcomings to our statistical approach which may explain why not all tests were significant. First, some clinical data were sparse at the county level so we used state-level data instead (**Table S2**). Lack of concordance between estimates from untargeted wastewater monitoring and clinical data may be attributed to geographic differences between the localized and statewide population. Second, the duration of targeted wastewater monitoring was shorter than other data sources. *R_t_* estimates towards the end of the time window of a data source are less reliable; therefore, the end date of targeted wastewater monitoring occurring before that of clinical surveillance and untargeted wastewater monitoring data may have contributed to estimates derived from targeted wastewater monitoring sometimes lacking concordance with other data sources. Third, the permutation testing we adopted to infer significance of similarity depends on temporal variation in the data. Permuting a time series with its own time points randomizes that variation and enables principled construction of a null distribution and comparison. By contrast, time series with relatively low degrees of temporal variation (e.g., those that fluctuate minimally around *R_t_* = 1) do not differ much once permuted. Consequently, achieving significant differentiation between the observed and null posterior distributions is challenging with time series that do not exhibit substantial temporal variation. Lack of a complete year of concurrent surveillance (aside from Boston) may explain why estimates derived from untargeted wastewater monitoring were not always significantly concordant with other data sources despite visually appearing so. Lastly, we convolved latent infections to expected case reports using incubation period and reporting delay distributions for clinical data and fecal shedding distributions for wastewater data sources. While these distributions theoretically account for the delays unique to each data source, meaning inferred peak *R_t_* estimates should coincide across data sources, information to deduce these distributions—especially reporting and shedding delay distributions—is sparse. Lack of reporting and shedding delay data for viruses may further explain why derived *R_t_* estimates were not always concordant across data sources.

Most viruses included in this study only had untargeted wastewater monitoring data available. For the viruses with multiple surveillance sources, estimates from untargeted wastewater monitoring data were overall concordant when compared to other data sources. Therefore, we derived *r_t_*from untargeted wastewater monitoring alone for remaining viruses. Overall, *r_t_* estimates from untargeted wastewater monitoring data aligned with known epidemiological features of these viruses according to historical surveillance (**Table 1**). Because many of these viruses are less prioritized compared to SARS-CoV-2, influenza, and RSV, future work should focus on better understanding the epidemiology of these less-studied viruses. Wastewater metagenomic sequencing offers a compelling means to do so as it does not rely on testing-seeking behaviors, clinical testing of specimens, or targeted assay design. For example, Smith et al.^41^ retrospectively elucidated seasonal dynamics for a range of targets detected via wastewater metagenomic sequencing data using linear mixed effects models. Future studies should build upon this work to develop methods for prospectively identifying levels and trends or seasonal onset, peak, offset, and duration. Such methods have already been developed for targeted wastewater monitoring data;^13,16–19^ extending this work to untargeted wastewater monitoring data would greatly expand its utility.

**Table 1.**
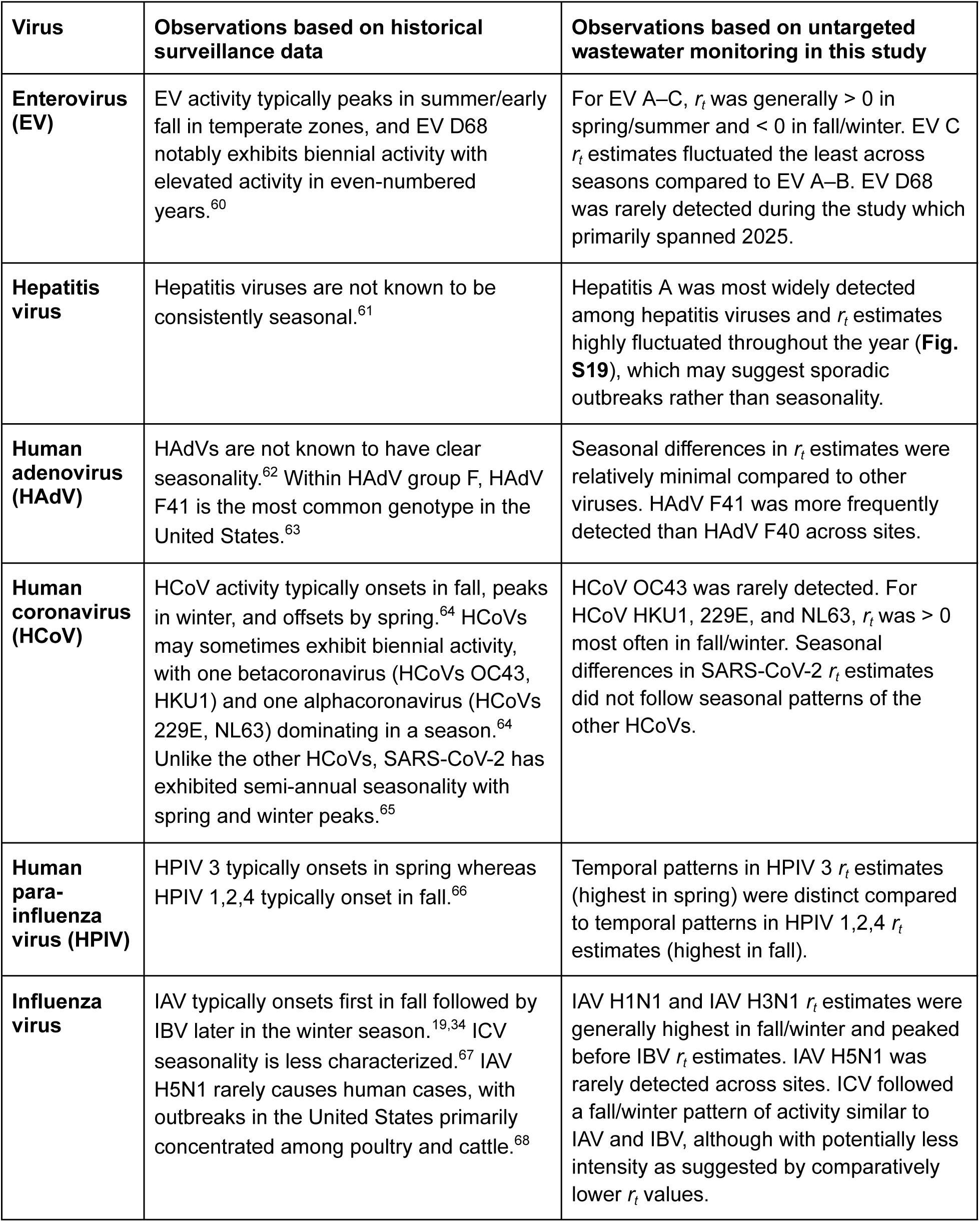
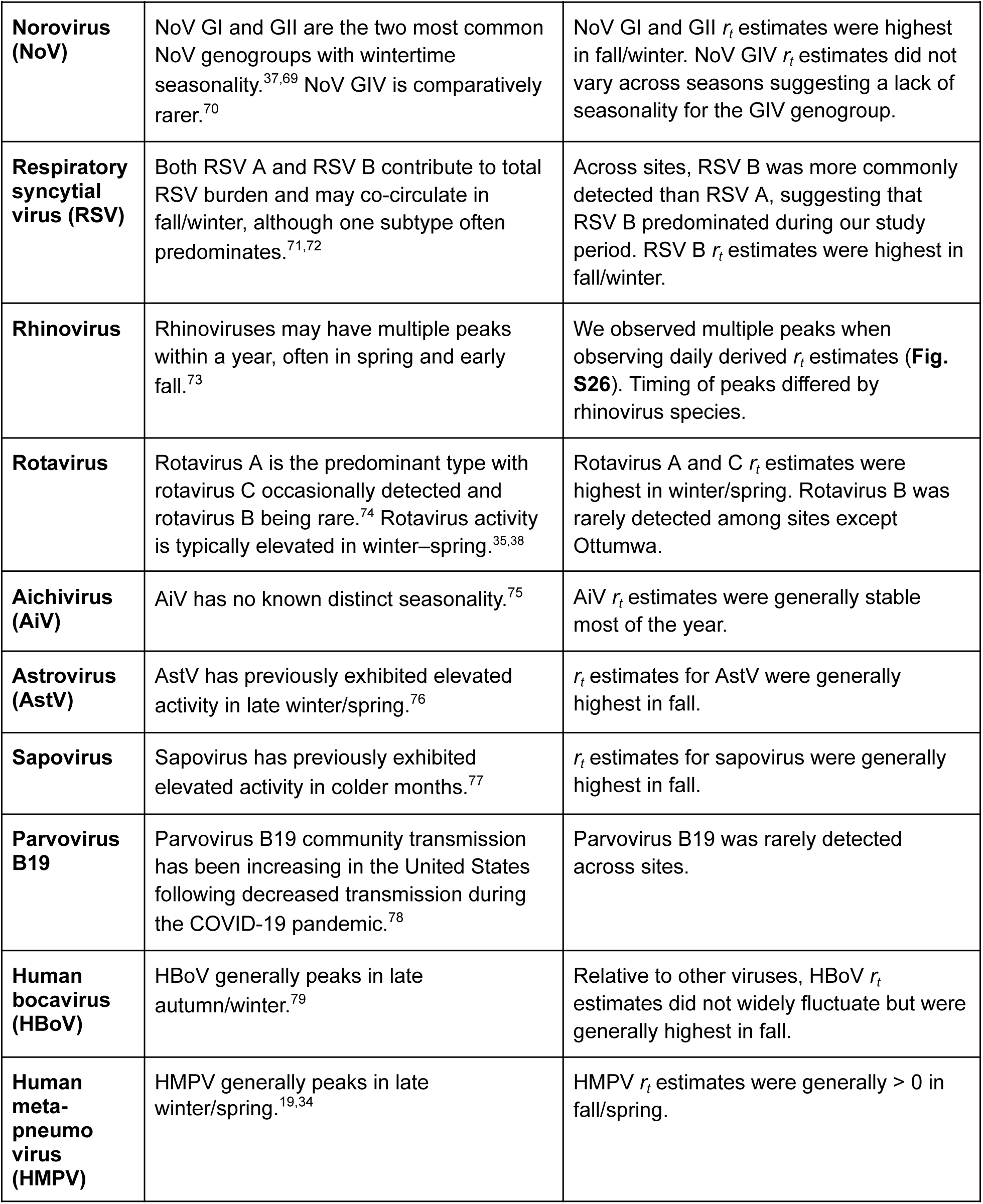
Known epidemiological features of viruses compared to observations in this study

| **Virus** | **Observations based on historical surveillance data** | **Observations based on untargeted wastewater monitoring in this study** |
|---|---|---|
| **Enterovirus (EV)** | EV activity typically peaks in summer/early fall in temperate zones, and EV D68 notably exhibits biennial activity with elevated activity in even-numbered years.<sup>60</sup> | For EV A–C, *r<sub>t</sub>* was generally > 0 in spring/summer and < 0 in fall/winter. EV C *r<sub>t</sub>* estimates fluctuated the least across seasons compared to EV A–B. EV D68 was rarely detected during the study which primarily spanned 2025. |
| **Hepatitis virus** | Hepatitis viruses are not known to be consistently seasonal.<sup>61</sup> | Hepatitis A was most widely detected among hepatitis viruses and *r<sub>t</sub>* estimates highly fluctuated throughout the year (**Fig. S19**), which may suggest sporadic outbreaks rather than seasonality. |
| **Human adenovirus (HAdV)** | HAdVs are not known to have clear seasonality.<sup>62</sup> Within HAdV group F, HAdV F41 is the most common genotype in the United States.<sup>63</sup> | Seasonal differences in *r<sub>t</sub>* estimates were relatively minimal compared to other viruses. HAdV F41 was more frequently detected than HAdV F40 across sites. |
| **Human coronavirus (HCoV)** | HCoV activity typically onsets in fall, peaks in winter, and offsets by spring.<sup>64</sup> HCoVs may sometimes exhibit biennial activity, with one betacoronavirus (HCoVs OC43, HKU1) and one alphacoronavirus (HCoVs 229E, NL63) dominating in a season.<sup>64</sup> Unlike the other HCoVs, SARS-CoV-2 has exhibited semi-annual seasonality with spring and winter peaks.<sup>65</sup> | HCoV OC43 was rarely detected. For HCoV HKU1, 229E, and NL63, *r<sub>t</sub>* was > 0 most often in fall/winter. Seasonal differences in SARS-CoV-2 *r<sub>t</sub>* estimates did not follow seasonal patterns of the other HCoVs. |
| **Human para-influenza virus (HPIV)** | HPIV 3 typically onsets in spring whereas HPIV 1,2,4 typically onset in fall.<sup>66</sup> | Temporal patterns in HPIV 3 *r<sub>t</sub>* estimates (highest in spring) were distinct compared to temporal patterns in HPIV 1,2,4 *r<sub>t</sub>* estimates (highest in fall). |
| **Influenza virus** | IAV typically onsets first in fall followed by IBV later in the winter season.<sup>19,34</sup> ICV seasonality is less characterized.<sup>67</sup> IAV H5N1 rarely causes human cases, with outbreaks in the United States primarily concentrated among poultry and cattle.<sup>68</sup> | IAV H1N1 and IAV H3N1 *r<sub>t</sub>* estimates were generally highest in fall/winter and peaked before IBV *r<sub>t</sub>* estimates. IAV H5N1 was rarely detected across sites. ICV followed a fall/winter pattern of activity similar to IAV and IBV, although with potentially less intensity as suggested by comparatively lower *r<sub>t</sub>* values. |
| **Norovirus (NoV)** | NoV GI and GII are the two most common NoV genogroups with wintertime seasonality.<sup>37,69</sup> NoV GIV is comparatively rarer.<sup>70</sup> | NoV GI and GII *r<sub>t</sub>* estimates were highest in fall/winter. NoV GIV *r<sub>t</sub>* estimates did not vary across seasons suggesting a lack of seasonality for the GIV genogroup. |
| **Respiratory syncytial virus (RSV)** | Both RSV A and RSV B contribute to total RSV burden and may co-circulate in fall/winter, although one subtype often predominates.<sup>71,72</sup> | Across sites, RSV B was more commonly detected than RSV A, suggesting that RSV B predominated during our study period. RSV B *r<sub>t</sub>* estimates were highest in fall/winter. |
| **Rhinovirus** | Rhinoviruses may have multiple peaks within a year, often in spring and early fall.<sup>73</sup> | We observed multiple peaks when observing daily derived *r<sub>t</sub>* estimates (**Fig. S26**). Timing of peaks differed by rhinovirus species. |
| **Rotavirus** | Rotavirus A is the predominant type with rotavirus C occasionally detected and rotavirus B being rare.<sup>74</sup> Rotavirus activity is typically elevated in winter–spring.<sup>35,38</sup> | Rotavirus A and C *r<sub>t</sub>* estimates were highest in winter/spring. Rotavirus B was rarely detected among sites except Ottumwa. |
| **Aichivirus (AiV)** | AiV has no known distinct seasonality.<sup>75</sup> | AiV *r<sub>t</sub>* estimates were generally stable most of the year. |
| **Astrovirus (AstV)** | AstV has previously exhibited elevated activity in late winter/spring.<sup>76</sup> | *r<sub>t</sub>* estimates for AstV were generally highest in fall. |
| **Sapovirus** | Sapovirus has previously exhibited elevated activity in colder months.<sup>77</sup> | *r<sub>t</sub>* estimates for sapovirus were generally highest in fall. |
| **Parvovirus B19** | Parvovirus B19 community transmission has been increasing in the United States following decreased transmission during the COVID-19 pandemic.<sup>78</sup> | Parvovirus B19 was rarely detected across sites. |
| **Human bocavirus (HBoV)** | HBoV generally peaks in late autumn/winter.<sup>79</sup> | Relative to other viruses, HBoV *r<sub>t</sub>* estimates did not widely fluctuate but were generally highest in fall. |
| **Human meta-pneumovirus (HMPV)** | HMPV generally peaks in late winter/spring.<sup>19,34</sup> | HMPV *r<sub>t</sub>* estimates were generally > 0 in fall/spring. |

While our study highlights a compelling use of wastewater metagenomic sequencing, it has limitations. Foremost, longitudinal shedding concentration data are not available for most viruses. Viruses rarely detected may be widely present in a population but not shed into wastewater in detectable quantities. Moreover, viruses with low read counts may be widely present but shed in low quantities (e.g., respiratory viruses), making it more challenging to differentiate signal from noise. Second, metagenomic read counts are influenced by genome size, nucleic acid extraction efficiency, and sequencing depth, and it is possible reads may be incorrectly mapped to reference genomes or that reference genomes may be sparse. However, estimates derived from untargeted wastewater monitoring were concordant with those derived from the other data sources for the sequencing depth and normalization used. Still, future work should repeat analyses using other normalization approaches or probe-based hybrid capture “semi-targeted” wastewater monitoring data which reduce sequencing depth by enriching for viral targets.^24,42–49^ Third, the frequency of targeted wastewater monitoring was higher than that of clinical and untargeted wastewater monitoring data which may have introduced more day-to-day variability in derived estimates. Nonetheless, derived estimates from targeted wastewater monitoring data remained similar at downsampled frequencies (**Figs. S30–31**) although future work could investigate optimal metagenomic sequencing frequencies for inferring transmission dynamics. Lastly, EpiNow2 was readily adaptable to other types of surveillance data but ultimately built for clinical data. While EpiNow2 generates real-time *R_t_* estimates, these estimates are highly affected when recent observations are outliers as are more common in environmental data (**Fig. S32**). Real-time outlier detection methods have been proposed for some targeted wastewater monitoring program setups,^50^ so further developing methods for other program setups would support prospective monitoring. Evidently, if wastewater metagenomic sequencing expands, developing real-time estimation tools tailored specifically to the data would support public health adoption. Estimation tools for targeted wastewater monitoring data exist but are often data intensive, requiring PCR and sample metadata we did not have.^39,51,52^ No tools for untargeted wastewater monitoring data yet exist but could account for sequencing depth and data processing time lags unique to untargeted wastewater monitoring data.

Notwithstanding, our study is the first of our knowledge to estimate *r_t_* and *R_t_* from untargeted wastewater monitoring data and evaluate concordance with existing surveillance data using permutation analysis within a Bayesian framework. We highlight a practical use case for wastewater metagenomic reads for tracking transmission dynamics of known viruses, including those not widely monitored via other surveillance means. As interest and planned investment in wastewater metagenomic sequencing grows nationally, our work emphasizes that broader public health value that may be derived from the data, ranging from early detection of novel pathogens to routine surveillance of endemic pathogens.

## Methods

We estimated *r_t_* and *R_t_* for several human pathogenic viruses using clinical surveillance, targeted wastewater monitoring, and untargeted wastewater monitoring data. *r_t_*, defined as the instantaneous rate of change of log-transformed case incidence, describes the speed at which cases are increasing (*r_t_* > 0) or decreasing (*r_t_* < 0) in a population at time *t*.^25^ *R_t_*, defined as the average number of secondary infections caused by a single infection at time *t*, describes whether an epidemic is growing (*R_t_* > 1) or shrinking (*R_t_*< 1).^25^ For viruses with two or more surveillance data sources available, we then assessed concordance among the time-varying *r_t_* and *R_t_* estimates derived from each data source. An overview of the data sources, *r_t_*and *R_t_* estimation approaches, and concordance metrics is provided below. We conducted the analysis for the following locations in the United States during periods with routine untargeted wastewater monitoring: Boise, Idaho (November 2024–March 2026); Boston, Massachusetts (September 2024–March 2026); Riverside, California (December 2024–March 2026); and Ottumwa, Iowa (February 2025–March 2026). These sites were selected because they had at least one year of routine untargeted wastewater monitoring data, alongside at least six concurrent months of routine targeted wastewater monitoring data, from the data sources described below. All analyses were conducted in R (version 4.6.0). Data and code are available at https://github.com/petal-code/casper_rt_paper.^53^ We used Claude Code Opus 5.5 to assist in drafting analytic code, replicating analyses across multiple sites and viruses, and structuring the GitHub repository. All code was manually reviewed and validated by the authors.

### Overview of surveillance data sources

In this study, we used clinical surveillance, targeted wastewater monitoring, and untargeted wastewater monitoring data to estimate *r_t_*and *R_t_*. Human pathogenic viruses included in this study, along with available surveillance data sources, are listed in **Fig. 1**.

Clinical surveillance data were obtained from Epic Cosmos (Epic Systems Corporation, Verona, WI, USA). Epic Cosmos is an electronic health record data aggregation platform that includes data from over 300 million patients across more than 2,000 hospitals and 52,000 clinics; it is representative of patients seeking healthcare in the United States.^29^ Previously, Epic Cosmos data were shown to be less prone to week-to-week variability compared to sentinel, voluntary reporting systems (e.g., the National Respiratory and Enteric Virus Surveillance System) and attainable at localized spatial scales.^35^ For viruses in **Fig. 1**, aside from influenza A and B virus (IAV/IBV), we obtained weekly totals of patient encounters resulting in an ICD-10 (International Classification of Diseases, 10th revision code) diagnosis (**Table S1**). For IAV and IBV, we obtained weekly totals of positive lab results since ICD-10 influenza diagnoses do not always differentiate between IAV versus IBV. We obtained data for the corresponding county (or state, when county-level data were sparse) of wastewater treatment plants included in this study during the period of untargeted wastewater monitoring (**Table S2**).

Both targeted and untargeted wastewater monitoring data were obtained from the same wastewater treatment plants at each location (**Table S2**). Targeted wastewater monitoring data between July 2024–September 2025 were obtained from Boehm et al.^21^; we used the subset of data concurrent with untargeted wastewater monitoring for each site. Refer to the data descriptor publication for complete methodological details but, briefly, concentrations for all viruses were measured in wastewater solids in units of gene copies per gram of dry weight (cp/g-dry) approximately three times per week using droplet digital reverse-transcription– polymerase chain reaction (ddRT-PCR).^21^ For this analysis, we (1) imputed non-detect measurements with half the assay limit of detection (LOD = 1,000 gc/g-dry) and (2) normalized human pathogen viral nucleic acids by pepper mild mottle viruses (PMMoV) RNA concentrations in the same sample.^21^ PMMoV is an endogenous control originating in the human diet; PMMoV-normalization serves to correct for differences in human fecal strength and viral recovery across wastewater samples.^54–56^

Untargeted wastewater surveillance data were obtained from Justen et al.^23,24^ from the same wastewater treatment plants with available targeted wastewater monitoring data. Routine untargeted wastewater monitoring began at study sites between September 2024–February 2025; we obtained data through March 2026. Refer to the data descriptor for complete methodological details but, briefly, wastewater influent samples were collected and sequenced approximately one time per week using Illumina NovaSeq platforms.^23^ For this analysis, we only used sequencing data from the University of Missouri protocol.^23^ Bioinformatic processing included shallow taxonomic profiling on a subset of reads and deep vertebrate-infecting virus profiling on all reads following a published metagenomics pipeline by Bradshaw et al.^57^ (https://github.com/securebio/nao-mgs-workflow). For this analysis, we (1) imputed samples with 0 read counts for a viral target with half the LOD, herein defined as the lowest possible number of read counts (LOD = 1 read), and (2) normalized virus read counts by scaled PMMoV counts (PMMoV counts ✕ total read pairs / number of reads profiled) to account for fluctuations in human fecal strength, viral recovery, and sequencing depth.^23^ Because human pathogenic viruses are rare considering all microbes present in a wastewater sample, we assumed absolute clade counts are approximately proportional to relative abundance. Additionally, we opted to normalize virus read counts by PMMoV read counts for consistency with the targeted wastewater monitoring data. Preliminary work comparing normalization approaches for wastewater metagenomic sequencing data suggests normalization by tobamovirus read counts, the genus of PMMoV, may be most preferable although PMMoV normalization was still among the best performing normalization markers.^24^

### Assessing temporal association across surveillance data sources

For viruses with surveillance data from multiple sources (**Fig. 1**), we evaluated the null hypothesis that weekly surveillance signals (i.e., clinical counts, concentrations of viral nucleic acids, or virus-specific read counts) are not temporally correlated. We used nonparametric Kendall’s tau correlation test as surveillance time series were not always normally distributed (Shapiro-Wilk, p < 0.01). We excluded any wastewater time series where >75% of samples were below the LOD. Virus presence or absence was instead reported for these rarely detected viruses in subsequent analyses, rather than *r_t_* or *R_t_* estimates. Furthermore, because targeted wastewater monitoring data were reported multiple times per week, we computed weekly average concentrations prior to assessing correlation. For each virus, we assessed correlations at each location and between each pair of available data sources. Therefore, we used an adjusted alpha value of 0.05 divided by the total number of tests conducted (i.e., the number of locations times the number of distinct data source pairs with sufficient detections) to account for multiple testing when interpreting *p* values for each virus.

### Estimating instantaneous epidemic growth rate (*r_t_*)

We estimated *r_t_* using all available data sources for viruses as *r_t_* does not require assumptions about disease transmission. *r_t_* is defined as the logarithmic derivative of smoothed incidence.^26^ The clinical surveillance data represent incident infections; we assumed wastewater measurements similarly represent incident infections as wastewater signals have been previously shown to positively correlate with disease incidence.^4,23^

To smooth each surveillance time series, we fit a generalized additive model (GAM) using the mgcv R package.^30^ Specifically, we used a negative binomial distribution for clinical data, which were discrete counts, and a tweedie distribution for both targeted and untargeted wastewater monitoring data, which were continuous and positive. Additionally, we removed outlier wastewater measurements prior to smoothing; set *k*, the dimension basis of the smooth term, generously (i.e., the number of observations remaining in the time series divided by 4); and used REML (restricted maximum likelihood) as the smoothing parameter estimation method with the outer BFGS (Broyden–Fletcher–Goldfarb–Shanno) optimization algorithm.^30^ To remove wastewater outliers, we first calculated the median and median absolute deviation (MAD) of log_10_-transformed wastewater measurements above the LOD in the time series. We then calculated the modified z-score for each log_10_-transformed measurement using this median and MAD and defined outliers as measurements with a modified z-score above 3.5.^58^ We assumed all defaults otherwise. Refer to **Fig. S32** for smoothed time series without outlier removal for comparison.

GAMs implemented using the mgcv R package have an empirical Bayesian interpretation as the smoothing parameter estimated via REML uses a frequentist approach and the uncertainty calculation to obtain credible intervals (CrIs) uses a Bayesian approach. Thus, for each GAM fit, we extracted 1,000 posterior draws and estimated the median and 90% CrI each day to produce a smoothed time series. Next, to estimate daily *r_t_* from each smoothed time series, we approximated the logarithmic derivative of the smoothed time series *S_t_* using **Equation 1** with a daily time step. We approximated the logarithmic derivative each day using all 1,000 posterior draws to calculate the median and 90% CrI of the *r_t_* estimate.

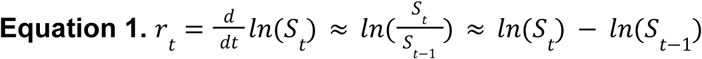

### Estimating time-varying effective reproduction number (*R_t_*)

Estimating *R_t_* requires an epidemiological model to link an observed surveillance signal to latent infections in the population. Therefore, for each location, we estimated *R_t_* only for viruses with sufficient data to inform model parameters: SARS-CoV-2, IAV, IBV, RSV, NoV, and rotavirus. For each virus, we estimated *R_t_*from each available surveillance data source using the renewal equation model in a Bayesian framework, as described by the R package EpiNow2 (**Equation 2**).^31,32^ The United States Centers for Disease Control and Prevention (CDC) similarly used EpiNow2 to estimate *R_t_* from emergency department visit data between 2022–May 2026.^59^

According to the renewal equation, latent infections each day *I_t_*are given by the total infectiousness of infected individuals on day *t*, multiplied by the effective reproduction number *R_t_*. Total infectiousness is described by past incidence and an infected individual’s infectivity profile or generation time *w* probability mass function (pmf), both of which depend on the time since infection *τ_t_*.

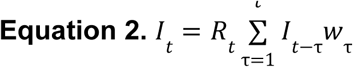

Latent infections are then convolved to expected case reports *D ^c^* according to **Equation 3**. Specifically, *ζ* is the infection-to-reporting delay pmf, which is a combination of incubation period and reporting delays for clinical data, and *α*_c_ is a scaling factor representing ascertainment of cases.

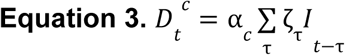

Next, expected case reports *D_t_^c^* are mapped to observed case reports *C_t_^c^*using a negative binomial distribution, where *ψ* is an overdispersion factor (**Equation 4**).

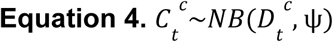

To control the smoothness of *R_t_* estimates, we used the default Gaussian process with the default length scale and magnitude parameters. Note we did not remove outlier wastewater measurements prior to estimating *R_t_* because Gaussian process smoothing is less sensitive to extrema compared to GAM smoothing. Furthermore, the Gaussian process smoothing allows EpiNow2 to still generate daily *R_t_* estimates even with nondaily observed data. Refer to the EpiNow2 documentation for a complete overview of the renewal equation modeling framework.^32^

Although EpiNow2 is designed to estimate *R_t_* from case counts, we adapted the framework to estimate *R_t_*from both targeted and untargeted wastewater monitoring data. Namely, *D ^ww^* and *C ^ww^* instead represent the expected and observed wastewater signal, respectively, arising from latent infections *I_t_* in this adapted framework (**Equation 5–6**). The wastewater signal is PMMoV-normalized concentrations of viral nucleic acids using targeted wastewater monitoring data and PMMoV-normalized virus clade counts scaled to account for sequencing depth using untargeted wastewater monitoring data. EpiNow2 can only handle integer counts; therefore, we multiplied wastewater signals in a time series by a constant factor and rounded such that they represented integer counts analogous to clinical data. When both clinical and wastewater data were available for a given site and virus, we multiplied wastewater signals by a factor that resulted in the median rounded wastewater signal being on the same order of magnitude as the median clinical count. When only wastewater data were available, we multiplied wastewater signals so the median rounded wastewater signal was above one and on the same order of magnitude as the other wastewater data source, if both wastewater data sources were available. *R_t_* is primarily affected by relative changes in the surveillance signal rather than absolute changes. Thus, *R_t_* estimates are robust to the choice of multiplication factor (see **Fig. S33** for a comparison of different choices of this multiplication factor). Next, to represent the data generating process associated with wastewater monitoring data, we replaced the infection-to-reporting delay with pathogen-specific infection shedding dynamics which describe the time-varying evolution of virus shedding following infection. We parameterized shedding dynamics using a factor representing the total amount of viral nucleic acids shed over the course of an infection (⍺*_ww_*) and a time-varying pmf describing the fraction of the total amount shed on a given day following infection (*s_τ_*). We assumed decay, sewer transport time, and shedding of viral nucleic acids from other human excreta were negligible.

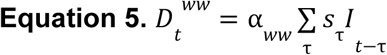

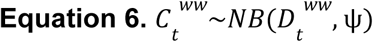

A complete overview of the parameters and priors used in the modeling framework for each virus is provided in **Table S4**. Briefly, generation time, incubation period, reporting delay, and fecal shedding load distributions for all viruses were obtained from the literature. Longitudinal fecal shedding load data do not exist for RSV; we assumed fecal shedding of RSV RNA follows the same distribution as SARS-CoV-2. We also used a fixed distribution at 1 for the scaling factors (⍺*_c_* and ⍺*_ww_*) for all viruses because we only aimed to estimate *R_t_*, or relative changes in the surveillance signal, rather than actual case counts or wastewater measurements. For SARS-CoV-2, IAV, and RSV, we additionally conducted a sensitivity analysis using alternative fecal shedding distributions from the literature (**Table S7**).

### Assessing concordance in time-varying estimates derived from different surveillance data sources

When we estimated daily *r_t_* and *R_t_*from two or more surveillance data sources, we assessed concordance among the time-varying estimates between all pairs of data sources. We used three complementary concordance metrics: (1) mean absolute error (MAE), (2) agreement on binary (AOB), and (3) correlation. MAE (**Equation 7**) quantifies numerical agreement between two time series. AOB (**Equation 8**) assesses agreement on epidemic direction between two time series; we used thresholds *ɸ* of 0 (AOB0) and 1 (AOB1) for *r_t_* and *R_t_*, respectively, as these thresholds are used to assess whether an epidemic is growing or shrinking for each metric. In **Equations 7–8**, *y* represents the derived time-varying estimate each day *t* from data source *a* and *b* and *n* represents the total number of concurrent days between these same data sources. Correlation evaluates whether two time series are temporally associated. Given that time series estimates may not always be normally distributed, we used Kendall’s tau correlation to calculate correlation coefficients. Positive tau values indicate positive temporal association between the daily time-varying estimates derived from data source *a* and *b* whereas negative tau values indicate negative temporal association.

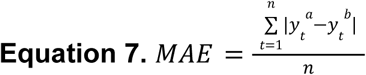

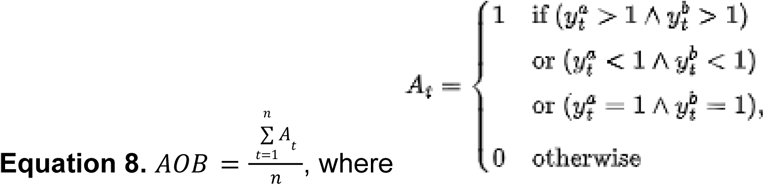

Specifically, for both *r_t_* and *R_t_*, we calculated MAE, AOB, and Kendall’s tau between all pairs of posterior draws derived from data source *a* and *b*. Doing so generated an observed posterior distribution for each concordance metric and each pair of data sources. Since each surveillance data source has unique biases and limitations, no surveillance data source represents a gold standard. Therefore, to assess whether two surveillance data sources generated significantly similar *r_t_* and *R_t_* estimates than would be expected by random chance, we generated null posterior distributions for each pair of data sources using Bayesian permutation analysis. Specifically, we permuted each posterior draw across time points of data source *b* and recalculated each metric between data source *a* and *b_perm_*, generating null posterior distributions. For each pair of data sources and concordance metric, the null posterior distribution represented the posterior distribution under a null hypothesis of no temporal concordance or relationship between the two data sources. We then calculated Bayesian permutation *p* values which represent the proportion of posterior draws in which the null scenario was more similar than the observed scenario. For MAE, values closer to 0 indicate similarity, so we calculated permutation *p* values as the proportion of null draw pairs less than observed draw pairs. In contrast, AOB and tau coefficients closer to 1 indicate similarity; we calculated permutation p values as the proportion of null draw pairs greater than observed draw pairs. We interpreted permutation *p* values < 0.05 as suggesting significant concordance between two data sources as this indicated the null hypothesis was not rejected in less than 5% of posterior draw pairs.

## Data availability

All datasets used in this study are described and cited in the text. Clinical surveillance data were obtained from Epic Cosmos by LSM and interpreted by RYL. Access can be obtained through approved users with affiliation at a participating organization or by contacting Epic Cosmos. Targeted wastewater monitoring data were obtained from Boehm et al. 2026 (https://doi.org/10.1016/j.dib.2026.112503). These data were collected as part of the WastewaterSCAN / SCAN project, a partnership between Stanford University, Emory University, and Verily funded philanthropically through a gift to Stanford University. Untargeted wastewater monitoring data were obtained from Justen et al. 2026 (https://doi.org/10.64898/2026.07.14.26356442). Raw sequencing data are available under NCBI BioProject accession PRJNA1247874 and the dataset is described in Justen et al. 2026 (https://doi.org/10.64898/2026.03.05.26345726). Publicly available datasets are on GitHub at https://github.com/petal-code/casper_rt_paper and Zenodo at https://doi.org/10.5281/zenodo.24117526.

## Code availability

The code to reproduce the analysis is publicly available on GitHub at https://github.com/petal-code/casper_rt_paper and Zenodo at https://doi.org/10.5281/zenodo.24117526. All analyses were conducted in R 4.6.0 and all R package versions are recorded in the GitHub repository.

## Acknowledgements

We thank the participating wastewater treatment plant and laboratory staff for collecting and analyzing samples used in this study. We especially thank the Coalition for Agnostic Sequencing of Pathogens from Environmental Reservoirs (CASPER) network for sharing wastewater metagenomic sequencing data and helpful insight.

## Author information

### Authors and affiliations

Division of Infectious Diseases and Vaccinology, School of Public Health, University of California, Berkeley, Berkeley, CA, USA

Elana M. G. Chan

Charles Whittaker

Massachusetts Institute of Technology, Media Lab, Cambridge, MA, USA

Lennart Justen

Draper Scholar, The Charles Stark Draper Laboratory, Inc., Cambridge, MA, USA

Lennart Justen

Broad Institute of MIT and Harvard, Cambridge, MA, USA

Lennart Justen

SecureBio, Cambridge, MA, USA

Alessandro Zulli Michael R. McLaren

Division of Infectious Diseases, Department of Medicine, David Geffen School of Medicine, University of California, Los Angeles, Los Angeles, CA, USA

Rebecca Y. Linfield

Department of Neurology, Stanford University, Palo Alto, CA, USA

Leon S. Moskatel

## Contributions

**EMGC**: Conceptualization, Data curation, Formal analysis, Investigation, Methodology, Project administration, Software, Supervision, Validation, Visualization, Writing -original draft, Writing -reviewing & editing **LJ:** Data curation, Resources, Writing -reviewing & editing **AZ:** Data curation, Resources **MRM:** Data curation, Resources, Writing -reviewing & editing **RYL**: Data curation, Resources, Writing -reviewing & editing **LSM**: Data curation, Resources **CW**: Conceptualization, Funding acquisition, Methodology, Project administration, Supervision, Validation, Writing -reviewing & editing All authors approved the final version of this manuscript.

Corresponding author Correspondence to Prof. Charles Whittaker.

## Supplementary information

The Supplementary Information PDF contains Supplementary Tables 1–7. and Figs. 1–33.

