## Supporting Information for "Wastewater metagenomic sequencing quantitatively tracks transmission dynamics for human pathogenic viruses"

Fig. S2. IAV surveillance data and $R_t$ estimates for all locations. 14

Fig. S3. IAV surveillance data and $r_t$ estimates for all locations. 15

Fig. S4. IBV surveillance data and $R_t$ estimates for all locations. 16

Fig. S5. IBV surveillance data and $R_t$ estimates for Boston, Massachusetts without y-axis adjustment. 17

Fig. S6. IBV surveillance data and $r_t$ estimates for all locations. 18

Fig. S7. RSV surveillance data and $R_t$ estimates for all locations. 19

Fig. S8. RSV surveillance data and $r_t$ estimates for all locations. 20

Fig. S9. NoV surveillance data and $R_t$ estimates for all locations. 21

Fig. S10. NoV surveillance data and $r_t$ estimates for all locations. 22

Fig. S11. Rotavirus surveillance data and $R_t$ estimates for all locations. 23

Fig. S12. Rotavirus surveillance data and $r_t$ estimates for all locations. 24

Table S5. Observed posterior distributions for metrics of concordance among derived $R_t$ estimates. 25

Fig. S13. Concordance between $r_t$ estimates derived from different data sources. 34

Table S6. Observed posterior distributions for metrics of concordance among derived $r_t$ estimates. 35

Fig. S14. Hepatitis A surveillance data and $r_t$ estimates for all locations. 40

Fig. S15. Influenza A H1N1 surveillance data and $r_t$ estimates for all locations. 41

Fig. S16. Human metapneumovirus surveillance data and $r_t$ estimates for all locations. 42

Table S7. Alternative fecal shedding load distributions used for $R_t$ sensitivity analyses. 43

Fig. S17. Concordance between $R_t$ estimates derived from different data sources using alternative shedding distributions, Boston, Massachusetts. 44

Fig. S18. Untargeted enterovirus wastewater monitoring data and $r_t$ estimates for all locations, differentiated by species/type. 45

Fig. S19. Untargeted hepatitis virus wastewater monitoring data and $r_t$ estimates for all locations, differentiated by virus. 46

Fig. S20. Untargeted HAdV wastewater monitoring data and $r_t$ estimates for all locations, differentiated by species/type. 47

Fig. S21. Untargeted HCoV wastewater monitoring data and $r_t$ estimates for all locations, differentiated by species. 48

Fig. S22. Untargeted HPIV wastewater monitoring data and $r_t$ estimates for all locations, differentiated by type. 49

**Table S1. International Classification of Diseases, 10th Revision (ICD-10) diagnosis codes**

| Virus | ICD-10 diagnosis code(s) |
|---|---|
| SARS-CoV-2 | U07: COVID-19 |
| Respiratory syncytial virus (RSV) | B97.4: RSV<br>J12.1: RSV pneumonia<br>J20.5: Acute bronchitis due to RSV<br>J21.0: Acute bronchiolitis due to RSV |
| Norovirus (all genogroups) | A08.11: Acute gastroenteropathy due to Norwalk agent |
| Rotavirus | A08.0: Rotaviral enteritis |

For influenza, we obtained positive lab results (differentiated by influenza A vs influenza B) from Epic Cosmos instead of diagnosis encounters, which do not always differentiate by influenza type.

**Table S2. Study locations, corresponding wastewater treatment plants, and Epic Cosmos spatial scales**

| Location | Wastewater treatment plant(s) | Wastewater treatment plant population[a] | County of wastewater treatment plant(s) | Virus type[b] | Epic Cosmos spatial scale |
|---|---|---|---|---|---|
| Boise, Idaho[c] | Lander Street Water Renewal Facility<br>West Boise Water Renewal Facility | 108,556<br>186,901 | Ada county | Respiratory | County |
| | | | | Enteric | State |
| Boston, Massachusetts | Deer Island Treatment Plant[d] | 2,400,000 | Suffolk county | Respiratory | County |
| | | | | Enteric | State |
| Ottumwa, Iowa | Ottumwa WPCF | 25,529 | Wapello county | Respiratory | State |
| | | | | Enteric | State |
| Riverside, California | Riverside Water Quality Control Plant | 350,000 | Riverside county | Respiratory | County |
| | | | | Enteric | State |

[a] As reported in the targeted wastewater monitoring data descriptor by Boehm et al.[1]

[b] Respiratory viruses include SARS-CoV-2, influenza A, influenza B, and respiratory syncytial virus; enteric viruses include norovirus and rotavirus.

[c] In the targeted wastewater monitoring dataset, samples were collected, analyzed, and reported separately from both plants. In the untargeted wastewater monitoring dataset, a composite wastewater sample from both plants was collected and analyzed; data were reported for one combined site. Therefore, in the targeted wastewater monitoring dataset, we aggregated reported concentrations from the two sites using a population-weighted average as described by Chan et al.[2]

[d] In the untargeted wastewater monitoring dataset only, samples were collected and analyzed separately from the north and south catchments of the plant. Therefore, in the untargeted wastewater monitoring dataset, we aggregated reported read counts from the two catchments by summing values prior to downstream analysis. Also note, this plant includes samples sequenced by both the MU and SB laboratories (see Justen et al.[3] for further details). For this analysis, we only used MU-sequenced samples as samples from all other sites were sequenced by the MU laboratory as well.

**Table S3. Kendall's tau correlations assessing temporal association between data sources**

| Virus | Location | Weeks | Comparison[a] | Tau | *p* value[b] |
|---|---|---|---|---|---|
| SARS-CoV-2 | Boston, MA | 54 | Clinical vs targeted wastewater | 0.389 | **< 0.001** |
| | | 76 | Clinical vs untargeted wastewater | 0.545 | **< 0.001** |
| | | 49 | Targeted vs untargeted wastewater | 0.340 | **< 0.001** |
| | Boise, ID | 47 | Clinical vs targeted wastewater | 0.441 | **< 0.001** |
| | | 69 | Clinical vs untargeted wastewater | 0.365 | **< 0.001** |
| | | 42 | Targeted vs untargeted wastewater | 0.303 | 0.004 |
| | Ottumwa, IA | 33 | Clinical vs targeted wastewater | 0.398 | **0.001** |
| | | 58 | Clinical vs untargeted wastewater | 0.470 | **< 0.001** |
| | | 31 | Targeted vs untargeted wastewater | 0.475 | **< 0.001** |
| | Riverside, CA | 42 | Clinical vs targeted wastewater | 0.497 | **< 0.001** |
| | | 68 | Clinical vs untargeted wastewater | 0.754 | **< 0.001** |
| | | 41 | Targeted vs untargeted wastewater | 0.463 | **< 0.001** |
| Influenza A | Boston, MA | 54 | Clinical vs targeted wastewater | 0.714 | **< 0.001** |
| | | 76 | Clinical vs untargeted wastewater | 0.603 | **< 0.001** |
| | | 49 | Targeted vs untargeted wastewater | 0.583 | **< 0.001** |
| | Boise, ID | 47 | Clinical vs targeted wastewater | 0.634 | **< 0.001** |
| | | 69 | Clinical vs untargeted wastewater | 0.534 | **< 0.001** |
| | | 42 | Targeted vs untargeted wastewater | 0.600 | **< 0.001** |
| | Ottumwa, IA | 33 | Clinical vs targeted wastewater | 0.579 | **< 0.001** |
| | | 58 | Clinical vs untargeted wastewater | 0.266 | **0.003** |
| | | 31 | Targeted vs untargeted wastewater | 0.428 | **0.001** |
| | Riverside, CA | 42 | Clinical vs targeted wastewater | 0.608 | **< 0.001** |
| | | 68 | Clinical vs untargeted wastewater | 0.565 | **< 0.001** |
| | | 41 | Targeted vs untargeted wastewater | 0.563 | **< 0.001** |
| Influenza B | Boston, MA | 54 | Clinical vs targeted wastewater | 0.374 | **< 0.001** |
| | | 76 | Clinical vs untargeted wastewater | 0.400 | **< 0.001** |
| | | 49 | Targeted vs untargeted wastewater | 0.568 | **< 0.001** |
| | Boise, ID | 47 | Clinical vs targeted wastewater | 0.453 | **< 0.001** |
| | | 69 | Clinical vs untargeted wastewater | 0.535 | **< 0.001** |
| | | 42 | Targeted vs untargeted wastewater | 0.361 | **0.001** |
| | Ottumwa, IA | 33 | Clinical vs targeted wastewater | 0.482 | **< 0.001** |
| | Riverside, CA | 42 | Clinical vs targeted wastewater | 0.473 | **< 0.001** |
| | | 68 | Clinical vs untargeted wastewater | 0.495 | **< 0.001** |
| | | 41 | Targeted vs untargeted wastewater | 0.554 | **< 0.001** |
| Respiratory Syncytial Virus | Boston, MA | 54 | Clinical vs targeted wastewater | 0.599 | **< 0.001** |
| | | 76 | Clinical vs untargeted wastewater | 0.548 | **< 0.001** |
| | | 49 | Targeted vs untargeted wastewater | 0.500 | **< 0.001** |
| | Boise, ID | 47 | Clinical vs targeted wastewater | 0.674 | **< 0.001** |
| | | 69 | Clinical vs untargeted wastewater | 0.433 | **< 0.001** |
| | | 42 | Targeted vs untargeted wastewater | 0.440 | **< 0.001** |
| | Ottumwa, IA | 33 | Clinical vs targeted wastewater | 0.651 | **< 0.001** |

| Virus | Location | Weeks | Comparison[a] | Tau | *p* value[b] |
|---|---|---|---|---|---|
| | | 58 | Clinical vs untargeted wastewater | 0.463 | **< 0.001** |
| | | 31 | Targeted vs untargeted wastewater | 0.437 | **< 0.001** |
| | Riverside, CA | 42 | Clinical vs targeted wastewater | 0.670 | **< 0.001** |
| | | 68 | Clinical vs untargeted wastewater | 0.699 | **< 0.001** |
| | | 41 | Targeted vs untargeted wastewater | 0.576 | **< 0.001** |
| Norovirus[c] | Boston, MA | 54 | Clinical vs targeted wastewater | 0.524 | **< 0.001** |
| | | 76 | Clinical vs untargeted wastewater | 0.553 | **< 0.001** |
| | | 49 | Targeted vs untargeted wastewater | 0.827 | **< 0.001** |
| | Boise, ID | 47 | Clinical vs targeted wastewater | 0.366 | **< 0.001** |
| | | 69 | Clinical vs untargeted wastewater | 0.387 | **< 0.001** |
| | | 42 | Targeted vs untargeted wastewater | 0.691 | **< 0.001** |
| | Ottumwa, IA | 33 | Clinical vs targeted wastewater | 0.269 | 0.031 |
| | | 58 | Clinical vs untargeted wastewater | 0.373 | **< 0.001** |
| | | 31 | Targeted vs untargeted wastewater | 0.484 | **< 0.001** |
| | Riverside, CA | 42 | Clinical vs targeted wastewater | 0.521 | **< 0.001** |
| | | 68 | Clinical vs untargeted wastewater | 0.340 | **< 0.001** |
| | | 41 | Targeted vs untargeted wastewater | 0.732 | **< 0.001** |
| Rotavirus | Boston, MA | 76 | Clinical vs untargeted wastewater | 0.339 | **< 0.001** |
| | Boise, ID | 69 | Clinical vs untargeted wastewater | 0.200 | 0.031 |
| | Ottumwa, IA | 58 | Clinical vs untargeted wastewater | 0.128 | 0.176 |
| | Riverside, CA | 68 | Clinical vs untargeted wastewater | 0.421 | **< 0.001** |
| Human metapneumo-virus | Boston, MA | 49 | Targeted vs untargeted wastewater | 0.325 | **0.001** |
| | Boise, ID | 42 | Targeted vs untargeted wastewater | 0.459 | **< 0.001** |
| Influenza A H1 | Boston, MA | 43 | Targeted vs untargeted wastewater | 0.475 | **< 0.001** |
| | Boise, ID | 40 | Targeted vs untargeted wastewater | 0.523 | **< 0.001** |
| | Ottumwa, IA | 31 | Targeted vs untargeted wastewater | 0.432 | **< 0.001** |
| Hepatitis A | Boston, MA | 49 | Targeted vs untargeted wastewater | 0.463 | **< 0.001** |

[a] We excluded wastewater data sources with >75% of samples below the limit of detection from comparisons.

[b] To account for multiple hypothesis testing for each virus, we used an adjusted alpha value of 0.05 divided by the total number of tests conducted (e.g., 12 tests for SARS-CoV-2) when interpreting *p* values. Bolded *p* values were below the adjusted alpha value for a given virus.

[c] For norovirus, we compared clinical data inclusive of all genogroups to wastewater monitoring data specific to genogroup II. Only genogroup II had available data from both targeted and untargeted wastewater data sources for subsequent analyses and is the predominant norovirus genogroup.

**Table S4. Model parameters used to estimate $R_t$ using EpiNow2**

| Parameter | Definition | Distribution | Source |
|---|---|---|---|
| Generation time (days) | The time between infection in an individual and subsequent infection in an infectee. Because it is hard to assess when an individual is infected, the serial interval—or time between symptom onsets in an infector and infectee—is often used as a proxy. EpiNow2 cannot take in time-varying generation time distributions; we assumed generation time is time invariant across the analysis period of our study. | *SARS-CoV-2:* Gamma(mean = 3.5, sd = 2.1, max = 14)<br><br>*IAV:* Gamma(mean = 3.2, sd = 2.1, max = 10)<br><br>*IBV:* See IAV<br><br>*RSV:* Gamma(mean = 5.6, sd = 4, max = 19)<br><br>*NoV:* Gamma(mean = 3.3, sd = 1.91, max = 9)<br><br>*Rotavirus:* NonParametric(pmf = 0, 0.21, 0.22, 0.23, 0.16, 0.10, 0.08) | *SARS-CoV-2:* Mean and sd of the estimated intrinsic generation time for the Omicron SARS-CoV-2 variant (Dec 2021 – May 2023) based on United States households from Chan et al. (2026).[4]<br><br>*IAV:* Mean and sd of the estimated intrinsic generation time for IAV (2021/2022 – 2022/2023) based on United States households from Chan et al. (2025).[5]<br><br>*IBV:* Assumed to be the same as IAV.[5]<br><br>*RSV:* Estimates from the literature are sparse and mixed.[6–8] We used data from Hall et al. (1976) which represents a middle ground estimate and includes both adults and children.[7] Given only the mean and range were reported,[7] we approximated the sd as the range divided by 4.<br><br>*NoV:* Conservative estimate by Tsang et al. (2018)[9] using data from Götz et al. (2001)[10] as the latter study site is most similar to the United States. Given only the mean and 95% CI were reported, we approximated the sd as the CI width / 3.92 * √n, where n = 39.[9]<br><br>*Rotavirus:* Estimates from the literature are sparse and mixed.[11,12] We extracted the serial interval distribution according to a multiple mixture model by van Gaalen et al. (2017)[11] based on data from Lopman et al. (2013).[13] EpiNow2 cannot accommodate nonzero probabilities for the generation time on day 0; we shifted the distribution by 1 day such that the probability on day 0 is 0. Given that the mean serial interval according to Grimwood et al. (1983)[12] (4.9 days for children and 6.4 days for adults) is larger than the mean of the serial interval in the model by van Gaalen et al.[11] (2.1 days), we believe this decision is sensible. |
| Incubation period (days) | Delay from infection to symptom onset. We convolved the incubation period distribution with the reporting delay distribution to generate the final infection-to-reporting delay distribution when estimating $R_t$ from clinical data. | *SARS-CoV-2:* Gamma(mean = 3.63, sd = 0.83, max = 10)<br><br>*IAV:* Gamma(mean = 1.55, sd = 0.66, max = 5) | *SARS-CoV-2:* Estimates across all Omicron subvariants from a systematic review and meta-analysis by Xu et al. (2023).[14] Only the pooled mean and 95% CI were reported; we approximated the sd as the CI width / 3.92 * √n, where n = 18.[14]<br><br>*IAV:* Mean and sd of the estimated incubation period from Chan et al. (2025)[5] based on a previous systematic review by Lessler et al. (2009).[15] |

| Parameter | Definition | Distribution | Source |
| --- | --- | --- | --- |
| | | *IBV:* Gamma(mean = 0.61, sd = 0.25, max = 5)<br><br>*RSV:* LogNormal(meanlog = 1.48, sdlog = 0.22, max = 8)<br><br>*NoV:* LogNormal(meanlog = 0.18, sdlog = 0.44, max = 7)<br><br>*Rotavirus:* LogNormal(meanlog = 0.69, sdlog = 0.31, max = 7) | *IBV:* Mean and sd of the estimated incubation period from Chan et al. (2025)[5] based on a previous systematic review by Lessler et al. (2009).[15]<br><br>*RSV:* Estimates from a systematic review by Lessler et al. (2009).[15] Median and dispersion (95% CI) estimates are based on a log-normal distribution;[15] we calculated meanlog and sdlog as ln(median) and ln(dispersion), respectively.<br><br>*NoV:* Estimates for genogroup II from a systematic review by Lee et al. (2013).[16] Median and dispersion (95% CI) estimates are based on a log-normal distribution;[16] we calculated meanlog and sdlog as ln(median) and ln(dispersion), respectively.<br><br>*Rotavirus:* Estimates from a systematic review by Lee et al. (2013).[16] Median and dispersion (95% CI) estimates are based on a log-normal distribution;[16] we calculated meanlog and sdlog as ln(median) and ln(dispersion), respectively. |
| Reporting delay (days) | Delay from symptom onset to clinical care. In this study, clinical care refers to the date of positive lab result (for IAV/IBV, assuming results are returned within 24 hr[17,18]) or date of diagnosis encounter (all other pathogens) as obtained from Epic Cosmos.[19] We assumed the reporting delay distribution is time invariant over the study period, and convolved the reporting delay distribution with the incubation period distribution to generate the final infection-to-reporting delay distribution when estimating $R_t$ from clinical data. | NonParametric(pmf = 15.82, 28.28, 20.54, 19.36, 6.73, 6.17, 2.27, 0.83) | Based on a population-based study on healthcare-seeking behaviors among persons with respiratory and gastrointestinal infections in Hong Kong by Zhang et al. (2020).[20] Since we only estimated Rt for respiratory and gastrointestinal pathogens in this study, we assumed the same reporting delay distribution across all viruses. Specifically, we digitized Fig. 3 (all participants, any symptoms) from Zhang et al.[20] to approximate the reporting delay distribution, assuming “immediately” = 0 days, “within 12 hr” = 1 day, “within 2 days” = 2 days, “2–3 days” = 3 days, and “3–4” days = days. For the “more than 4 days” category, we assumed a maximum delay of 7 days with exponential decay of the remaining probability mass between days 5–7. |
| Fecal shedding dynamics (days) | Delay from infection to shedding into wastewater as characterized by longitudinal fecal shedding patterns of viral nucleic acids. Used as the | *SARS-CoV-2:* NonParametric(pmf = 2.427277e-05, 3.853375e-05, 5.279472e-05, 6.705570e-05, | *SARS-CoV-2:* We derived a normalized fecal shedding load pmf from Wannigama et al. (2024)[29]. Specifically, for each SARS-CoV-2 variant (XBB.1.5, EG.5.1, HV.1, JD.1.1, BA.2.86, JN.1), we extracted the median fecal concentration of SARS-CoV-2 RNA (cp/mg) for all |

| Parameter | Definition | Distribution | Source |
|---|---|---|---|
| | infection-to-shedding delay distribution when estimating $R_t$ from wastewater monitoring data. We assumed inputs into the wastestream from other human excreta are negligible relative to feces[21], sewer transport time is negligible (<<24 hours)[22], and decay of viral RNA is negligible and accounted for by PMMoV normalization.[23–28] | 7.197659e-04, 1.372476e-03, 2.025186e-03, 3.459005e-03, 4.892823e-03, 4.822649e-02, 9.156016e-02, 1.363023e-01, 1.810444e-01, 1.529968e-01, 1.249491e-01, 9.690147e-02, 6.885383e-02, 4.080619e-02, 1.275854e-02, 1.068224e-02, 8.605932e-03, 6.529626e-03, 4.453319e-03, 2.377013e-03, 3.007065e-04) | reported days after symptom onset. We then calculated the mean fecal concentration each day across all variants and imputed values on missing days using linear interpolation. Since the first fecal samples were collected 3 days before symptom onset (approximately the incubation period length), we assumed the derived normalized pmf represents days since infection. |
| | | *IAV:* Gamma(mean = 12.2, sd = 7.6, max = 30) | *IAV:* Fecal shedding load distribution taken from Nadeau et al. (2024)[30] based on fecal shedding load distributions from Hirose et al. (2016)[31] and Chan et al. (2011).[32] |
| | | *IBV:* See IAV | *IBV*: Given the paucity of longitudinal fecal shedding concentration data for IBV RNA,[33] we assumed the same distribution as IAV. |
| | | *RSV:* See SARS-CoV-2 | *RSV:* Longitudinal fecal shedding concentration data are not available for RSV RNA.[33] Given the generation time and incubation period of RSV are more similar to SARS-CoV-2 than those of IAV/IBV, we assumed a SARS-CoV-2-like fecal shedding load distribution. |
| | | *NoV:* NonParametric(pmf = 0.000000e+00, 1.634193e-03, 2.664704e-02, 1.026991e-01, 1.863745e-01, 2.133237e-01, 1.807872e-01, 1.253305e-01, 7.596084e-02, 4.215972e-02, 2.215130e-02, 1.128742e-02, 5.678045e-03, 2.856885e-03, 1.451593e-03, 7.500333e-04, 3.960543e-04, 2.144628e-04, 1.193548e-04, 6.835723e-05, 4.031255e-05, 2.448044e-05, 1.530191e-05, 9.837812e-06, 6.498986e-06, 4.406282e-06, 3.061991e-06, 2.177839e-06, 1.583068e-06, 1.174297e-06, | *NoV:* We derived a normalized fecal shedding load pmf from a systematic review and meta-analysis by Zheng et al. (2025).[34] Specifically, we used the model parameters provided for NoV RNA concentration in stool measured days after infection to predict concentrations (cp/g-stool) on days 0–30; we then normalized the pmf. |

| Parameter | Definition | Distribution | Source |
|---|---|---|---|
| | | 8.875974e-07)<br><br>*Rotavirus:* NonParametric(pmf = 0.000000e+00, 7.626647e-02, 1.471949e-01, 1.687350e-01, 1.547742e-01, 1.259491e-01, 9.551032e-02, 6.937532e-02, 4.909180e-02, 3.421980e-02, 2.367562e-02, 1.634539e-02, 1.130364e-02, 7.851874e-03, 5.489519e-03, 3.868433e-03, 2.750610e-03, 1.974843e-03, 1.432374e-03, 1.049856e-03, 7.777091e-04, 5.822808e-04, 4.406054e-04, 3.369081e-04, 2.602763e-04, 2.031033e-04, 1.600444e-04, 1.273138e-04, 1.022077e-04, 8.277889e-05, 6.761366e-05) | *Rotavirus:* We derived a normalized fecal shedding load pmf from a systematic review and meta-analysis by Zheng et al. (2025). Specifically, we used the model parameters provided for rotavirus RNA concentration in stool measured days after infection to predict concentrations (cp/g-stool) on days 0–30; we then normalized the pmf. |
| Initial reproduction number | Initial belief about the reproduction number before data are observed. | LogNormal(mean = 1, sd = 1) | We used the EpiNow2 default as we did not have prior information about Rt across pathogens, sites, and datasets. |
| Observation scale | *Clinical data:* Underascertainment rate, or the scaling factor mapping true latent infections to observed clinical counts. E.g., not everyone who is infected seeks care.<br><br>*Wastewater data:* Scaling factor mapping latent infections to the observed wastewater signal. E.g., not everyone who is infected sheds viral nucleic acids. | Fixed(1) | We used the EpiNow2 default of complete ascertainment or complete wastewater detection because we only aimed to estimate $R_t$ (relative changes in the surveillance signal) rather than actual counts. |
| Week effect | Adjustment factor accounting for periodic reporting fluctuations. | FALSE | The data used in this study were not subject to periodic reporting fluctuations. Clinical data were reported on a weekly basis, and wastewater monitoring data are independent of test-seeking and test-reporting behaviors. |

| Parameter | Definition | Distribution | Source |
|---|---|---|---|
| Dispersion | Overdispersion parameter of the negative binomial observation model. | Normal(mean = 0, sd = 1) | We used a larger standard deviation than the EpiNow2 default to allow for more overdispersion. Wastewater monitoring data, in particular, are prone to high within- and between-day variability.[35] |
| Gaussian process length scale (days) | Controls the smoothness of $R_t$ estimates over time. | LogNormal(mean = 21, sd = 7, max = 60) | We used the EpiNow2 default. No rapid policy changes occurred during our analysis period (e.g., lockdowns). |
| Gaussian process magnitude | Controls the amplitude of variations in log($R_t$) over time. | Normal(mean = 0, sd = 0.01) | We used the EpiNow2 default which works well for the renewal equation model in most scenarios. |
| Truncation | Allows for right-trunction of recent data due to reporting delays whereby recent observations are revised upward. | Fixed(0) | We used the EpiNow2 default (no truncation) as we conducted the analysis in a retrospective manner; the most recent observations were fully observed. |

For further details about the various model parameters, refer to the EpiNow2 R documentation.[36]

Abbreviations: CI = confidence interval, cp/mg = copies per milligram, IAV/IBV = influenza A/B virus, NoV = norovirus, pmf = probability mass function, PMMoV = pepper mild mottle virus, RSV = respiratory syncytial virus, sd = standard deviation.

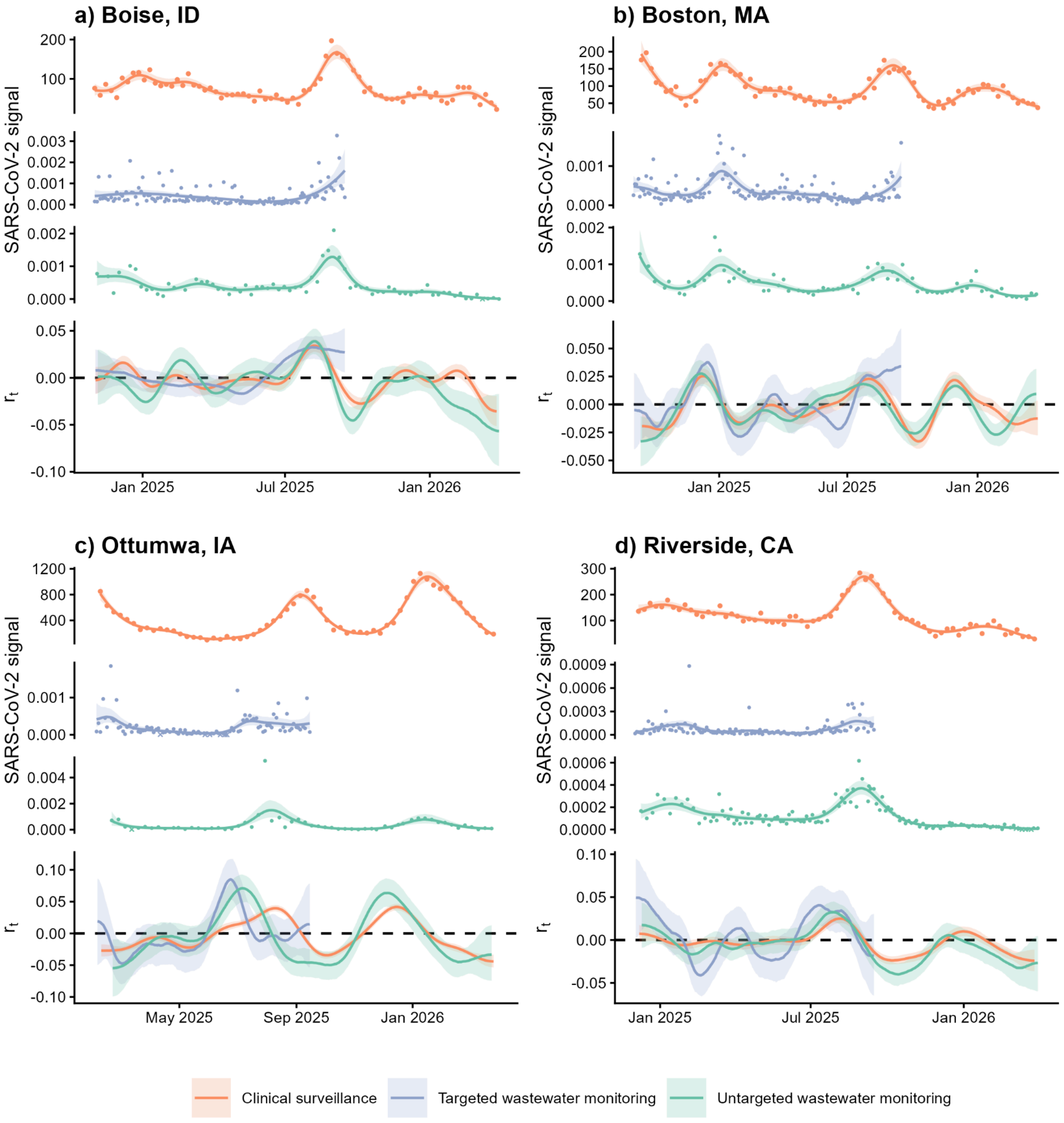

**Fig. S1. SARS-CoV-2 surveillance data and $r_t$ estimates for all locations.**
Time series data from each data source with GAM smoothing alongside $r_t$ estimates are shown for (a) Boise, Idaho; (b) Boston, Massachusetts; (c) Ottumwa, Iowa; and (d) Riverside, California. "Signal" indicates counts of COVID-19 diagnoses from Epic Cosmos for clinical data (orange), PMMoV-normalized concentrations of SARS-CoV-2 RNA for targeted wastewater data (purple), and PMMoV-normalized SARS-CoV-2 clade counts scaled to account for sequencing depth for untargeted wastewater data (green). For wastewater monitoring data sources, x's denote non-detect samples; outlier measurements are not shown for visualization. Lines represent median estimates with 90% CrIs shaded. Abbreviations: CrI = credible interval, GAM = generalized additive model, PMMoV = pepper mild mottle virus.

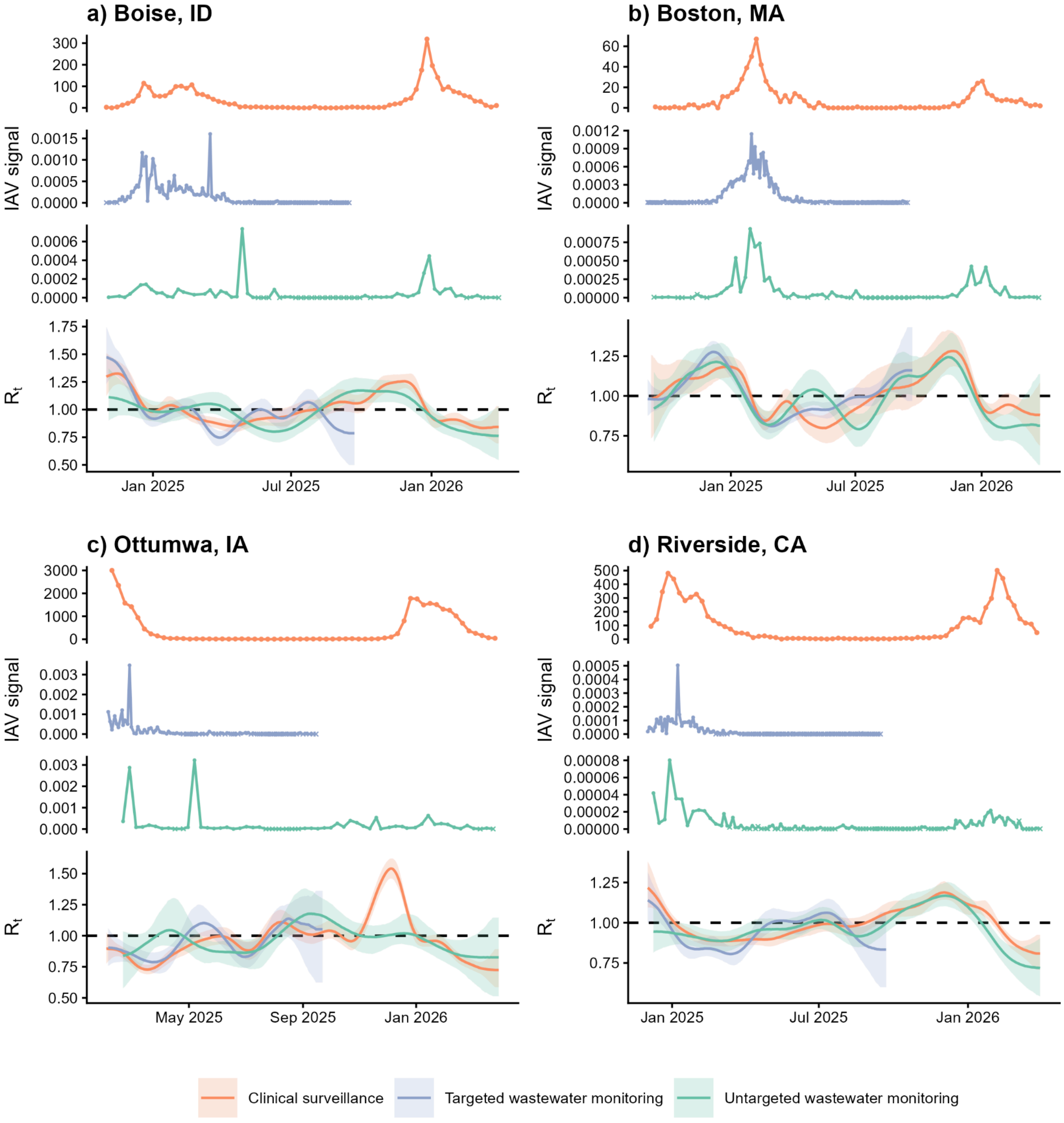

**Fig. S2. IAV surveillance data and $R_t$ estimates for all locations.**

Time series data from each data source alongside $R_t$ estimates are shown for (**a**) Boise, Idaho; (**b**) Boston, Massachusetts; (**c**) Ottumwa, Iowa; and (**d**) Riverside, California. "Signal" indicates counts of positive IAV lab results from Epic Cosmos for clinical data (orange), PMMoV-normalized concentrations of IAV RNA for targeted wastewater data (purple), and PMMoV-normalized IAV clade counts scaled to account for sequencing depth for untargeted wastewater data (green). For wastewater monitoring data sources, x's denote non-detect samples; outlier measurements are not shown for visualization. On the $R_t$ panels, lines represent median estimates with 90% CrIs shaded. Abbreviations: CrI = credible interval, IAV = influenza A virus, PMMoV = pepper mild mottle virus.

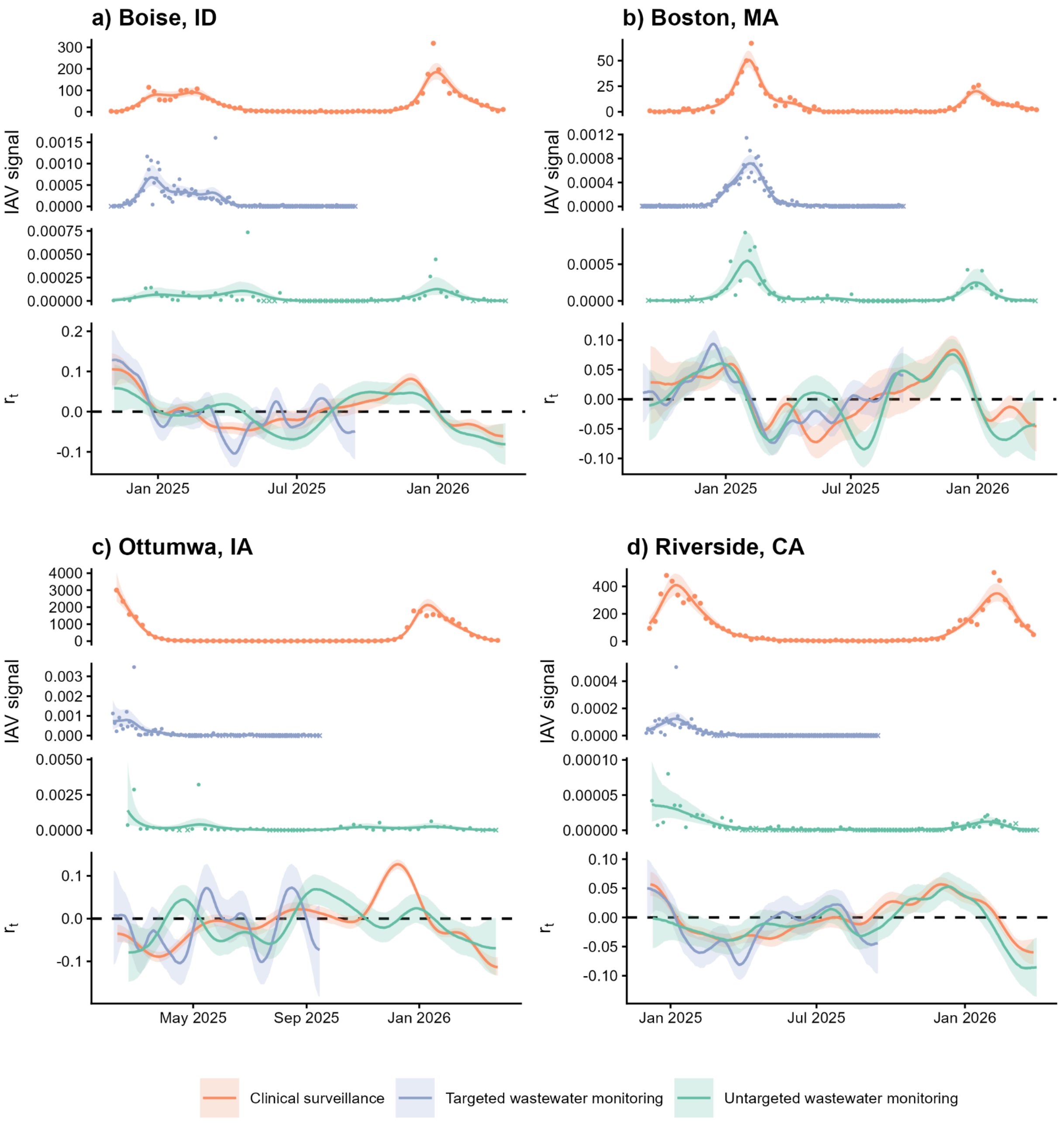

**Fig. S3. IAV surveillance data and $r_t$ estimates for all locations.**

Time series data from each data source with GAM smoothing alongside $r_t$ estimates are shown for (a) Boise, Idaho; (b) Boston, Massachusetts; (c) Ottumwa, Iowa; and (d) Riverside, California. "Signal" indicates counts of positive IAV lab results from Epic Cosmos for clinical data (orange), PMMoV-normalized concentrations of IAV RNA for targeted wastewater data (purple), and PMMoV-normalized IAV clade counts scaled to account for sequencing depth for untargeted wastewater data (green). For wastewater monitoring data sources, x's denote non-detect samples; outlier measurements are not shown for visualization. Lines represent median estimates with 90% CrIs shaded. Abbreviations: CrI = credible interval, GAM = generalized additive model, IAV = influenza A virus, PMMoV = pepper mild mottle virus.

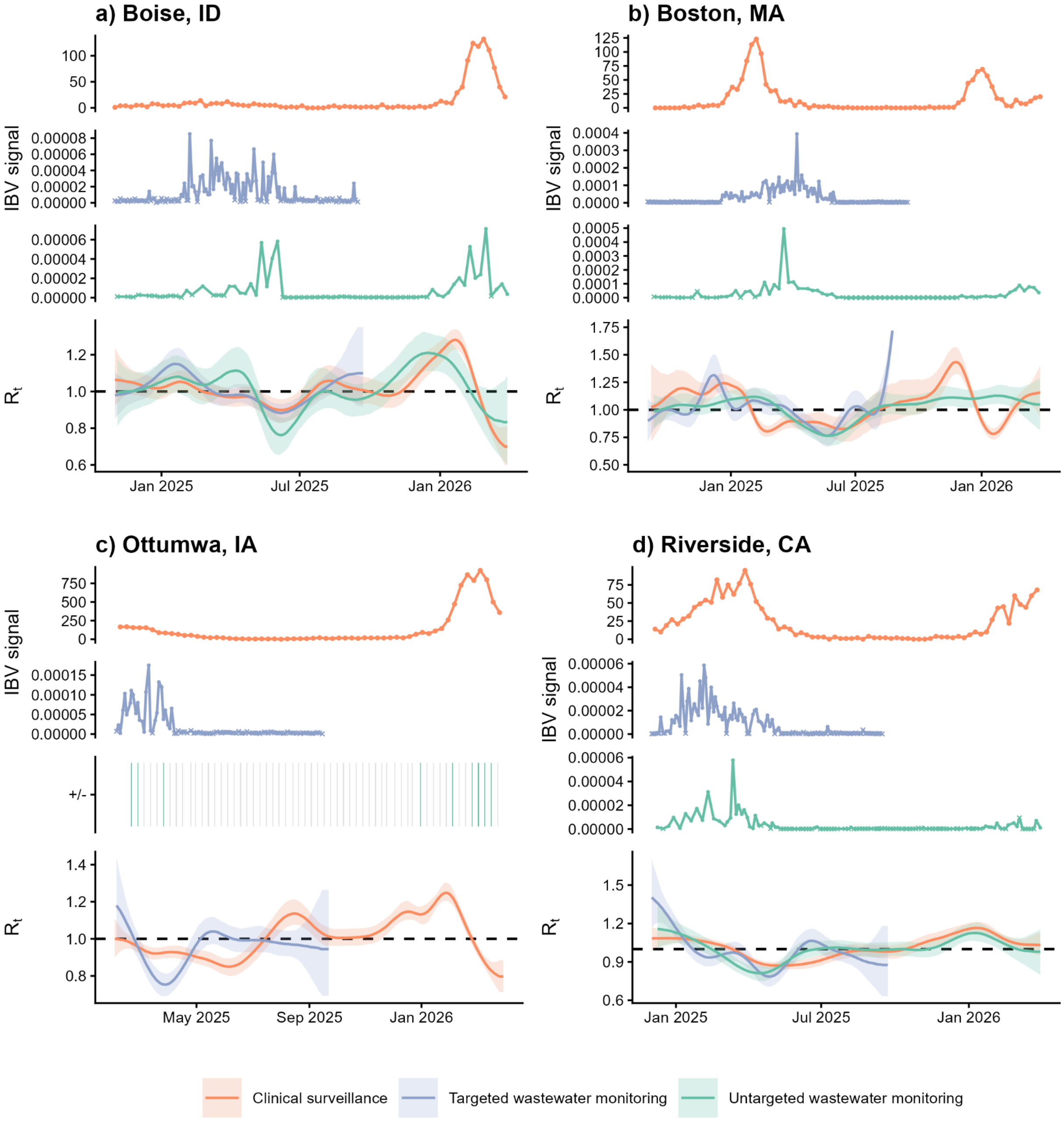

**Fig. S4. IBV surveillance data and $R_t$ estimates for all locations.**

Time series data from each data source alongside $R_t$ estimates are shown for (**a**) Boise, Idaho; (**b**) Boston, Massachusetts; (**c**) Ottumwa, Iowa; and (**d**) Riverside, California. “Signal” indicates counts of positive IBV lab results from Epic Cosmos for clinical data (orange), PMMoV-normalized concentrations of IBV RNA for targeted wastewater data (purple), and PMMoV-normalized IBV clade counts scaled to account for sequencing depth for untargeted wastewater data (green). For wastewater monitoring data sources, x’s denote non-detect samples; outlier measurements are not shown for visualization. Heatmaps show virus presence/absence when the virus was not detected in >75% of wastewater samples. On the $R_t$ panels, lines represent median estimates with 90% CrIs shaded. The $R_t$ y-axis in panel **b** was adjusted such that extreme estimates are not displayed for visualization purposes only; refer to **Fig. S5** for this figure without adjustment. Abbreviations: CrI = credible interval, IBV = influenza B virus, PMMoV = pepper mild mottle virus.

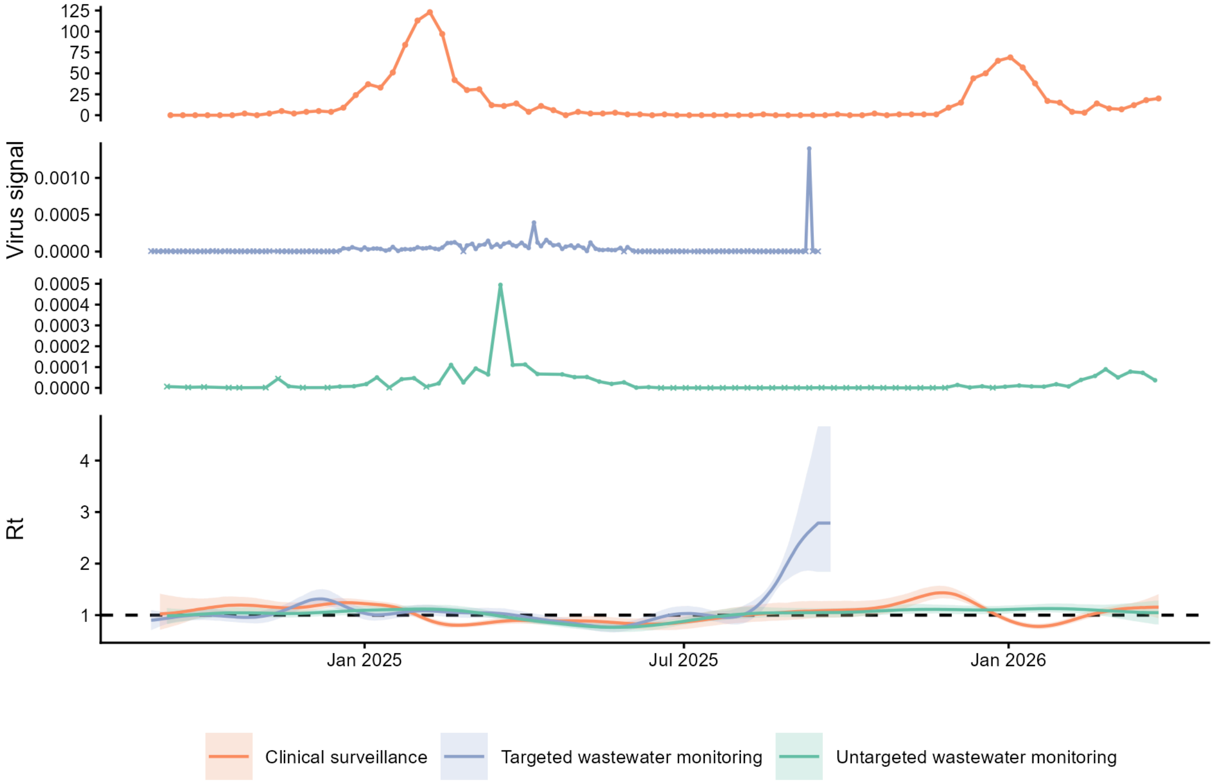

**Fig. S5. IBV surveillance data and $R_t$ estimates for Boston, Massachusetts without y-axis adjustment.**

Time series data from each data source alongside $R_t$ estimates are shown for Boston, Massachusetts. Virus "signal" indicates counts of positive IBV lab results from Epic Cosmos for clinical data (orange), PMMoV-normalized concentrations of IBV RNA for targeted wastewater data (purple), and PMMoV-normalized IBV clade counts scaled to account for sequencing depth for untargeted wastewater data (green). For wastewater monitoring data sources, x's denote non-detect samples; outlier measurements are shown. On the $R_t$ panel, lines represent median estimates with 90% CrIs shaded. Abbreviations: CrI = credible interval, IBV = influenza B virus, PMMoV = pepper mild mottle virus.

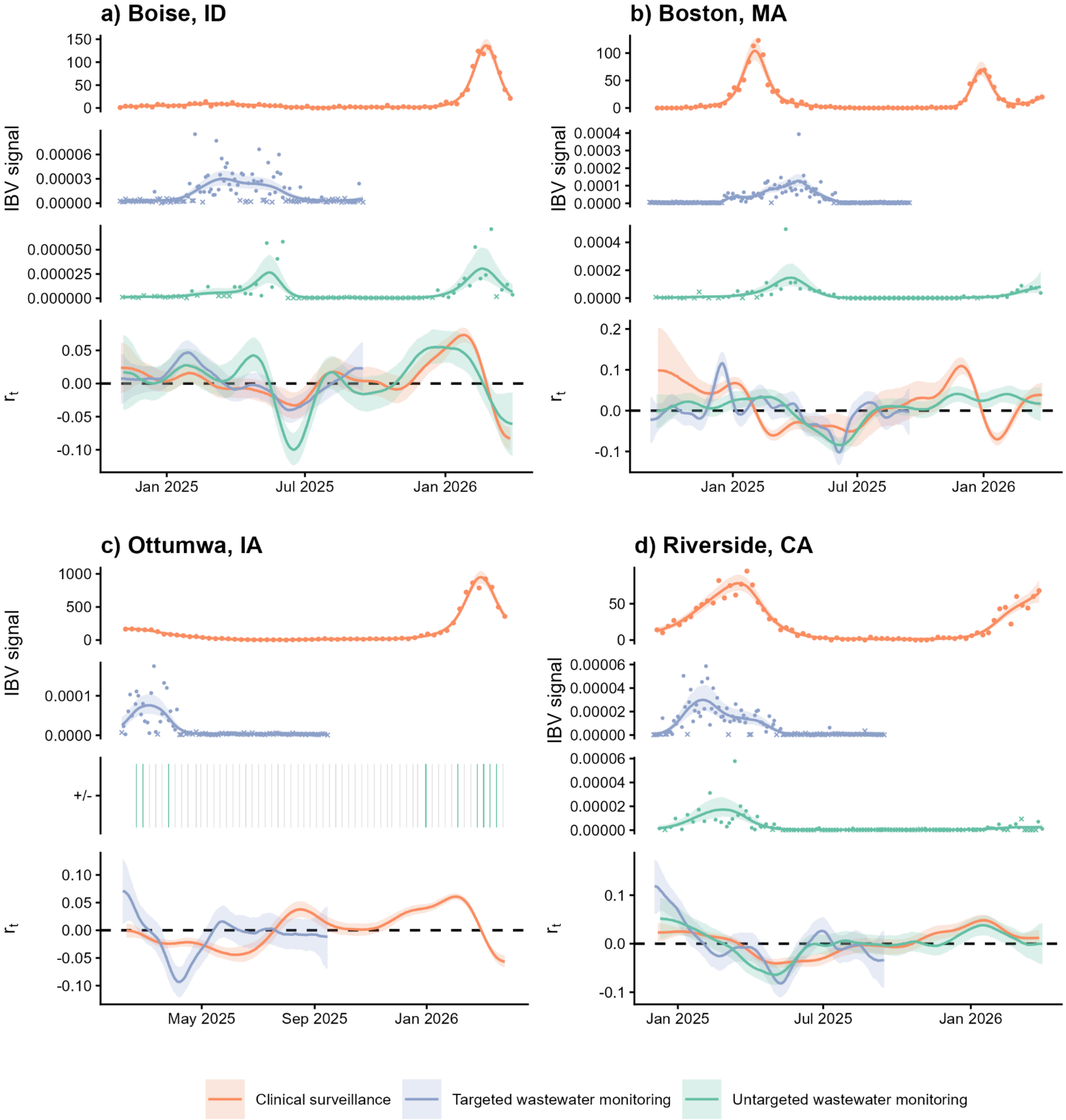

**Fig. S6. IBV surveillance data and $r_t$ estimates for all locations.**

Time series data from each data source with GAM smoothing alongside $r_t$ estimates are shown for (a) Boise, Idaho; (b) Boston, Massachusetts; (c) Ottumwa, Iowa; and (d) Riverside, California. "Signal" indicates counts of positive IBV lab results from Epic Cosmos for clinical data (orange), PMMoV-normalized concentrations of IBV RNA for targeted wastewater data (purple), and PMMoV IBV clade counts scaled to account for sequencing depth for untargeted wastewater data (green). For wastewater monitoring data sources, x's denote non-detect samples; outlier measurements are not shown for visualization. Heatmaps show presence/absence when the virus was not detected in >75% of wastewater samples. Lines represent median estimates with 90% CrIs shaded. Abbreviations: CrI = credible interval, GAM = generalized additive model, IBV = influenza B virus, PMMoV = pepper mild mottle virus.

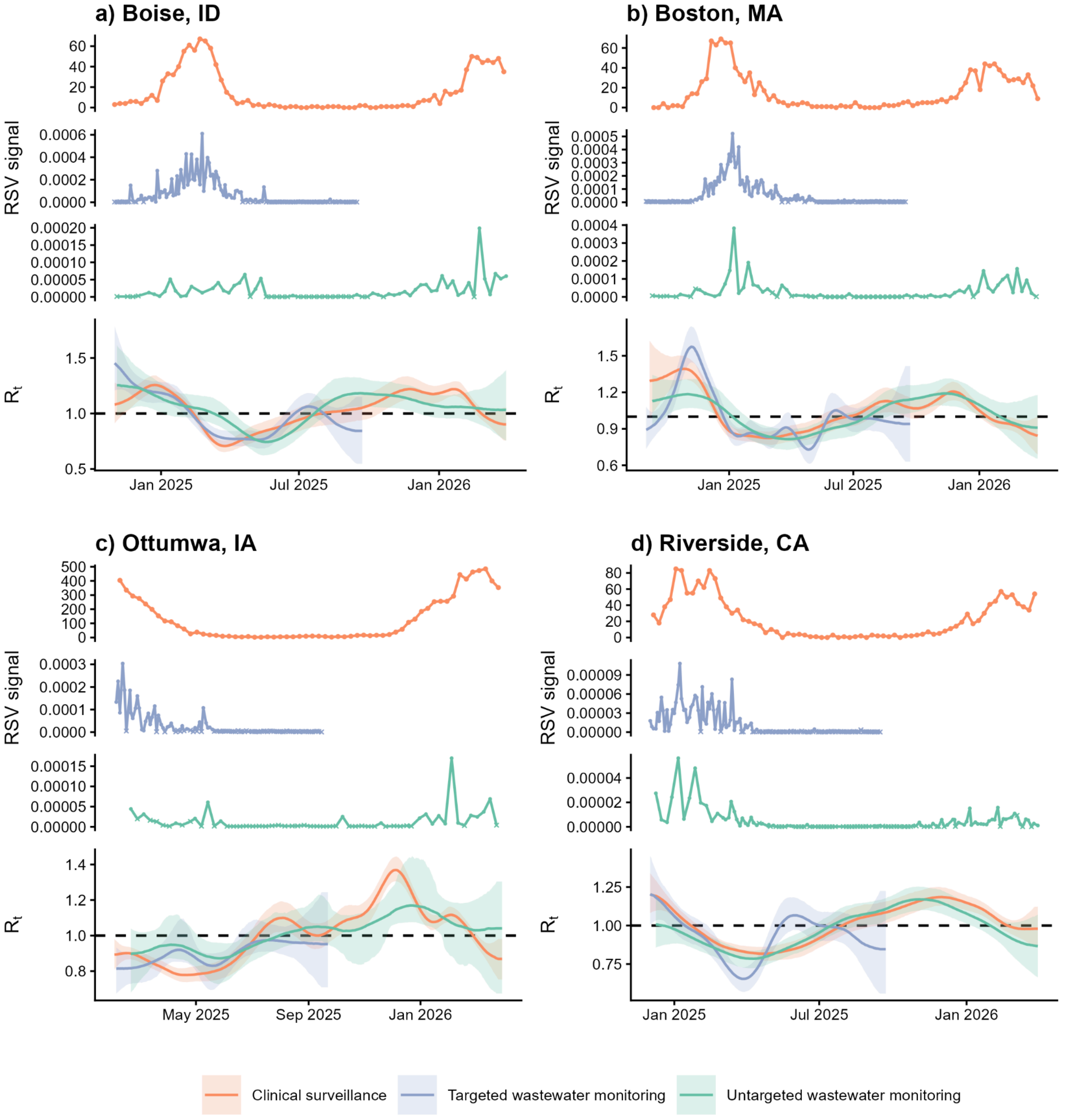

**Fig. S7. RSV surveillance data and $R_t$ estimates for all locations.**
Time series data from each data source alongside $R_t$ estimates are shown for (**a**) Boise, Idaho; (**b**) Boston, Massachusetts; (**c**) Ottumwa, Iowa; and (**d**) Riverside, California. "Signal" indicates counts of RSV diagnoses from Epic Cosmos for clinical data (orange), PMMoV-normalized concentrations of RSV RNA for targeted wastewater data (purple), and PMMoV-normalized RSV clade counts scaled to account for sequencing depth for untargeted wastewater data (green). For wastewater monitoring data sources, x's denote non-detect samples; outlier measurements are not shown for visualization. On the $R_t$ panels, lines represent median estimates with 90% CrIs shaded. Abbreviations: CrI = credible interval, PMMoV = pepper mild mottle virus, RSV = respiratory syncytial virus.

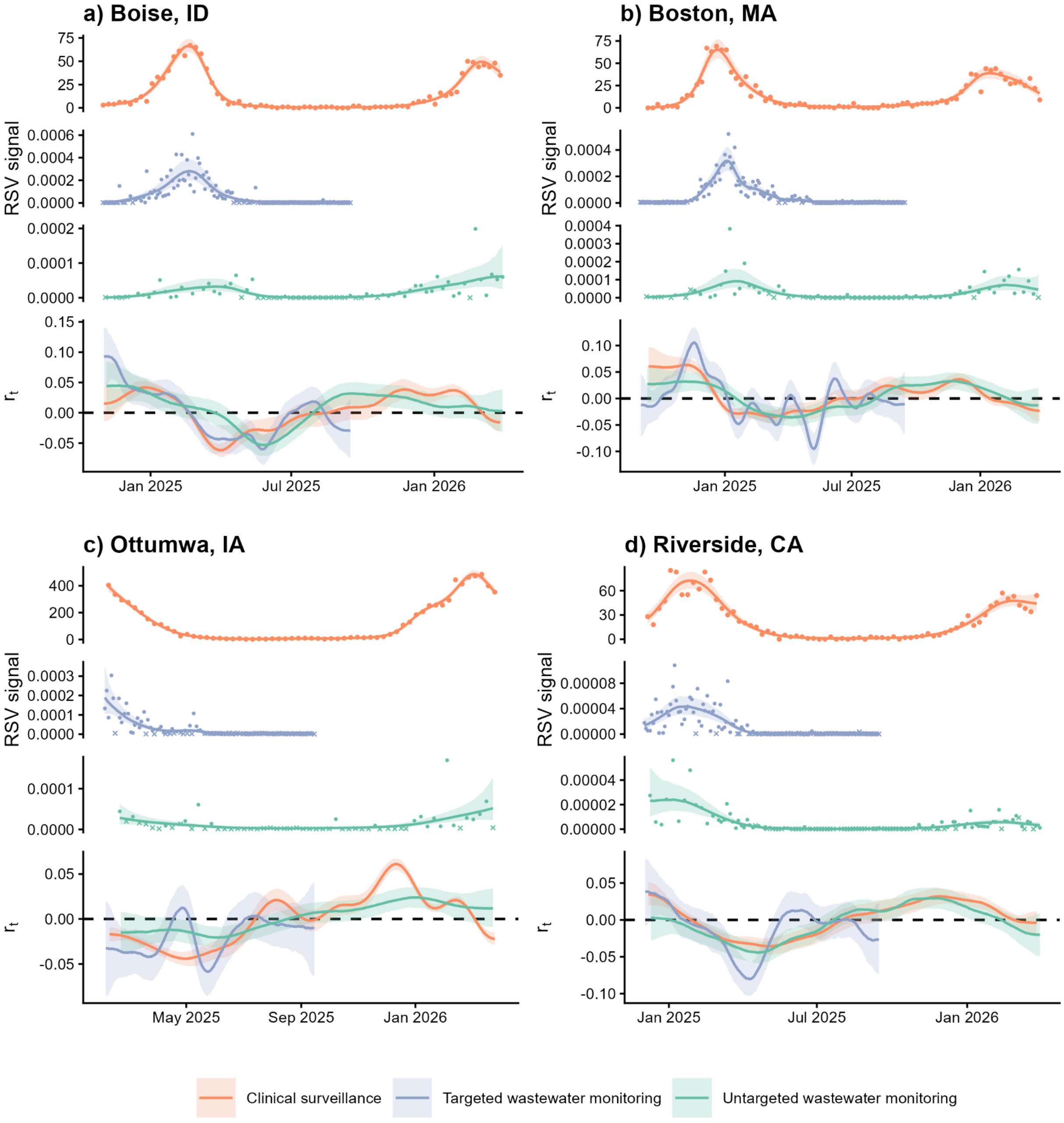

**Fig. S8. RSV surveillance data and $r_t$ estimates for all locations.**

Time series data from each data source with GAM smoothing alongside $r_t$ estimates are shown for (a) Boise, Idaho; (b) Boston, Massachusetts; (c) Ottumwa, Iowa; and (d) Riverside, California. "Signal" indicates counts of RSV diagnoses from Epic Cosmos for clinical data (orange), PMMoV-normalized concentrations of RSV RNA for targeted wastewater data (purple), and PMMoV-normalized RSV clade counts scaled to account for sequencing depth for untargeted wastewater data (green). For wastewater monitoring data sources, x's denote non-detect samples; outlier measurements are not shown for visualization. Lines represent median estimates with 90% CrIs shaded. Abbreviations: CrI = credible interval, GAM = generalized additive model, PMMoV = pepper mild mottle virus, RSV = respiratory syncytial virus.

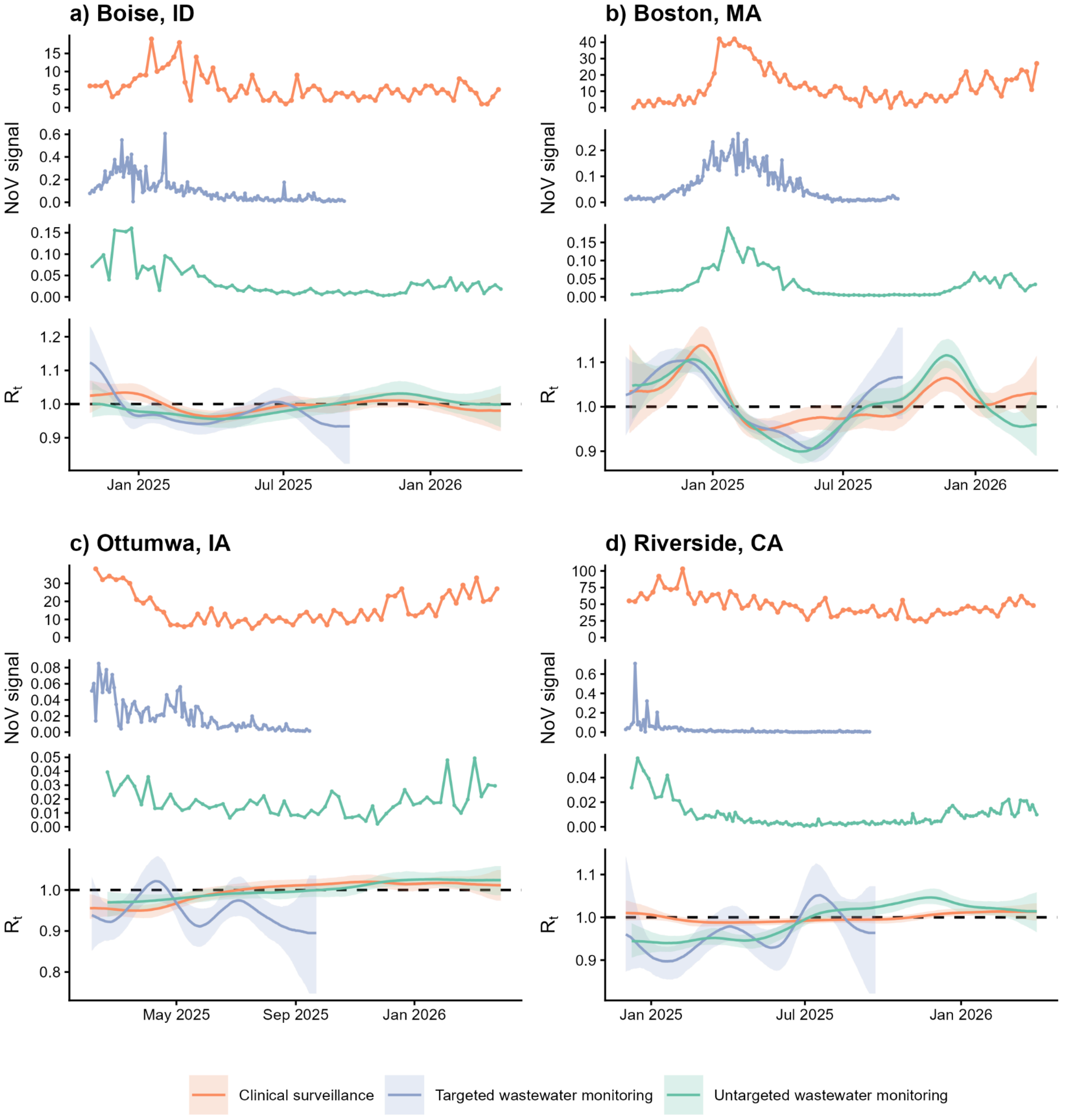

**Fig. S9. NoV surveillance data and $R_t$ estimates for all locations.**
Time series data from each data source alongside $R_t$ estimates are shown for (**a**) Boise, Idaho; (**b**) Boston, Massachusetts; (**c**) Ottumwa, Iowa; and (**d**) Riverside, California. "Signal" indicates counts of NoV diagnoses from Epic Cosmos for clinical data (orange), PMMoV-normalized concentrations of NoV GII RNA for targeted wastewater data (purple), and PMMoV-normalized NoV GII clade counts scaled to account for sequencing depth for untargeted wastewater data (green). For wastewater monitoring data sources, x's denote non-detect samples; outlier measurements are not shown for visualization. On the $R_t$ panels, lines represent median estimates with 90% CrIs shaded. Abbreviations: CrI = credible interval, NoV = norovirus, PMMoV = pepper mild mottle virus.

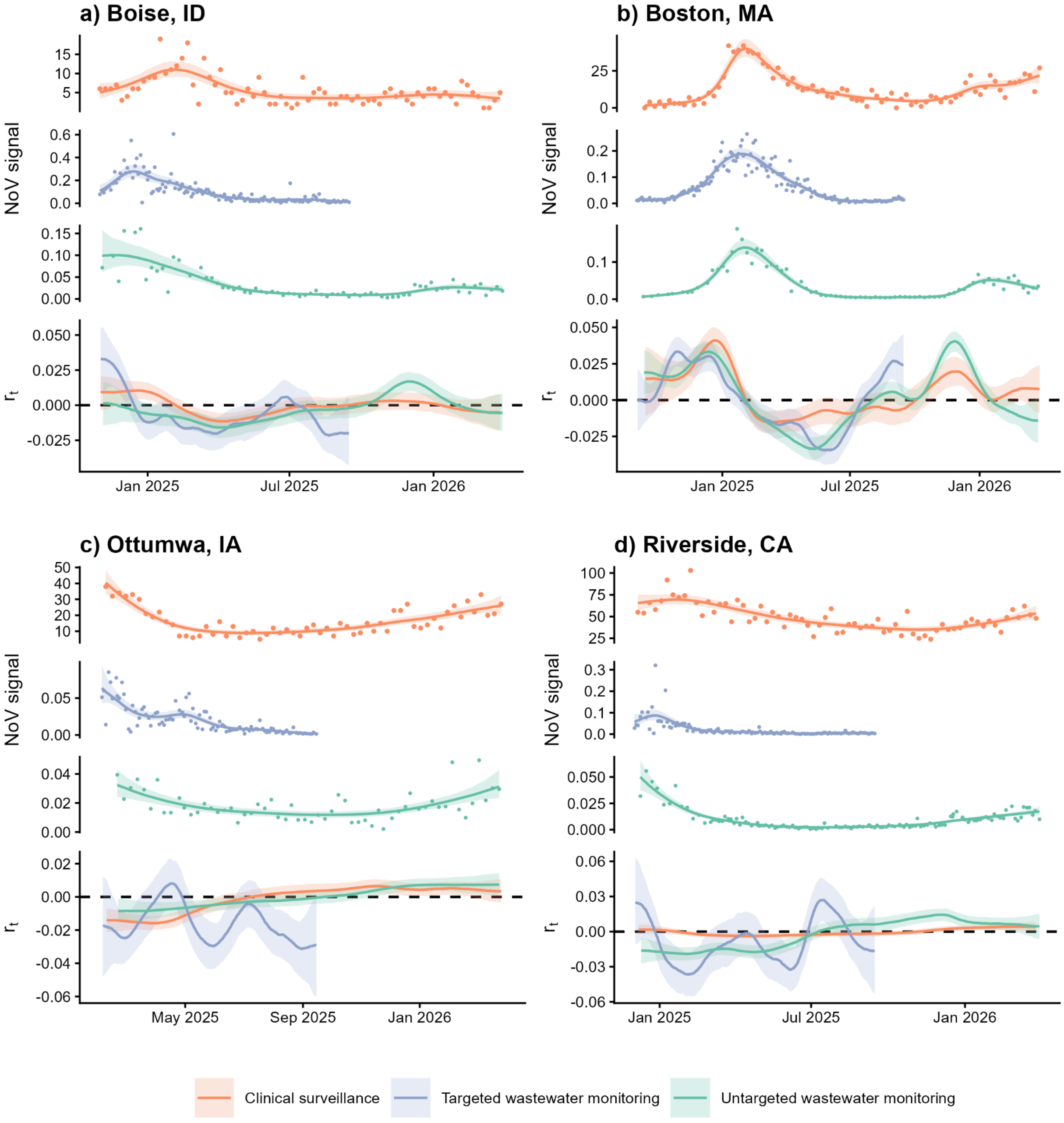

**Fig. S10. NoV surveillance data and $r_t$ estimates for all locations.**

Time series data from each data source with GAM smoothing alongside $r_t$ estimates are shown for (a) Boise, Idaho; (b) Boston, Massachusetts; (c) Ottumwa, Iowa; and (d) Riverside, California. “Signal” indicates counts of NoV diagnoses from Epic Cosmos for clinical data (orange), PMMoV-normalized concentrations of NoV GII RNA for targeted wastewater data (purple), and PMMoV-normalized NoV GII clade counts scaled to account for sequencing depth for untargeted wastewater data (green). For wastewater monitoring data sources, x’s denote non-detect samples; outlier measurements are not shown for visualization. Lines represent median estimates with 90% CrIs shaded. Abbreviations: CrI = credible interval, GAM = generalized additive model, NoV = norovirus, PMMoV = pepper mild mottle virus.

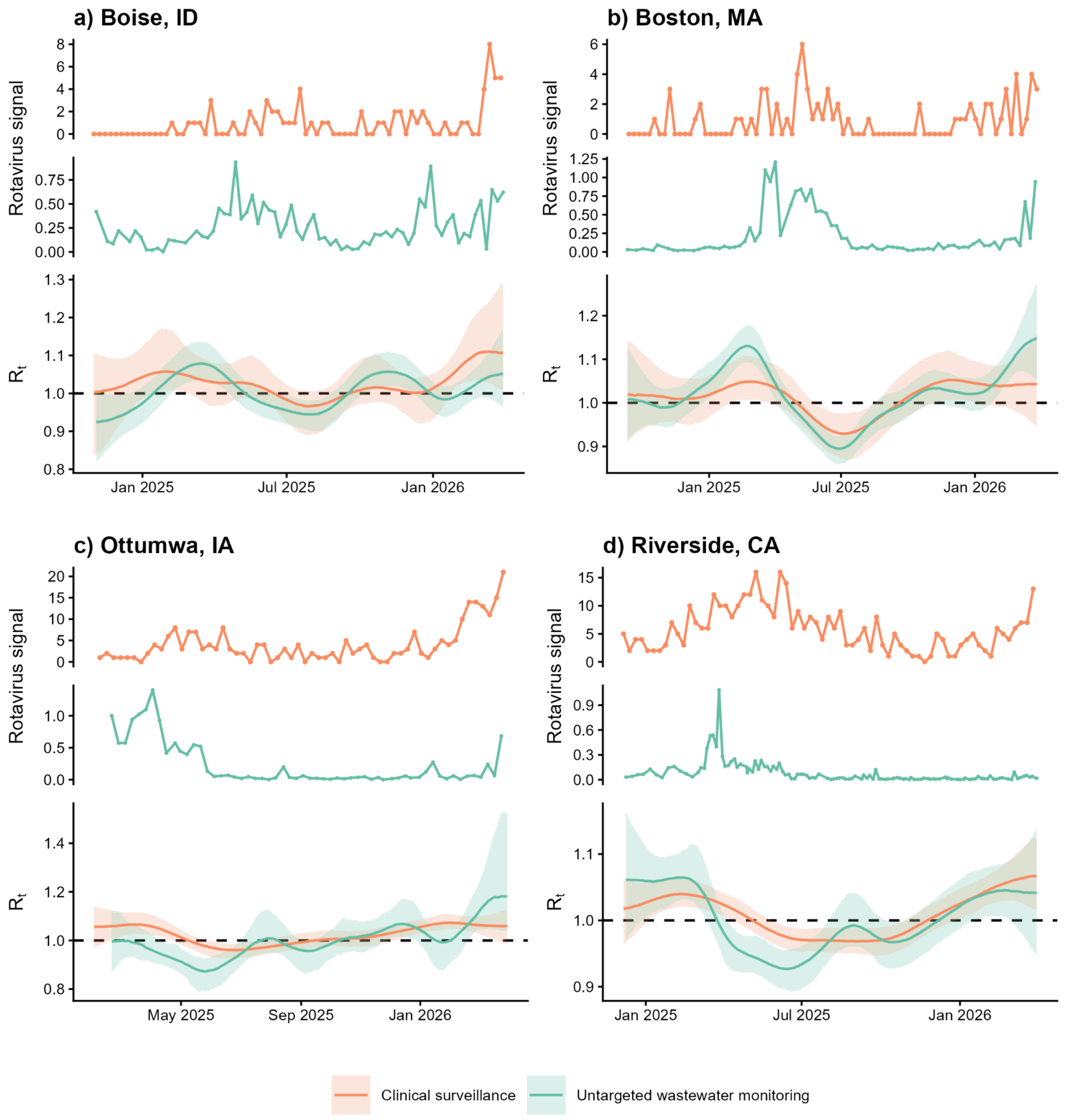

**Fig. S11. Rotavirus surveillance data and $R_t$ estimates for all locations.**
Time series data from each data source alongside $R_t$ estimates are shown for (**a**) Boise, Idaho; (**b**) Boston, Massachusetts; (**c**) Ottumwa, Iowa; and (**d**) Riverside, California. "Signal" indicates counts of rotavirus diagnoses from Epic Cosmos for clinical data (orange) and PMMoV-normalized rotavirus clade counts scaled to account for sequencing depth for untargeted wastewater data (green). For wastewater monitoring data sources, x's denote non-detect samples; outlier measurements are not shown for visualization. On the $R_t$ panels, lines represent median estimates with 90% CrIs shaded. Abbreviations: CrI = credible interval, NoV = norovirus, PMMoV = pepper mild mottle virus.

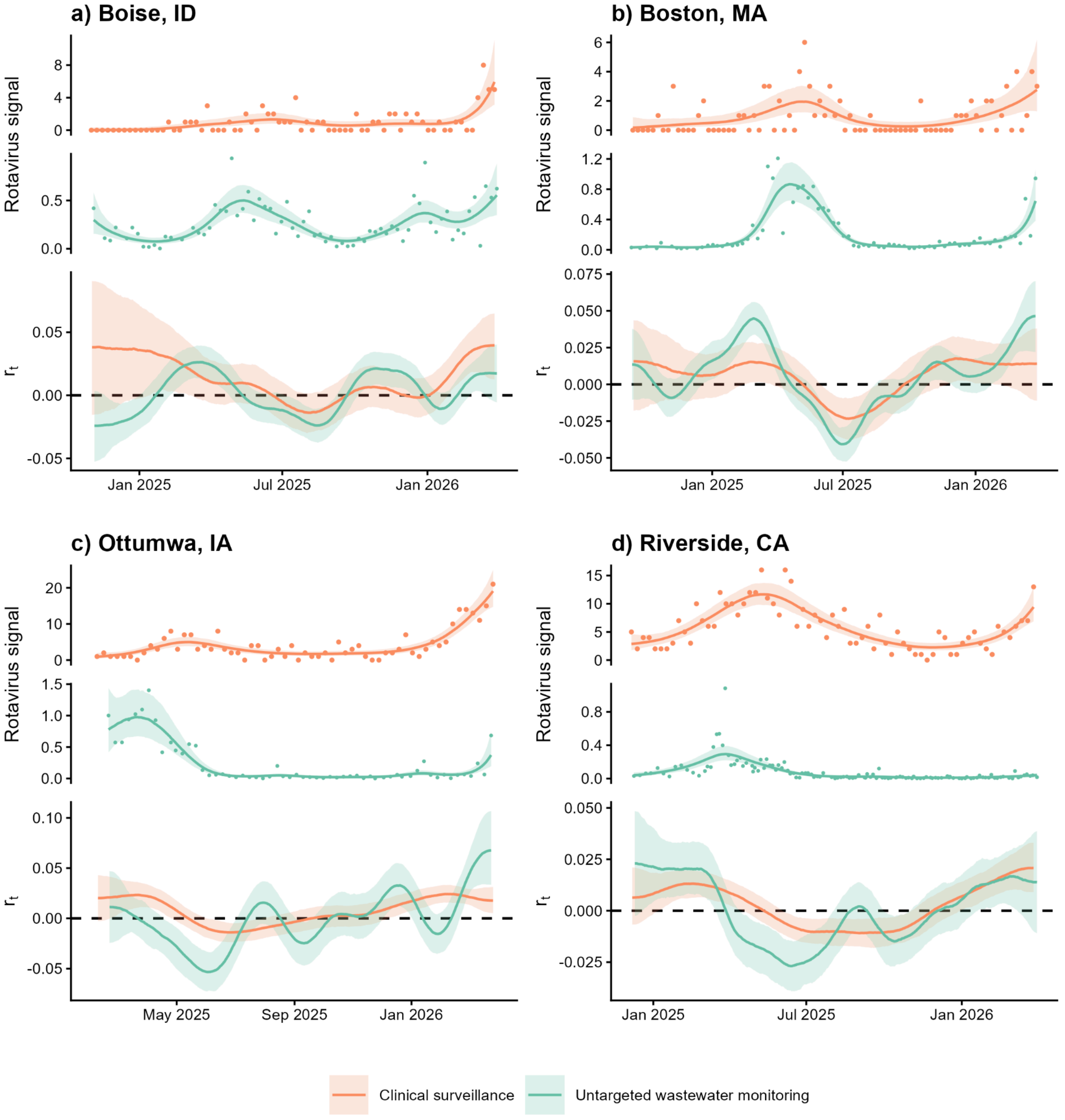

**Fig. S12. Rotavirus surveillance data and $r_t$ estimates for all locations.**
Time series data from each data source with GAM smoothing alongside $r_t$ estimates are shown for (a) Boise, Idaho; (b) Boston, Massachusetts; (c) Ottumwa, Iowa; and (d) Riverside, California. "Signal" indicates counts of rotavirus diagnoses from Epic Cosmos for clinical data (orange) and PMMoV-normalized rotavirus clade counts scaled to account for sequencing depth for untargeted wastewater data (green). For wastewater monitoring data sources, x's denote non-detect samples; outlier measurements are not shown for visualization. Lines represent median estimates with 90% CrIs shaded. Abbreviations: CrI = credible interval, GAM = generalized additive model, PMMoV = pepper mild mottle virus.

**Table S5. Observed posterior distributions for metrics of concordance among derived $R_t$ estimates**

| Virus | Shedding distribution | Location | Comparison | MAE (95% CrI) | MAE permutation *p* value | AOB1 (95% CrI) | AOB1 permutation *p* value | Tau (95% CrI) | Tau permutation *p* value |
|---|---|---|---|---|---|---|---|---|---|
| SARS-CoV-2 | Main analysis | Boston, MA | Clinical vs targeted wastewater | 0.0527 (0.0351, 0.0764) | 0.0005 | 0.7218 (0.5906, 0.8478) | 0.0015 | 0.3275 (0.1555, 0.5227) | < 0.0001 |
| | | | Clinical vs untargeted wastewater | 0.0420 (0.0292, 0.0592) | < 0.0001 | 0.7482 (0.6514, 0.8556) | < 0.0001 | 0.4353 (0.2722, 0.5933) | < 0.0001 |
| | | | Targeted vs untargeted wastewater | 0.0671 (0.0448, 0.0931) | 0.026 | 0.6340 (0.5066, 0.7586) | 0.027 | 0.2088 (0.0130, 0.4294) | 0.029 |
| | | Boise, ID | Clinical vs targeted wastewater | 0.0462 (0.0322, 0.0658) | 0.001 | 0.6503 (0.4939, 0.8129) | 0.044 | 0.2578 (0.0389, 0.4847) | 0.016 |
| | | | Clinical vs untargeted wastewater | 0.0532 (0.0387, 0.0735) | 0.0005 | 0.6172 (0.5019, 0.7365) | 0.077 | 0.3055 (0.1251, 0.4711) | 0.001 |
| | | | Targeted vs untargeted wastewater | 0.0620 (0.0402, 0.0911) | 0.0035 | 0.5573 (0.3901, 0.7461) | 0.2725 | 0.0982 (-0.1390, 0.4094) | 0.234 |
| | | Ottumwa, IA | Clinical vs targeted wastewater | 0.0977 (0.0679, 0.1569) | 0.064 | 0.6767 (0.5388, 0.8190) | 0.0225 | 0.2302 (0.0066, 0.4623) | 0.0335 |
| | | | Clinical vs untargeted wastewater | 0.0936 (0.0728, 0.1250) | < 0.0001 | 0.7666 (0.6535, 0.8526) | < 0.0001 | 0.4341 (0.2237, 0.5780) | < 0.0001 |
| | | | Targeted vs untargeted wastewater | 0.1001 (0.0499, 0.1769) | 0.0135 | 0.7176 (0.4954, 0.9491) | 0.0355 | 0.3055 (-0.0337, 0.7176) | 0.0495 |
| | | Riverside, CA | Clinical vs targeted wastewater | 0.0689 (0.0387, 0.1086) | 0.0025 | 0.6898 (0.5051, 0.8610) | 0.0225 | 0.3670 (0.0855, 0.6083) | 0.005 |
| | | | Clinical vs untargeted wastewater | 0.0352 (0.0264, 0.0477) | < 0.0001 | 0.7568 (0.6445, 0.8649) | < 0.0001 | 0.5646 (0.3800, 0.7078) | < 0.0001 |
| | | | Targeted vs untargeted wastewater | 0.0594 (0.0316, 0.0985) | < 0.0001 | 0.7222 (0.5382, 0.8854) | 0.007 | 0.4497 (0.1203, 0.7110) | 0.0045 |
| | Alternative A | Boston, MA | Clinical vs targeted wastewater | 0.0538 (0.0363, 0.0773) | 0.0005 | 0.7165 (0.5879, 0.8346) | 0.0005 | 0.3211 (0.1539, 0.5152) | 0.0005 |
| | | | Clinical vs untargeted wastewater | 0.0443 (0.0294, 0.0611) | < 0.0001 | 0.7430 (0.5968, 0.8539) | < 0.0001 | 0.4218 (0.1258, 0.6045) | < 0.0001 |
| | | | Targeted vs untargeted wastewater | 0.0686 (0.0468, 0.0959) | 0.0995 | 0.6207 (0.4589, 0.7587) | 0.0965 | 0.1891 (-0.0438, 0.4189) | 0.08 |
| | | Boise, ID | Clinical vs targeted | 0.0466 (0.0319, | < 0.0001 | 0.6442 (0.4816, | 0.0635 | 0.2575 (0.0319, | 0.0175 |

| Virus | Shedding distribution | Location | Comparison | MAE (95% CrI) | MAE permutation *p* value | AOB1 (95% CrI) | AOB1 permutation *p* value | Tau (95% CrI) | Tau permutation *p* value |
|---|---|---|---|---|---|---|---|---|---|
| | | | wastewater | 0.0673) | | 0.8067) | | 0.4840) | |
| | | | Clinical vs untargeted wastewater | 0.0524 (0.0383, 0.0734) | 0.0005 | 0.6163 (0.5039, 0.7306) | 0.074 | 0.3095 (0.1368, 0.4723) | 0.0015 |
| | | | Targeted vs untargeted wastewater | 0.0613 (0.0387, 0.0893) | 0.0025 | 0.5666 (0.3932, 0.7523) | 0.2275 | 0.1127 (-0.1382, 0.4028) | 0.2035 |
| | | Ottumwa, IA | Clinical vs targeted wastewater | 0.0962 (0.0653, 0.1522) | 0.0445 | 0.6897 (0.5603, 0.8362) | 0.013 | 0.2568 (0.0439, 0.4844) | 0.0155 |
| | | | Clinical vs untargeted wastewater | 0.0943 (0.0715, 0.1268) | < 0.0001 | 0.7764 (0.6708, 0.8600) | < 0.0001 | 0.4532 (0.2625, 0.6006) | < 0.0001 |
| | | | Targeted vs untargeted wastewater | 0.1002 (0.0517, 0.1701) | 0.011 | 0.7338 (0.5046, 0.9444) | 0.0325 | 0.3314 (-0.0074, 0.7127) | 0.0305 |
| | | Riverside, CA | Clinical vs targeted wastewater | 0.0697 (0.0384, 0.1120) | 0.002 | 0.6949 (0.5254, 0.8542) | 0.0145 | 0.3653 (0.0884, 0.6268) | 0.0065 |
| | | | Clinical vs untargeted wastewater | 0.0350 (0.0254, 0.0496) | < 0.0001 | 0.7692 (0.6466, 0.8732) | < 0.0001 | 0.5866 (0.4012, 0.7230) | < 0.0001 |
| | | | Targeted vs untargeted wastewater | 0.0612 (0.0339, 0.1028) | < 0.0001 | 0.7153 (0.5207, 0.8854) | 0.0125 | 0.4365 (0.0919, 0.6997) | 0.007 |
| | Alternative B | Boston, MA | Clinical vs targeted wastewater | 0.0540 (0.0374, 0.0750) | 0.0005 | 0.7060 (0.5748, 0.8294) | 0.001 | 0.2987 (0.1239, 0.4784) | 0.001 |
| | | | Clinical vs untargeted wastewater | 0.0379 (0.0260, 0.0536) | < 0.0001 | 0.7799 (0.6813, 0.8768) | < 0.0001 | 0.4878 (0.3226, 0.6451) | < 0.0001 |
| | | | Targeted vs untargeted wastewater | 0.0645 (0.0440, 0.0890) | 0.021 | 0.6419 (0.5066, 0.7640) | 0.021 | 0.2257 (0.0145, 0.4426) | 0.0205 |
| | | Boise, ID | Clinical vs targeted wastewater | 0.0458 (0.0314, 0.0641) | < 0.0001 | 0.6564 (0.4938, 0.8190) | 0.044 | 0.2571 (0.0347, 0.4747) | 0.0155 |
| | | | Clinical vs untargeted wastewater | 0.0503 (0.0367, 0.0700) | < 0.0001 | 0.6279 (0.5174, 0.7384) | 0.049 | 0.3351 (0.1450, 0.4894) | < 0.0001 |
| | | | Targeted vs untargeted wastewater | 0.0602 (0.0363, 0.0888) | 0.003 | 0.5666 (0.3962, 0.7524) | 0.2455 | 0.1182 (-0.1425, 0.4082) | 0.1865 |
| | | Ottumwa, IA | Clinical vs targeted wastewater | 0.0945 (0.0640, 0.1488) | 0.0465 | 0.6940 (0.5517, 0.8405) | 0.015 | 0.2532 (0.0209, 0.4903) | 0.027 |

| Virus | Shedding distribution | Location | Comparison | MAE (95% Crl) | MAE permutation *p* value | AOB1 (95% Crl) | AOB1 permutation *p* value | Tau (95% Crl) | Tau permutation *p* value |
|---|---|---|---|---|---|---|---|---|---|
| | | | Clinical vs untargeted wastewater | 0.0888 (0.0663, 0.1196) | < 0.0001 | 0.7862 (0.6806, 0.8722) | < 0.0001 | 0.4721 (0.2913, 0.6069) | < 0.0001 |
| | | | Targeted vs untargeted wastewater | 0.0969 (0.0492, 0.1655) | 0.006 | 0.7315 (0.4954, 0.9491) | 0.0385 | 0.3225 (0.0088, 0.7231) | 0.0295 |
| | | Riverside, CA | Clinical vs targeted wastewater | 0.0688 (0.0399, 0.1053) | 0.0015 | 0.6915 (0.5219, 0.8508) | 0.0175 | 0.3670 (0.0980, 0.6027) | 0.004 |
| | | | Clinical vs untargeted wastewater | 0.0336 (0.0242, 0.0464) | < 0.0001 | 0.7755 (0.6694, 0.8815) | < 0.0001 | 0.6018 (0.4182, 0.7361) | < 0.0001 |
| | | | Targeted vs untargeted wastewater | 0.0593 (0.0328, 0.0957) | < 0.0001 | 0.7292 (0.5347, 0.8889) | 0.0045 | 0.4515 (0.1348, 0.7092) | 0.003 |
| | Alternative C | Boston, MA | Clinical vs targeted wastewater | 0.0537 (0.0367, 0.0774) | < 0.0001 | 0.7139 (0.5853, 0.8320) | 0.0005 | 0.3178 (0.1450, 0.5016) | < 0.0001 |
| | | | Clinical vs untargeted wastewater | 0.0420 (0.0287, 0.0603) | < 0.0001 | 0.7667 (0.6637, 0.8697) | < 0.0001 | 0.4630 (0.2981, 0.6238) | < 0.0001 |
| | | | Targeted vs untargeted wastewater | 0.0674 (0.0465, 0.0941) | 0.023 | 0.6419 (0.5066, 0.7692) | 0.0265 | 0.2300 (0.0247, 0.4367) | 0.022 |
| | | Boise, ID | Clinical vs targeted wastewater | 0.0465 (0.0318, 0.0677) | < 0.0001 | 0.6472 (0.4755, 0.8067) | 0.0565 | 0.2472 (0.0338, 0.4711) | 0.011 |
| | | | Clinical vs untargeted wastewater | 0.0519 (0.0379, 0.0722) | 0.0005 | 0.6182 (0.5039, 0.7306) | 0.0715 | 0.3136 (0.1268, 0.4730) | 0.0005 |
| | | | Targeted vs untargeted wastewater | 0.0602 (0.0385, 0.0886) | 0.0035 | 0.5728 (0.3932, 0.7523) | 0.2185 | 0.1144 (-0.1453, 0.4156) | 0.2095 |
| | | Ottumwa, IA | Clinical vs targeted wastewater | 0.0963 (0.0671, 0.1561) | 0.0425 | 0.6853 (0.5560, 0.8362) | 0.012 | 0.2593 (0.0286, 0.5021) | 0.022 |
| | | | Clinical vs untargeted wastewater | 0.0932 (0.0704, 0.1259) | < 0.0001 | 0.7813 (0.6732, 0.8649) | < 0.0001 | 0.4602 (0.2776, 0.5987) | < 0.0001 |
| | | | Targeted vs untargeted wastewater | 0.1001 (0.0522, 0.1724) | 0.0135 | 0.7315 (0.4907, 0.9352) | 0.0355 | 0.3346 (-0.0114, 0.7236) | 0.035 |
| | | Riverside, CA | Clinical vs targeted wastewater | 0.0709 (0.0403, 0.1110) | 0.002 | 0.6915 (0.5153, 0.8576) | 0.0245 | 0.3718 (0.0949, 0.6041) | 0.006 |
| | | | Clinical vs untargeted wastewater | 0.0348 (0.0253, 0.0483) | < 0.0001 | 0.7692 (0.6632, 0.8773) | < 0.0001 | 0.5847 (0.4112, 0.7283) | < 0.0001 |

| Virus | Shedding distribution | Location | Comparison | MAE (95% CrI) | MAE permutation *p* value | AOB1 (95% CrI) | AOB1 permutation *p* value | Tau (95% CrI) | Tau permutation *p* value |
|---|---|---|---|---|---|---|---|---|---|
| | | | Targeted vs untargeted wastewater | 0.0624 (0.0346, 0.1016) | < 0.0001 | 0.7222 (0.5312, 0.8750) | 0.009 | 0.4433 (0.1059, 0.7007) | 0.007 |
| Influenza A | Main analysis | Boston, MA | Clinical vs targeted wastewater | 0.0871 (0.0605, 0.1269) | < 0.0001 | 0.8215 (0.6929, 0.9344) | < 0.0001 | 0.5027 (0.3366, 0.6441) | < 0.0001 |
| | | | Clinical vs untargeted wastewater | 0.1049 (0.0715, 0.1481) | < 0.0001 | 0.8011 (0.6637, 0.9085) | < 0.0001 | 0.4432 (0.2619, 0.5926) | < 0.0001 |
| | | | Targeted vs untargeted wastewater | 0.0878 (0.0607, 0.1297) | < 0.0001 | 0.7533 (0.6233, 0.8859) | 0.0005 | 0.4785 (0.3246, 0.6144) | < 0.0001 |
| | | Boise, ID | Clinical vs targeted wastewater | 0.0945 (0.0565, 0.1446) | < 0.0001 | 0.6595 (0.5215, 0.8006) | 0.0375 | 0.3477 (0.1443, 0.5764) | 0.001 |
| | | | Clinical vs untargeted wastewater | 0.1026 (0.0704, 0.1421) | < 0.0001 | 0.7597 (0.6357, 0.8798) | 0.0005 | 0.4543 (0.2625, 0.6188) | < 0.0001 |
| | | | Targeted vs untargeted wastewater | 0.1598 (0.1122, 0.2169) | 0.2285 | 0.5201 (0.3529, 0.6904) | 0.453 | -0.0554 (-0.3310, 0.2467) | 0.652 |
| | | Ottumwa, IA | Clinical vs targeted wastewater | 0.0835 (0.0515, 0.1432) | < 0.0001 | 0.7543 (0.6033, 0.8880) | 0.0225 | 0.5428 (0.2567, 0.7120) | < 0.0001 |
| | | | Clinical vs untargeted wastewater | 0.1422 (0.1051, 0.2096) | 0.0265 | 0.6978 (0.5479, 0.8477) | 0.018 | 0.2984 (0.0425, 0.4989) | 0.014 |
| | | | Targeted vs untargeted wastewater | 0.1307 (0.0737, 0.2370) | 0.231 | 0.6343 (0.4119, 0.8241) | 0.1975 | 0.1583 (-0.3176, 0.5124) | 0.24 |
| | | Riverside, CA | Clinical vs targeted wastewater | 0.0831 (0.0559, 0.1200) | 0.001 | 0.6678 (0.5153, 0.8305) | 0.1465 | 0.2859 (0.0749, 0.5323) | 0.008 |
| | | | Clinical vs untargeted wastewater | 0.0648 (0.0455, 0.0890) | < 0.0001 | 0.7775 (0.6653, 0.8773) | < 0.0001 | 0.5372 (0.3729, 0.6811) | < 0.0001 |
| | | | Targeted vs untargeted wastewater | 0.0727 (0.0446, 0.1165) | < 0.0001 | 0.7326 (0.5763, 0.8889) | 0.0185 | 0.3385 (-0.0044, 0.6359) | 0.032 |
| | Alternative A | Boston, MA | Clinical vs targeted wastewater | 0.0804 (0.0519, 0.1213) | < 0.0001 | 0.8346 (0.7033, 0.9501) | < 0.0001 | 0.5286 (0.3562, 0.6811) | < 0.0001 |
| | | | Clinical vs untargeted wastewater | 0.1012 (0.0680, 0.1413) | < 0.0001 | 0.8028 (0.6813, 0.9173) | < 0.0001 | 0.4282 (0.2689, 0.5875) | < 0.0001 |
| | | | Targeted vs untargeted wastewater | 0.0864 (0.0586, 0.1225) | < 0.0001 | 0.7427 (0.6154, 0.8859) | < 0.0001 | 0.4698 (0.3247, 0.6166) | < 0.0001 |

| Virus | Shedding distribution | Location | Comparison | MAE (95% CrI) | MAE permutation *p* value | AOB1 (95% CrI) | AOB1 permutation *p* value | Tau (95% CrI) | Tau permutation *p* value |
|---|---|---|---|---|---|---|---|---|---|
| | | Boise, ID | Clinical vs targeted wastewater | 0.0875 (0.0559, 0.1255) | < 0.0001 | 0.6626 (0.5275, 0.8068) | 0.0285 | 0.3860 (0.1949, 0.5991) | < 0.0001 |
| | | | Clinical vs untargeted wastewater | 0.0975 (0.0678, 0.1331) | < 0.0001 | 0.7500 (0.6240, 0.8741) | < 0.0001 | 0.4418 (0.2542, 0.6142) | < 0.0001 |
| | | | Targeted vs untargeted wastewater | 0.1506 (0.1075, 0.1956) | 0.1315 | 0.5480 (0.3745, 0.7090) | 0.3115 | 0.0144 (-0.2861, 0.2802) | 0.459 |
| | | Ottumwa, IA | Clinical vs targeted wastewater | 0.0850 (0.0507, 0.1380) | 0.001 | 0.7284 (0.5819, 0.8707) | 0.051 | 0.4665 (0.2176, 0.6876) | 0.001 |
| | | | Clinical vs untargeted wastewater | 0.1405 (0.1051, 0.1926) | 0.0285 | 0.6658 (0.5061, 0.8428) | 0.0605 | 0.2794 (0.0573, 0.5127) | 0.0105 |
| | | | Targeted vs untargeted wastewater | 0.1280 (0.0669, 0.2118) | 0.3095 | 0.6250 (0.4259, 0.8380) | 0.2395 | 0.0754 (-0.3184, 0.4374) | 0.3655 |
| | | Riverside, CA | Clinical vs targeted wastewater | 0.0717 (0.0469, 0.1037) | 0.0005 | 0.6915 (0.5525, 0.8542) | 0.043 | 0.3999 (0.1864, 0.6187) | 0.001 |
| | | | Clinical vs untargeted wastewater | 0.0557 (0.0381, 0.0807) | < 0.0001 | 0.7921 (0.6923, 0.8981) | < 0.0001 | 0.5869 (0.4045, 0.7344) | < 0.0001 |
| | | | Targeted vs untargeted wastewater | 0.0713 (0.0433, 0.1056) | < 0.0001 | 0.7361 (0.5764, 0.8854) | 0.0105 | 0.3957 (0.0358, 0.6710) | 0.0195 |
| | Alternative B | Boston, MA | Clinical vs targeted wastewater | 0.0810 (0.0545, 0.1197) | < 0.0001 | 0.8294 (0.6981, 0.9501) | < 0.0001 | 0.5238 (0.3656, 0.6744) | < 0.0001 |
| | | | Clinical vs untargeted wastewater | 0.1023 (0.0681, 0.1430) | < 0.0001 | 0.8063 (0.6778, 0.9225) | < 0.0001 | 0.4350 (0.2595, 0.5876) | < 0.0001 |
| | | | Targeted vs untargeted wastewater | 0.0866 (0.0604, 0.1236) | < 0.0001 | 0.7480 (0.6154, 0.8780) | < 0.0001 | 0.4727 (0.3170, 0.6056) | < 0.0001 |
| | | Boise, ID | Clinical vs targeted wastewater | 0.0893 (0.0541, 0.1323) | < 0.0001 | 0.6656 (0.5276, 0.8006) | 0.0265 | 0.3799 (0.1863, 0.5970) | 0.0005 |
| | | | Clinical vs untargeted wastewater | 0.1006 (0.0701, 0.1387) | < 0.0001 | 0.7519 (0.6259, 0.8760) | < 0.0001 | 0.4458 (0.2534, 0.6161) | < 0.0001 |
| | | | Targeted vs untargeted wastewater | 0.1535 (0.1088, 0.2037) | 0.1525 | 0.5387 (0.3714, 0.7029) | 0.356 | 0.0011 (-0.2935, 0.2789) | 0.4985 |
| | | Ottumwa, IA | Clinical vs targeted wastewater | 0.0818 (0.0495, 0.1391) | 0.0005 | 0.7371 (0.5862, 0.8836) | 0.0435 | 0.4909 (0.2369, 0.7011) | < 0.0001 |

| Virus | Shedding distribution | Location | Comparison | MAE (95% Crl) | MAE permutation *p* value | AOB1 (95% Crl) | AOB1 permutation *p* value | Tau (95% Crl) | Tau permutation *p* value |
|---|---|---|---|---|---|---|---|---|---|
| | | | Clinical vs untargeted wastewater | 0.1515 (0.1079, 0.3759) | 0.059 | 0.6658 (0.5135, 0.8329) | 0.099 | 0.2566 (0.0401, 0.4708) | 0.015 |
| | | | Targeted vs untargeted wastewater | 0.1481 (0.0756, 0.3678) | 0.437 | 0.5926 (0.3519, 0.8287) | 0.372 | 0.0387 (-0.4246, 0.4610) | 0.4455 |
| | | Riverside, CA | Clinical vs targeted wastewater | 0.0750 (0.0495, 0.1094) | < 0.0001 | 0.6881 (0.5458, 0.8441) | 0.0555 | 0.3667 (0.1479, 0.5808) | 0.0005 |
| | | | Clinical vs untargeted wastewater | 0.0582 (0.0399, 0.0846) | < 0.0001 | 0.7921 (0.6798, 0.8940) | < 0.0001 | 0.5747 (0.4050, 0.7243) | < 0.0001 |
| | | | Targeted vs untargeted wastewater | 0.0712 (0.0441, 0.1077) | < 0.0001 | 0.7292 (0.5694, 0.8785) | 0.0155 | 0.3844 (0.0244, 0.6706) | 0.021 |
| Influenza B | N/A | Boston, MA | Clinical vs targeted wastewater | 0.2441 (0.1667, 0.3520) | 0.023 | 0.6352 (0.4961, 0.7743) | 0.028 | 0.2272 (0.0469, 0.4297) | 0.0095 |
| | | | Clinical vs untargeted wastewater | 0.1388 (0.1111, 0.1773) | 0.0025 | 0.7042 (0.5739, 0.8081) | 0.001 | 0.2185 (0.0281, 0.4110) | 0.018 |
| | | | Targeted vs untargeted wastewater | 0.2066 (0.1359, 0.3166) | 0.0005 | 0.7135 (0.5491, 0.8674) | 0.0115 | 0.3818 (0.1555, 0.5887) | 0.0005 |
| | | Boise, ID | Clinical vs targeted wastewater | 0.0593 (0.0370, 0.0944) | 0.0035 | 0.7147 (0.5368, 0.8804) | 0.0135 | 0.3954 (0.1365, 0.6374) | 0.004 |
| | | | Clinical vs untargeted wastewater | 0.0896 (0.0649, 0.1238) | 0.0005 | 0.6492 (0.5213, 0.7810) | 0.0195 | 0.3759 (0.1718, 0.5397) | 0.0005 |
| | | | Targeted vs untargeted wastewater | 0.0854 (0.0536, 0.1326) | 0.0075 | 0.6254 (0.4613, 0.7988) | 0.0785 | 0.3045 (0.0551, 0.5556) | 0.01 |
| | | Ottumwa, IA | Clinical vs targeted wastewater | 0.1181 (0.0831, 0.1571) | 0.6155 | 0.5517 (0.3836, 0.7543) | 0.517 | -0.0035 (-0.2234, 0.2547) | 0.5195 |
| | | Riverside, CA | Clinical vs targeted wastewater | 0.0990 (0.0627, 0.1397) | 0.0015 | 0.6373 (0.4949, 0.7729) | 0.06 | 0.3096 (0.1182, 0.5053) | 0.0025 |
| | | | Clinical vs untargeted wastewater | 0.0614 (0.0417, 0.0865) | < 0.0001 | 0.7069 (0.5593, 0.8524) | 0.001 | 0.4938 (0.3212, 0.6547) | < 0.0001 |
| | | | Targeted vs untargeted wastewater | 0.0781 (0.0412, 0.1225) | < 0.0001 | 0.6892 (0.5035, 0.8785) | 0.026 | 0.4391 (0.1982, 0.7021) | < 0.0001 |
| RSV | Main analysis | Boston, MA | Clinical vs targeted wastewater | 0.1227 (0.0824, 0.1726) | < 0.0001 | 0.7323 (0.6246, 0.8583) | < 0.0001 | 0.4912 (0.3140, 0.6509) | < 0.0001 |

| Virus | Shedding distribution | Location | Comparison | MAE (95% CrI) | MAE permutation *p* value | AOB1 (95% CrI) | AOB1 permutation *p* value | Tau (95% CrI) | Tau permutation *p* value |
|---|---|---|---|---|---|---|---|---|---|
| | | | Clinical vs untargeted wastewater | 0.0940 (0.0627, 0.1341) | < 0.0001 | 0.8336 (0.6936, 0.9366) | < 0.0001 | 0.5557 (0.3519, 0.7159) | < 0.0001 |
| | | | Targeted vs untargeted wastewater | 0.1287 (0.0855, 0.1777) | 0.0005 | 0.7268 (0.5995, 0.8621) | 0.0005 | 0.4531 (0.2061, 0.6601) | 0.0005 |
| | | Boise, ID | Clinical vs targeted wastewater | 0.0969 (0.0533, 0.1587) | < 0.0001 | 0.7975 (0.6779, 0.9387) | < 0.0001 | 0.5755 (0.3454, 0.7778) | < 0.0001 |
| | | | Clinical vs untargeted wastewater | 0.1203 (0.0806, 0.1797) | 0.0005 | 0.7965 (0.6395, 0.9205) | 0.001 | 0.3871 (0.1386, 0.5978) | 0.0025 |
| | | | Targeted vs untargeted wastewater | 0.1362 (0.0782, 0.2129) | 0.003 | 0.6904 (0.5170, 0.8793) | 0.008 | 0.4142 (0.1002, 0.7104) | 0.0045 |
| | | Ottumwa, IA | Clinical vs targeted wastewater | 0.0862 (0.0506, 0.1456) | 0.0375 | 0.7457 (0.5775, 0.9267) | 0.38 | 0.3088 (-0.0473, 0.6584) | 0.046 |
| | | | Clinical vs untargeted wastewater | 0.1019 (0.0689, 0.1801) | < 0.0001 | 0.7690 (0.5725, 0.9140) | 0.0025 | 0.4502 (0.1381, 0.6980) | 0.0005 |
| | | | Targeted vs untargeted wastewater | 0.0817 (0.0342, 0.1593) | 0.003 | 0.7176 (0.4722, 0.9631) | 0.0485 | 0.3136 (-0.2213, 0.7019) | 0.137 |
| | | Riverside, CA | Clinical vs targeted wastewater | 0.1175 (0.0740, 0.1697) | 0.002 | 0.6339 (0.4915, 0.8203) | 0.1185 | 0.2868 (0.0346, 0.5906) | 0.014 |
| | | | Clinical vs untargeted wastewater | 0.0658 (0.0456, 0.0944) | < 0.0001 | 0.8191 (0.6860, 0.9231) | < 0.0001 | 0.6247 (0.4660, 0.7528) | < 0.0001 |
| | | | Targeted vs untargeted wastewater | 0.1187 (0.0683, 0.1733) | 0.009 | 0.5712 (0.4062, 0.7847) | 0.2935 | 0.2676 (-0.0197, 0.6120) | 0.0425 |
| | Alternative A | Boston, MA | Clinical vs targeted wastewater | 0.1286 (0.0881, 0.1793) | < 0.0001 | 0.7218 (0.6115, 0.8478) | < 0.0001 | 0.4701 (0.2927, 0.6294) | < 0.0001 |
| | | | Clinical vs untargeted wastewater | 0.0965 (0.0646, 0.1364) | < 0.0001 | 0.8257 (0.6884, 0.9296) | < 0.0001 | 0.5496 (0.3289, 0.7144) | < 0.0001 |
| | | | Targeted vs untargeted wastewater | 0.1308 (0.0851, 0.1830) | < 0.0001 | 0.7215 (0.5862, 0.8568) | 0.0005 | 0.4428 (0.1934, 0.6626) | < 0.0001 |
| | | Boise, ID | Clinical vs targeted wastewater | 0.0970 (0.0527, 0.1597) | < 0.0001 | 0.8006 (0.6840, 0.9387) | < 0.0001 | 0.5829 (0.3344, 0.7780) | < 0.0001 |
| | | | Clinical vs untargeted wastewater | 0.1205 (0.0767, 0.1795) | 0.0005 | 0.7946 (0.6434, 0.9187) | 0.0005 | 0.3881 (0.1381, 0.6042) | 0.0025 |

| Virus | Shedding distribution | Location | Comparison | MAE (95% CrI) | MAE permutation *p* value | AOB1 (95% CrI) | AOB1 permutation *p* value | Tau (95% CrI) | Tau permutation *p* value |
|---|---|---|---|---|---|---|---|---|---|
| | | | Targeted vs untargeted wastewater | 0.1379 (0.0800, 0.2130) | 0.0025 | 0.6966 (0.5262, 0.8855) | 0.009 | 0.4165 (0.1146, 0.6976) | 0.004 |
| | | Ottumwa, IA | Clinical vs targeted wastewater | 0.0871 (0.0517, 0.1480) | 0.0355 | 0.7457 (0.5862, 0.9267) | 0.3675 | 0.2955 (-0.0354, 0.6369) | 0.0475 |
| | | | Clinical vs untargeted wastewater | 0.1002 (0.0697, 0.1487) | < 0.0001 | 0.7789 (0.6167, 0.9091) | 0.0005 | 0.4678 (0.2366, 0.6896) | < 0.0001 |
| | | | Targeted vs untargeted wastewater | 0.0771 (0.0345, 0.1556) | 0.002 | 0.7361 (0.4722, 0.9722) | 0.0305 | 0.3199 (-0.2213, 0.7362) | 0.1225 |
| | | Riverside, CA | Clinical vs targeted wastewater | 0.1190 (0.0745, 0.1727) | 0.0035 | 0.6305 (0.4881, 0.8102) | 0.108 | 0.2908 (0.0437, 0.5777) | 0.0155 |
| | | | Clinical vs untargeted wastewater | 0.0650 (0.0434, 0.0937) | < 0.0001 | 0.8191 (0.6861, 0.9314) | < 0.0001 | 0.6302 (0.4715, 0.7590) | < 0.0001 |
| | | | Targeted vs untargeted wastewater | 0.1217 (0.0713, 0.1781) | 0.009 | 0.5625 (0.4132, 0.7604) | 0.3235 | 0.2592 (-0.0185, 0.5851) | 0.039 |
| Norovirus | N/A | Boston, MA | Clinical vs targeted wastewater | 0.0431 (0.0279, 0.0614) | < 0.0001 | 0.8005 (0.6745, 0.9161) | < 0.0001 | 0.4537 (0.2429, 0.6560) | < 0.0001 |
| | | | Clinical vs untargeted wastewater | 0.0379 (0.0284, 0.0491) | < 0.0001 | 0.7676 (0.6602, 0.8697) | < 0.0001 | 0.5348 (0.3865, 0.6723) | < 0.0001 |
| | | | Targeted vs untargeted wastewater | 0.0276 (0.0168, 0.0431) | < 0.0001 | 0.8912 (0.7771, 0.9788) | < 0.0001 | 0.6993 (0.5371, 0.8373) | < 0.0001 |
| | | Boise, ID | Clinical vs targeted wastewater | 0.0387 (0.0226, 0.0592) | 0.039 | 0.6503 (0.4816, 0.8191) | 0.0955 | 0.2519 (-0.0457, 0.5727) | 0.0555 |
| | | | Clinical vs untargeted wastewater | 0.0247 (0.0160, 0.0372) | 0.033 | 0.6182 (0.4360, 0.8043) | 0.143 | 0.2614 (-0.0259, 0.5318) | 0.039 |
| | | | Targeted vs untargeted wastewater | 0.0347 (0.0175, 0.0553) | 0.0255 | 0.7430 (0.5573, 0.9195) | 0.0275 | 0.1400 (-0.3039, 0.5467) | 0.276 |
| | | Ottumwa, IA | Clinical vs targeted wastewater | 0.0594 (0.0359, 0.0856) | 0.949 | 0.5776 (0.4137, 0.7846) | 0.917 | -0.2639 (-0.6484, 0.1028) | 0.924 |
| | | | Clinical vs untargeted wastewater | 0.0153 (0.0086, 0.0261) | 0.0005 | 0.7666 (0.5921, 0.9436) | 0.003 | 0.4900 (0.2035, 0.7582) | 0.0005 |
| | | | Targeted vs untargeted wastewater | 0.0538 (0.0309, 0.0825) | 0.021 | 0.7546 (0.5000, 1.0000) | 0.014 | -0.2212 (-0.6491, 0.2087) | 0.84 |

| Virus | Shedding distribution | Location | Comparison | MAE (95% CrI) | MAE permutation *p* value | AOB1 (95% CrI) | AOB1 permutation *p* value | Tau (95% CrI) | Tau permutation *p* value |
|---|---|---|---|---|---|---|---|---|---|
| | | Riverside, CA | Clinical vs targeted wastewater | 0.0529 (0.0369, 0.0775) | 0.529 | 0.6407 (0.4746, 0.8305) | 0.787 | -0.0908 (-0.4862, 0.2806) | 0.68 |
| | | | Clinical vs untargeted wastewater | 0.0318 (0.0275, 0.0375) | 0.003 | 0.6736 (0.5405, 0.8129) | 0.007 | 0.2792 (0.0675, 0.5169) | 0.005 |
| | | | Targeted vs untargeted wastewater | 0.0343 (0.0131, 0.0645) | < 0.0001 | 0.8090 (0.6528, 0.9861) | 0.0005 | 0.3901 (0.0102, 0.7482) | 0.0285 |
| Rotavirus | N/A | Boston, MA | Clinical vs untargeted wastewater | 0.0437 (0.0280, 0.0655) | < 0.0001 | 0.7852 (0.6180, 0.9208) | 0.0015 | 0.4375 (0.1501, 0.6770) | 0.0015 |
| | | Boise, ID | Clinical vs untargeted wastewater | 0.0524 (0.0299, 0.0905) | 0.1205 | 0.6453 (0.4418, 0.8120) | 0.1005 | 0.2193 (-0.1027, 0.5220) | 0.106 |
| | | Ottumwa, IA | Clinical vs untargeted wastewater | 0.0627 (0.0361, 0.1152) | 0.0345 | 0.6388 (0.4644, 0.8207) | 0.0485 | 0.2918 (0.0342, 0.5514) | 0.014 |
| | | Riverside, CA | Clinical vs untargeted wastewater | 0.0315 (0.0202, 0.0494) | < 0.0001 | 0.8087 (0.6465, 0.9293) | < 0.0001 | 0.4607 (0.2494, 0.6529) | < 0.0001 |

Refer to **Table S4** and **Table S7** for primary and alternative fecal shedding distributions. Permutation *p* values represent the proportion of posterior draws in which the null was more similar than the observed comparison; *p* values < 0.05 indicate significant similarity.

Abbreviations: AOB = agreement on binary, CrI = credible interval, MAE = mean absolute error, RSV = respiratory syncytial virus.

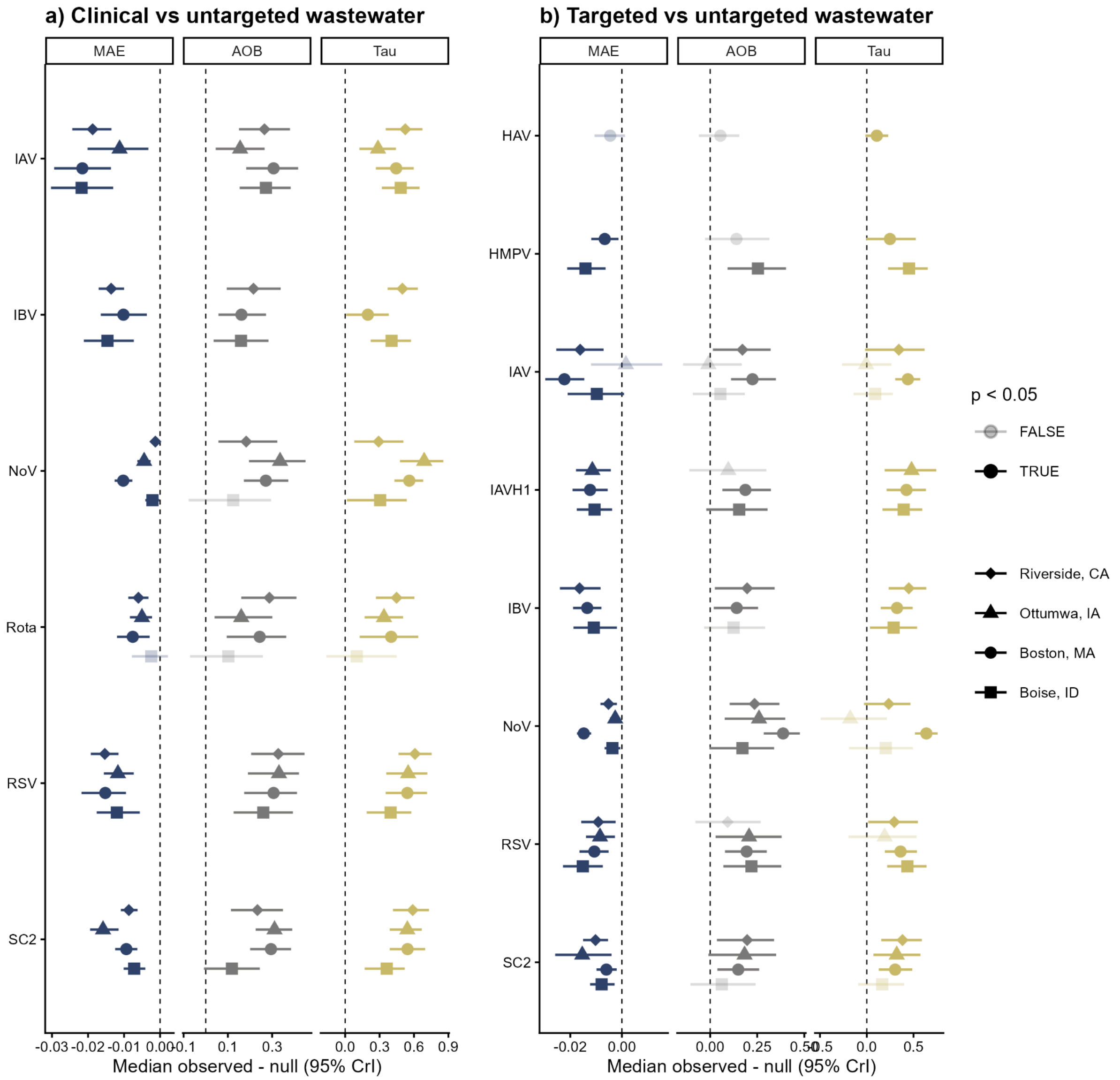

**Fig. S13. Concordance between $r_t$ estimates derived from different data sources.**
Difference between the observed and null posterior distributions of $r_t$ estimates derived from (**a**) Epic Cosmos clinical surveillance versus untargeted wastewater monitoring and (**b**) targeted versus untargeted wastewater monitoring data. Symbols denote median differences for unique locations with error bars representing 95% CrIs; darker hues indicate significantly similar derived $r_t$ estimates. For NoV, clinical data include all genogroups whereas wastewater monitoring data sources are specific to genogroup II. For all sites, $r_t$ estimates were only derived from wastewater monitoring data sources if at least 75% of samples had a positive detection. Here, $p$ values represent Bayesian permutation probability values, with $p < 0.05$ indicating significant concordance. Abbreviations: AOB = agreement on binary, CrI = credible interval, HAV = hepatitis A virus, HMPV = human metapneumovirus, IAV = influenza A virus, IAVH1 = IAV H1N1, IBV = influenza B virus, MAE = mean absolute error, NoV = norovirus, Rota = rotavirus, RSV = respiratory syncytial virus, SC2 = SARS-CoV-2.

**Table S6. Observed posterior distributions for metrics of concordance among derived $r_t$ estimates**

| Virus | Location | Comparison | MAE (95% CrI) | MAE permutation *p* value | AOB0 (95% CrI) | AOB0 permutation *p* value | Tau (95% CrI) | Tau permutation *p* value |
|---|---|---|---|---|---|---|---|---|
| SARS-CoV-2 | Boston, MA | Clinical vs targeted wastewater | 0.0136 (0.0102, 0.0171) | < 0.0001 | 0.7248 (0.6185, 0.8256) | < 0.0001 | 0.4171 (0.2827, 0.5492) | < 0.0001 |
| | | Clinical vs untargeted wastewater | 0.0093 (0.0066, 0.0124) | < 0.0001 | 0.8100 (0.7204, 0.9015) | < 0.0001 | 0.5457 (0.3981, 0.6931) | < 0.0001 |
| | | Targeted vs untargeted wastewater | 0.0165 (0.0123, 0.0214) | < 0.0001 | 0.6531 (0.5501, 0.7615) | 0.005 | 0.3063 (0.1274, 0.4702) | < 0.0001 |
| | Boise, ID | Clinical vs targeted wastewater | 0.0115 (0.0080, 0.0153) | < 0.0001 | 0.6635 (0.5031, 0.8176) | 0.029 | 0.3107 (0.1056, 0.5182) | < 0.0001 |
| | | Clinical vs untargeted wastewater | 0.0146 (0.0111, 0.0186) | < 0.0001 | 0.6398 (0.5196, 0.7520) | 0.041 | 0.3588 (0.1726, 0.5077) | 0.001 |
| | | Targeted vs untargeted wastewater | 0.0167 (0.0117, 0.0226) | < 0.0001 | 0.5619 (0.3968, 0.7333) | 0.243 | 0.1620 (-0.0893, 0.3860) | 0.109 |
| | Ottumwa, IA | Clinical vs targeted wastewater | 0.0294 (0.0219, 0.0373) | 0.014 | 0.7000 (0.5864, 0.8182) | 0.003 | 0.2622 (0.0856, 0.4541) | 0.005 |
| | | Clinical vs untargeted wastewater | 0.0231 (0.0181, 0.0286) | < 0.0001 | 0.8321 (0.7494, 0.8948) | < 0.0001 | 0.5422 (0.4070, 0.6625) | < 0.0001 |
| | | Targeted vs untargeted wastewater | 0.0292 (0.0188, 0.0419) | 0.004 | 0.7067 (0.5239, 0.8798) | 0.031 | 0.3278 (0.0816, 0.5676) | 0.003 |
| | Riverside, CA | Clinical vs targeted wastewater | 0.0209 (0.0158, 0.0273) | 0.002 | 0.6643 (0.4841, 0.8269) | 0.039 | 0.3031 (0.0676, 0.5668) | 0.009 |
| | | Clinical vs untargeted wastewater | 0.0101 (0.0078, 0.0128) | < 0.0001 | 0.7696 (0.6638, 0.8689) | < 0.0001 | 0.5864 (0.4330, 0.7240) | < 0.0001 |
| | | Targeted vs untargeted wastewater | 0.0191 (0.0139, 0.0256) | < 0.0001 | 0.6839 (0.5321, 0.8250) | 0.006 | 0.3878 (0.1668, 0.5712) | < 0.0001 |
| Influenza A | Boston, MA | Clinical vs targeted wastewater | 0.0280 (0.0214, 0.0370) | < 0.0001 | 0.8147 (0.7057, 0.9047) | < 0.0001 | 0.4756 (0.3316, 0.6042) | < 0.0001 |
| | | Clinical vs untargeted wastewater | 0.0320 (0.0237, 0.0415) | < 0.0001 | 0.8065 (0.6810, 0.9033) | < 0.0001 | 0.4413 (0.2821, 0.5878) | < 0.0001 |
| | | Targeted vs untargeted wastewater | 0.0306 (0.0229, 0.0390) | < 0.0001 | 0.7236 (0.6260, 0.8374) | < 0.0001 | 0.4426 (0.3174, 0.5536) | < 0.0001 |
| | Boise, ID | Clinical vs targeted | 0.0306 (0.0229, | < 0.0001 | 0.6792 (0.5503, | 0.009 | 0.4118 (0.2477, | < 0.0001 |

| Virus | Location | Comparison | MAE (95% CrI) | MAE permutation *p* value | AOB0 (95% CrI) | AOB0 permutation *p* value | Tau (95% CrI) | Tau permutation *p* value |
|---|---|---|---|---|---|---|---|---|
| | | wastewater | 0.0397) | | 0.8019) | | 0.5563) | |
| | | Clinical vs untargeted wastewater | 0.0292 (0.0215, 0.0376) | < 0.0001 | 0.7717 (0.6515, 0.8780) | < 0.0001 | 0.4837 (0.3289, 0.6369) | < 0.0001 |
| | | Targeted vs untargeted wastewater | 0.0485 (0.0377, 0.0602) | 0.039 | 0.5524 (0.4159, 0.6762) | 0.248 | 0.0897 (-0.1299, 0.2743) | 0.207 |
| | Ottumwa, IA | Clinical vs targeted wastewater | 0.0448 (0.0331, 0.0582) | 0.002 | 0.6773 (0.5545, 0.7955) | 0.032 | 0.3196 (0.1441, 0.4703) | 0.003 |
| | | Clinical vs untargeted wastewater | 0.0451 (0.0378, 0.0525) | 0.004 | 0.6842 (0.5814, 0.7769) | 0.007 | 0.2873 (0.1272, 0.4268) | < 0.0001 |
| | | Targeted vs untargeted wastewater | 0.0649 (0.0481, 0.0835) | 0.588 | 0.5240 (0.3942, 0.6636) | 0.562 | -0.0051 (-0.2603, 0.2478) | 0.518 |
| | Riverside, CA | Clinical vs targeted wastewater | 0.0248 (0.0188, 0.0319) | < 0.0001 | 0.7385 (0.5971, 0.8551) | 0.008 | 0.3809 (0.1810, 0.5553) | < 0.0001 |
| | | Clinical vs untargeted wastewater | 0.0204 (0.0154, 0.0261) | < 0.0001 | 0.7822 (0.6660, 0.8816) | < 0.0001 | 0.5245 (0.3697, 0.6604) | < 0.0001 |
| | | Targeted vs untargeted wastewater | 0.0255 (0.0180, 0.0353) | < 0.0001 | 0.7107 (0.5606, 0.8464) | 0.021 | 0.3502 (0.0000, 0.6038) | 0.032 |
| Influenza B | Boston, MA | Clinical vs targeted wastewater | 0.0476 (0.0353, 0.0613) | 0.024 | 0.5613 (0.4687, 0.6649) | 0.137 | 0.1464 (0.0015, 0.2874) | 0.042 |
| | | Clinical vs untargeted wastewater | 0.0422 (0.0339, 0.0529) | 0.001 | 0.6828 (0.5770, 0.7922) | 0.003 | 0.1970 (0.0154, 0.3742) | 0.023 |
| | | Targeted vs untargeted wastewater | 0.0289 (0.0224, 0.0357) | < 0.0001 | 0.6369 (0.5257, 0.7425) | 0.011 | 0.3195 (0.1712, 0.4724) | < 0.0001 |
| | Boise, ID | Clinical vs targeted wastewater | 0.0174 (0.0121, 0.0235) | 0.001 | 0.7060 (0.5377, 0.8553) | 0.009 | 0.3757 (0.1575, 0.5957) | < 0.0001 |
| | | Clinical vs untargeted wastewater | 0.0247 (0.0187, 0.0322) | < 0.0001 | 0.6713 (0.5531, 0.7953) | 0.004 | 0.4109 (0.2373, 0.5663) | < 0.0001 |
| | | Targeted vs untargeted wastewater | 0.0260 (0.0183, 0.0358) | 0.011 | 0.6349 (0.4794, 0.7937) | 0.057 | 0.2816 (0.0502, 0.5197) | 0.015 |
| | Ottumwa, IA | Clinical vs targeted wastewater | 0.0363 (0.0285, 0.0439) | 0.442 | 0.5591 (0.4227, 0.6955) | 0.453 | 0.0297 (-0.1300, 0.2180) | 0.368 |

| Virus | Location | Comparison | MAE (95% CrI) | MAE permutation *p* value | AOB0 (95% CrI) | AOB0 permutation *p* value | Tau (95% CrI) | Tau permutation *p* value |
|---|---|---|---|---|---|---|---|---|
| | Riverside, CA | Clinical vs targeted wastewater | 0.0301 (0.0232, 0.0379) | < 0.0001 | 0.6466 (0.5194, 0.7633) | 0.02 | 0.3509 (0.1826, 0.4980) | < 0.0001 |
| | | Clinical vs untargeted wastewater | 0.0183 (0.0136, 0.0232) | < 0.0001 | 0.7230 (0.6089, 0.8415) | < 0.0001 | 0.4982 (0.3763, 0.6194) | < 0.0001 |
| | | Targeted vs untargeted wastewater | 0.0259 (0.0179, 0.0361) | < 0.0001 | 0.6893 (0.5321, 0.8250) | 0.012 | 0.4491 (0.2571, 0.6177) | < 0.0001 |
| RSV | Boston, MA | Clinical vs targeted wastewater | 0.0298 (0.0232, 0.0360) | < 0.0001 | 0.6975 (0.6021, 0.8011) | < 0.0001 | 0.4027 (0.2834, 0.5325) | < 0.0001 |
| | | Clinical vs untargeted wastewater | 0.0169 (0.0117, 0.0224) | < 0.0001 | 0.8118 (0.6935, 0.9104) | < 0.0001 | 0.5432 (0.3640, 0.6977) | < 0.0001 |
| | | Targeted vs untargeted wastewater | 0.0299 (0.0231, 0.0364) | < 0.0001 | 0.6856 (0.5881, 0.7778) | < 0.0001 | 0.3587 (0.2043, 0.5073) | < 0.0001 |
| | Boise, ID | Clinical vs targeted wastewater | 0.0219 (0.0156, 0.0292) | < 0.0001 | 0.8082 (0.6917, 0.9308) | < 0.0001 | 0.5490 (0.3631, 0.6899) | < 0.0001 |
| | | Clinical vs untargeted wastewater | 0.0212 (0.0150, 0.0283) | < 0.0001 | 0.7992 (0.6612, 0.9213) | < 0.0001 | 0.4009 (0.1967, 0.5730) | < 0.0001 |
| | | Targeted vs untargeted wastewater | 0.0262 (0.0186, 0.0347) | < 0.0001 | 0.7095 (0.5746, 0.8571) | 0.002 | 0.4360 (0.2289, 0.6265) | < 0.0001 |
| | Ottumwa, IA | Clinical vs targeted wastewater | 0.0222 (0.0158, 0.0301) | 0.04 | 0.7045 (0.5636, 0.8545) | 0.186 | 0.1731 (-0.0283, 0.3676) | 0.061 |
| | | Clinical vs untargeted wastewater | 0.0156 (0.0128, 0.0192) | < 0.0001 | 0.8346 (0.7017, 0.9198) | < 0.0001 | 0.5498 (0.3667, 0.7046) | < 0.0001 |
| | | Targeted vs untargeted wastewater | 0.0209 (0.0137, 0.0298) | 0.001 | 0.6875 (0.4806, 0.8462) | 0.017 | 0.1859 (-0.1944, 0.5211) | 0.177 |
| | Riverside, CA | Clinical vs targeted wastewater | 0.0229 (0.0170, 0.0293) | < 0.0001 | 0.6608 (0.5265, 0.7915) | 0.037 | 0.3368 (0.1254, 0.5257) | < 0.0001 |
| | | Clinical vs untargeted wastewater | 0.0119 (0.0088, 0.0159) | < 0.0001 | 0.8266 (0.7040, 0.9408) | < 0.0001 | 0.6103 (0.4749, 0.7336) | < 0.0001 |
| | | Targeted vs untargeted wastewater | 0.0236 (0.0160, 0.0322) | 0.004 | 0.6000 (0.4500, 0.7643) | 0.177 | 0.2960 (0.0178, 0.5415) | 0.019 |
| Norovirus | Boston, MA | Clinical vs targeted wastewater | 0.0145 (0.0112, 0.0176) | < 0.0001 | 0.7956 (0.6839, 0.9046) | < 0.0001 | 0.4231 (0.2490, 0.6012) | < 0.0001 |

| Virus | Location | Comparison | MAE (95% CrI) | MAE permutation *p* value | AOB0 (95% CrI) | AOB0 permutation *p* value | Tau (95% CrI) | Tau permutation *p* value |
|---|---|---|---|---|---|---|---|---|
| | | Clinical vs untargeted wastewater | 0.0111 (0.0088, 0.0135) | < 0.0001 | 0.7742 (0.6792, 0.8692) | < 0.0001 | 0.5602 (0.4406, 0.6706) | < 0.0001 |
| | | Targeted vs untargeted wastewater | 0.0099 (0.0074, 0.0128) | < 0.0001 | 0.8862 (0.8022, 0.9593) | < 0.0001 | 0.6384 (0.5343, 0.7342) | < 0.0001 |
| | Boise, ID | Clinical vs targeted wastewater | 0.0122 (0.0092, 0.0160) | 0.033 | 0.6572 (0.4906, 0.8082) | 0.066 | 0.2633 (-0.0097, 0.5058) | 0.035 |
| | | Clinical vs untargeted wastewater | 0.0076 (0.0057, 0.0100) | 0.016 | 0.6398 (0.4407, 0.8110) | 0.105 | 0.3072 (0.0162, 0.5271) | 0.018 |
| | | Targeted vs untargeted wastewater | 0.0109 (0.0075, 0.0148) | 0.007 | 0.7619 (0.5778, 0.8984) | 0.031 | 0.1996 (-0.1891, 0.4907) | 0.145 |
| | Ottumwa, IA | Clinical vs targeted wastewater | 0.0175 (0.0136, 0.0220) | 0.976 | 0.5909 (0.4364, 0.7819) | 0.895 | -0.2285 (-0.4523, 0.0374) | 0.956 |
| | | Clinical vs untargeted wastewater | 0.0036 (0.0027, 0.0045) | < 0.0001 | 0.8358 (0.6716, 0.9474) | < 0.0001 | 0.6889 (0.4830, 0.8297) | < 0.0001 |
| | | Targeted vs untargeted wastewater | 0.0160 (0.0123, 0.0206) | 0.005 | 0.7885 (0.5337, 0.9279) | 0.005 | -0.1844 (-0.4868, 0.1894) | 0.824 |
| | Riverside, CA | Clinical vs targeted wastewater | 0.0190 (0.0150, 0.0235) | 0.254 | 0.6537 (0.5052, 0.7881) | 0.306 | 0.0294 (-0.2958, 0.3265) | 0.435 |
| | | Clinical vs untargeted wastewater | 0.0099 (0.0087, 0.0111) | 0.004 | 0.6786 (0.5623, 0.8161) | 0.002 | 0.2862 (0.0882, 0.4879) | 0.004 |
| | | Targeted vs untargeted wastewater | 0.0159 (0.0116, 0.0216) | < 0.0001 | 0.7250 (0.6178, 0.8321) | < 0.0001 | 0.2361 (-0.0153, 0.4562) | 0.042 |
| Rotavirus | Boston, MA | Clinical vs untargeted wastewater | 0.0143 (0.0098, 0.0192) | 0.001 | 0.7832 (0.6253, 0.8997) | < 0.0001 | 0.3979 (0.1409, 0.6332) | 0.002 |
| | Boise, ID | Clinical vs untargeted wastewater | 0.0214 (0.0127, 0.0312) | 0.144 | 0.6102 (0.4310, 0.7737) | 0.125 | 0.1014 (-0.1562, 0.4480) | 0.239 |
| | Ottumwa, IA | Clinical vs untargeted wastewater | 0.0220 (0.0179, 0.0266) | < 0.0001 | 0.6541 (0.5212, 0.7945) | 0.008 | 0.3367 (0.1807, 0.4864) | < 0.0001 |
| | Riverside, CA | Clinical vs untargeted wastewater | 0.0111 (0.0082, 0.0144) | < 0.0001 | 0.7865 (0.6681, 0.8965) | < 0.0001 | 0.4436 (0.2723, 0.5928) | < 0.0001 |
| Human metapnuemo | Boston, MA | Targeted vs untargeted wastewater | 0.0187 (0.0131, 0.0254) | 0.008 | 0.6396 (0.4796, 0.8104) | 0.051 | 0.2425 (-0.0030, 0.5193) | 0.029 |

| Virus | Location | Comparison | MAE (95% CrI) | MAE permutation *p* value | AOB0 (95% CrI) | AOB0 permutation *p* value | Tau (95% CrI) | Tau permutation *p* value |
|---|---|---|---|---|---|---|---|---|
| virus | Boise, ID | Targeted vs untargeted wastewater | 0.0203 (0.0128, 0.0283) | < 0.0001 | 0.7556 (0.6031, 0.9016) | 0.002 | 0.4488 (0.2553, 0.6357) | < 0.0001 |
| Influenza A H1N1 | Boston, MA | Targeted vs untargeted wastewater | 0.0349 (0.0264, 0.0435) | < 0.0001 | 0.6883 (0.5718, 0.8157) | 0.001 | 0.4245 (0.2187, 0.6120) | < 0.0001 |
| | Boise, ID | Targeted vs untargeted wastewater | 0.0209 (0.0147, 0.0283) | 0.003 | 0.6678 (0.5034, 0.8185) | 0.042 | 0.3981 (0.1758, 0.5855) | < 0.0001 |
| | Ottumwa, IA | Targeted vs untargeted wastewater | 0.0140 (0.0086, 0.0218) | 0.002 | 0.6779 (0.4760, 0.8606) | 0.184 | 0.4762 (0.2035, 0.7291) | < 0.0001 |
| Hepatitis A virus | Boston, MA | Targeted vs untargeted wastewater | 0.0762 (0.0655, 0.0881) | 0.072 | 0.5556 (0.4526, 0.6531) | 0.198 | 0.1143 (0.0067, 0.2050) | 0.036 |

Permutation *p* values represent the proportion of posterior draws in which the null was more similar than the observed comparison; *p* values < 0.05 indicate significant similarity.

Abbreviations: AOB = agreement on binary, CrI = credible interval, MAE = mean absolute error, RSV = respiratory syncytial virus.

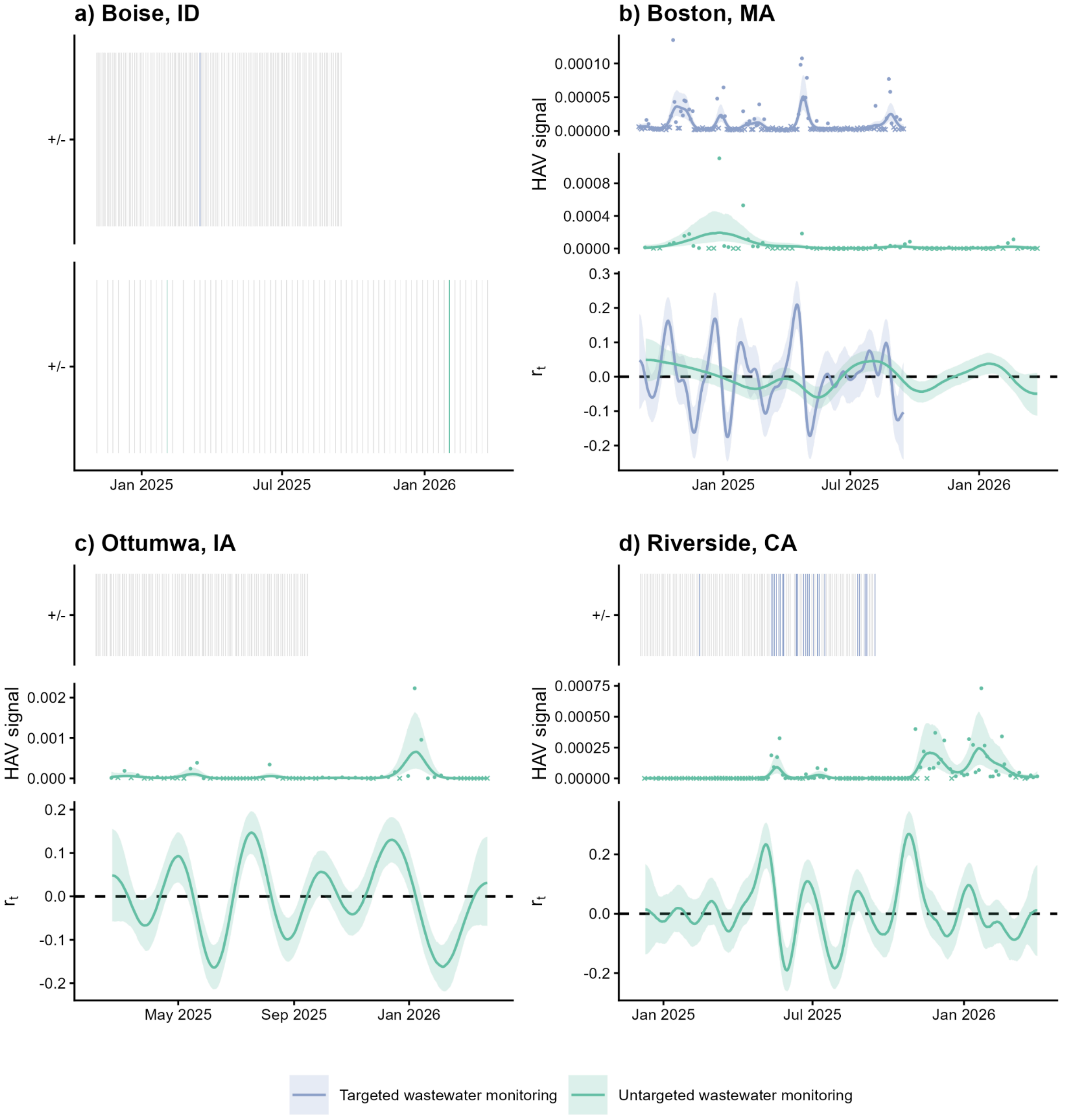

**Fig. S14. Hepatitis A surveillance data and $r_t$ estimates for all locations.**

Time series data from each data source with GAM smoothing alongside $r_t$ estimates are shown for (a) Boise, Idaho; (b) Boston, Massachusetts; (c) Ottumwa, Iowa; and (d) Riverside, California. "Signal" indicates PMMoV-normalized concentrations of HAV RNA for targeted wastewater data (purple) and PMMoV-normalized HAV clade counts scaled to account for sequencing depth for untargeted wastewater data (green). For wastewater monitoring data sources, x's denote non-detect samples; outlier measurements are not shown for visualization. Lines represent median estimates with 90% CrIs shaded. Heatmaps showing presence/absence are used when >75% of samples did not result in a positive virus detection. Abbreviations: CrI = credible interval, GAM = generalized additive model, HAV = hepatitis A virus, PMMoV = pepper mild mottle virus.

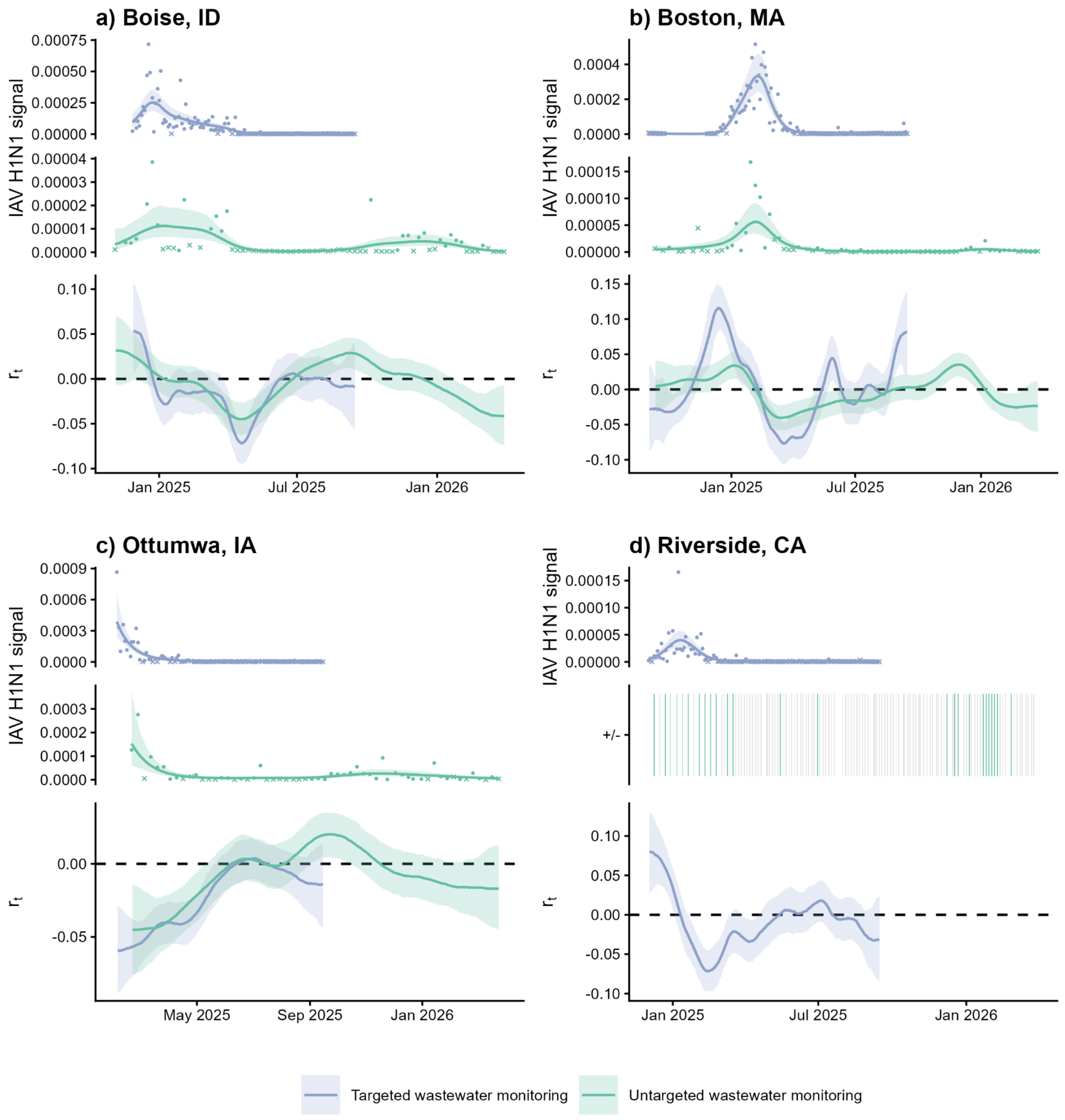

**Fig. S15. Influenza A H1N1 surveillance data and $r_t$ estimates for all locations.**

Time series data from each data source with GAM smoothing alongside $r_t$ estimates are shown for (a) Boise, Idaho; (b) Boston, Massachusetts; (c) Ottumwa, Iowa; and (d) Riverside, California. “Signal” indicates PMMoV-normalized concentrations of IAV H1N1 RNA for targeted wastewater data (purple) and PMMoV-normalized IAV H1N1 clade counts scaled to account for sequencing depth for untargeted wastewater data (green). For wastewater monitoring data sources, x’s denote non-detect samples; outlier measurements are not shown for visualization. Lines represent median estimates with 90% CrIs shaded. Heatmaps showing presence/absence are used when >75% of samples did not result in a positive virus detection. Abbreviations: CrI = credible interval, GAM = generalized additive model, IAV H1N1 = influenza A virus H1N1, PMMoV = pepper mild mottle virus.

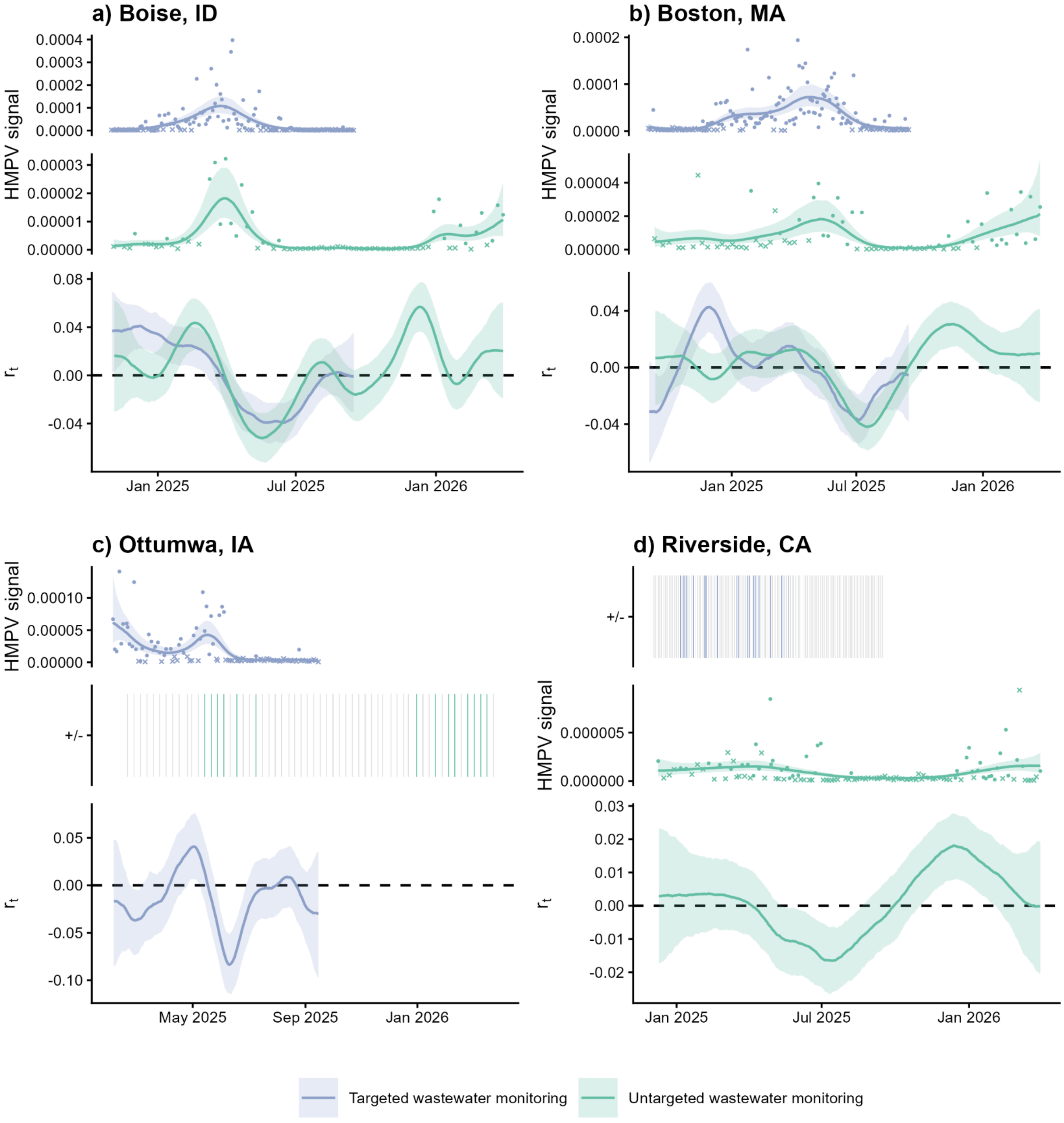

**Fig. S16. Human metapneumovirus surveillance data and $r_t$ estimates for all locations.**

Time series data from each data source with GAM smoothing alongside $r_t$ estimates are shown for (a) Boise, Idaho; (b) Boston, Massachusetts; (c) Ottumwa, Iowa; and (d) Riverside, California. "Signal" indicates PMMoV-normalized concentrations of HMPV RNA for targeted wastewater data (purple) and PMMoV-normalized HMPV clade counts scaled to account for sequencing depth for untargeted wastewater data (green). For wastewater monitoring data sources, x's denote non-detect samples; outlier measurements are not shown for visualization. Lines represent median estimates with 90% CrIs shaded. Heatmaps showing presence/absence are used when >75% of samples did not result in a positive virus detection. Abbreviations: CrI = credible interval, GAM = generalized additive model, HMPV = human metapneumovirus, PMMoV = pepper mild mottle virus.

**Table S7. Alternative fecal shedding load distributions used for $R_t$ sensitivity analyses**

| Virus | Alternative fecal shedding load distribution(s) (days) | Source |
|---|---|---|
| SARS-CoV-2 | *Alternative A:* Gamma(mean = 5.3, sd = 3.2, max = 14) + Gamma (mean = 6.7, sd = 7.0, max = 30)<br><br>*Alternative B:* Gamma(mean = 5.3, sd = 3.2, max = 14) + Gamma (mean = 4.7, sd = 1.7, max = 30)<br><br>*Alternative C:* Gamma (shape = 2.05, rate = 0.169, max = 30) | *Alternative A:* Fecal shedding load distribution used in the main analysis of Huisman et al. (2022).[37] The incubation period from Linton et al. (2020)[38] is convolved with the gastrointestinal shedding distribution based on days since symptom onset from Benefield et al. (2020).[39]<br><br>*Alternative B:* Fecal shedding load distribution used as a supplementary analysis in Huisman et al. (2022).[37] The incubation period from Linton et al. (2020)[38] is convolved with the gastrointestinal shedding distribution based on days since symptom onset from Han et al. (2020).[40]<br><br>*Alternative C:* Shedding load distribution (including all shedding routes) based on a modeling study by Cavany et al. (2022).[41] We used the model parameters of the baseline 2-day scenario. |
| Influenza A (IAV) | *Alternative A:* Gamma (mean = 2.5, sd = 1, max = 10)<br><br>*Alternative B:* NonParametric(pmf = 0.000000000, 0.182498915, 0.149417474, 0.122332681, 0.100157528, 0.082002048, 0.067137599, 0.054967617, 0.045003678, 0.036845895, 0.030166868, 0.024698542, 0.020221456, 0.016555928, 0.013554847, 0.011097770, 0.009086086, 0.007439058, 0.006090586, 0.004986550, 0.004082642, 0.003342584, 0.002736676, 0.002240601, 0.001834449, 0.001501920) | *Alternative A:* Respiratory shedding load distribution used in the main analysis of Nadeau et al. (2024)[30] based on a respiratory shedding load distribution from Carrat et al (2008).[42] Although we define the infection-to-detection delay as being characterized by fecal shedding dynamics, here we test this delay as being characterized by respiratory shedding dynamics.<br><br>*Alternative B:* Shedding load distribution (including all shedding routes) based on a modeling study by Ando et al. (2025).[43] Specifically, we used the median posterior estimates of the model parameters to predict concentrations in wastewater solids (cp/g-solid/case) on days 0–25; we then normalized the pmf. We assume days since shedding onset corresponds to days since infection. |
| Respiratory Syncytial Virus (RSV) | *Alternative A:* See IAV (**Table S4**) | *Alternative A:* See IAV (**Table S4**). As a sensitivity analysis, we also tested an influenza-like fecal shedding distribution. |

Abbreviations: cp/g = copies per gram, pmf = probability mass function, sd = standard deviation.

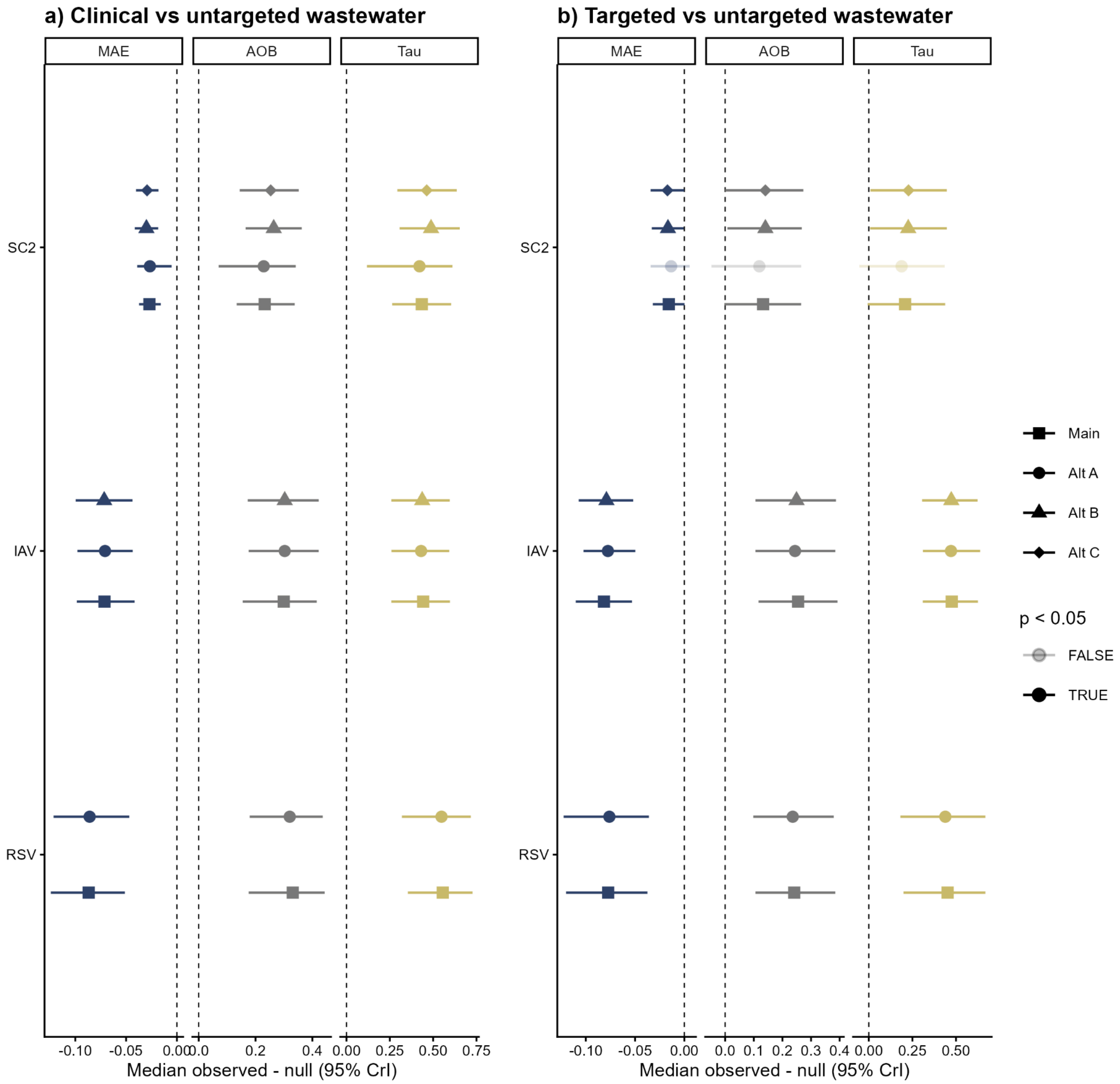

**Fig. S17. Concordance between $R_t$ estimates derived from different data sources using alternative shedding distributions, Boston, Massachusetts.**

Difference between the observed and null posterior distributions of $r_t$ estimates derived from (**a**) Epic Cosmos clinical surveillance versus untargeted wastewater monitoring and (**b**) targeted versus untargeted wastewater monitoring data. Symbols denote median differences for unique shedding distributions with error bars representing 95% CrIs; darker hues indicate significantly similar derived $r_t$ estimates. Here, $p$ values represent Bayesian permutation probability values, with $p < 0.05$ indicating significant concordance. Abbreviations: AOB = agreement on binary, Alt = alternative, CrI = credible interval, IAV = influenza A virus, MAE = mean absolute error, RSV = respiratory syncytial virus, SC2 = SARS-CoV-2.

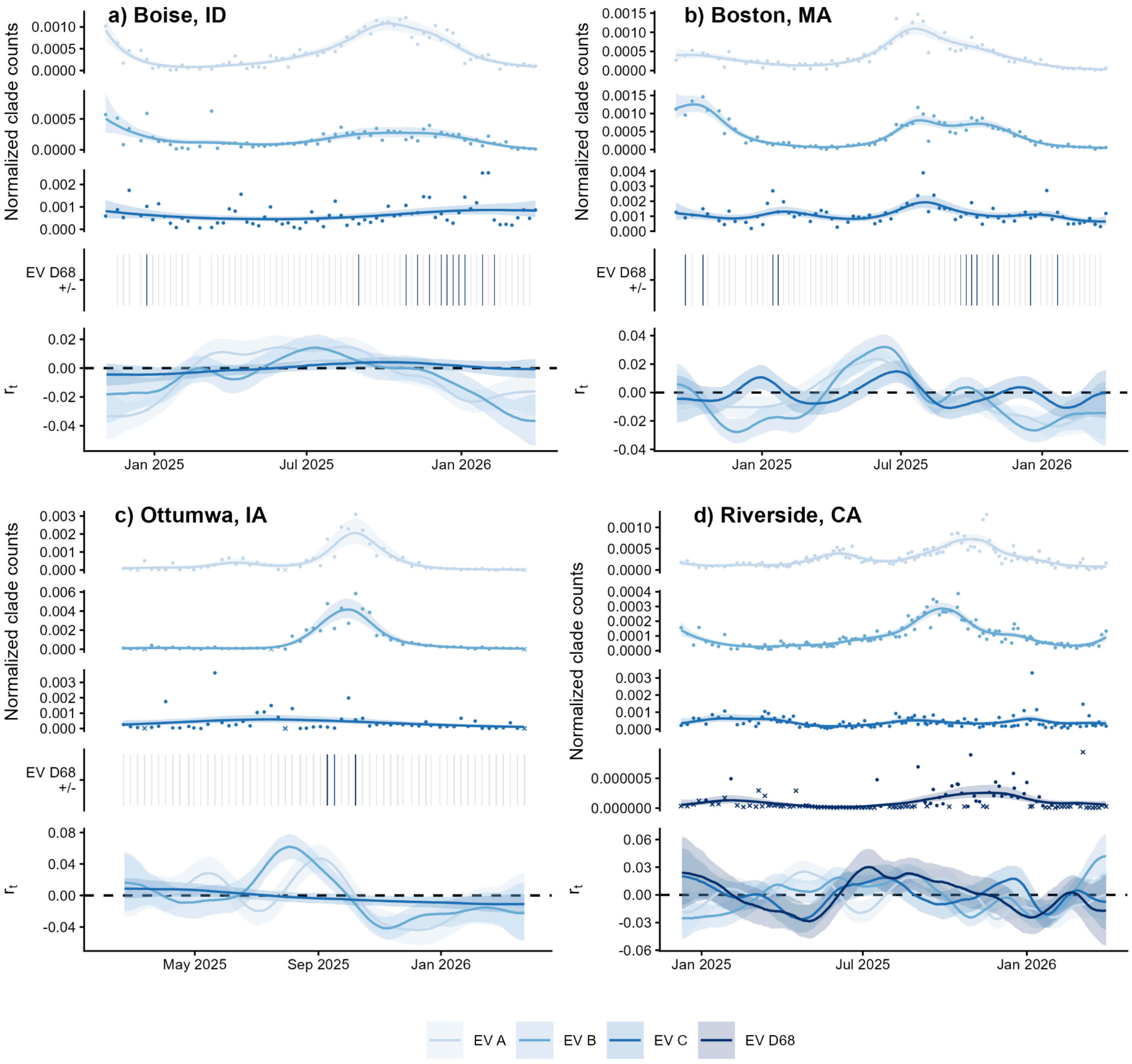

**Fig. S18. Untargeted enterovirus wastewater monitoring data and $r_t$ estimates for all locations, differentiated by species/type.**

Untargeted wastewater monitoring time series data with GAM smoothing alongside $r_t$ estimates are shown for (a) Boise, Idaho; (b) Boston, Massachusetts; (c) Ottumwa, Iowa; and (d) Riverside, California. EV clade counts are normalized by PMMoV clade counts and scaled to account for sequencing depth; x's denote non-detect samples and outlier measurements are not shown for visualization. Lines represent median estimates with 90% CrIs shaded. Heatmaps showing presence/absence are used when >75% of samples did not result in a positive virus detection. Abbreviations: CrI = credible interval, EV = enterovirus, GAM = generalized additive model, PMMoV = pepper mild mottle virus.

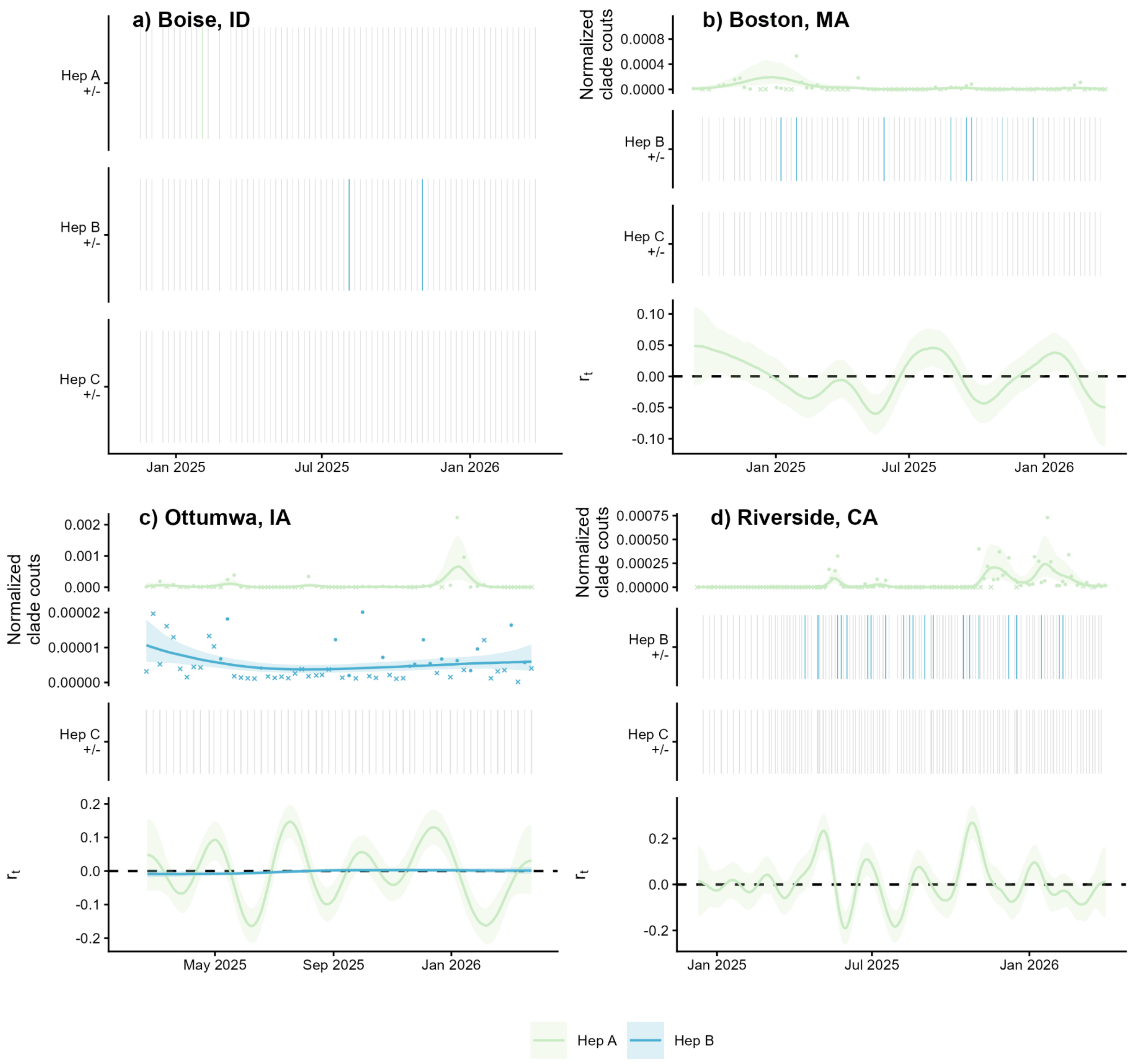

**Fig. S19. Untargeted hepatitis virus wastewater monitoring data and $r_t$ estimates for all locations, differentiated by virus.**

Untargeted wastewater monitoring time series data with GAM smoothing alongside $r_t$ estimates are shown for (a) Boise, Idaho; (b) Boston, Massachusetts; (c) Ottumwa, Iowa; and (d) Riverside, California. Hepatitis ("Hep") clade counts are normalized by PMMoV clade counts and scaled to account for sequencing depth; x's denote non-detect samples and outlier measurements are not shown for visualization. Lines represent median estimates with 90% CrIs shaded. Heatmaps showing presence/absence are used when >75% of samples did not result in a positive virus detection. Abbreviations: CrI = credible interval, GAM = generalized additive model, PMMoV = pepper mild mottle virus.

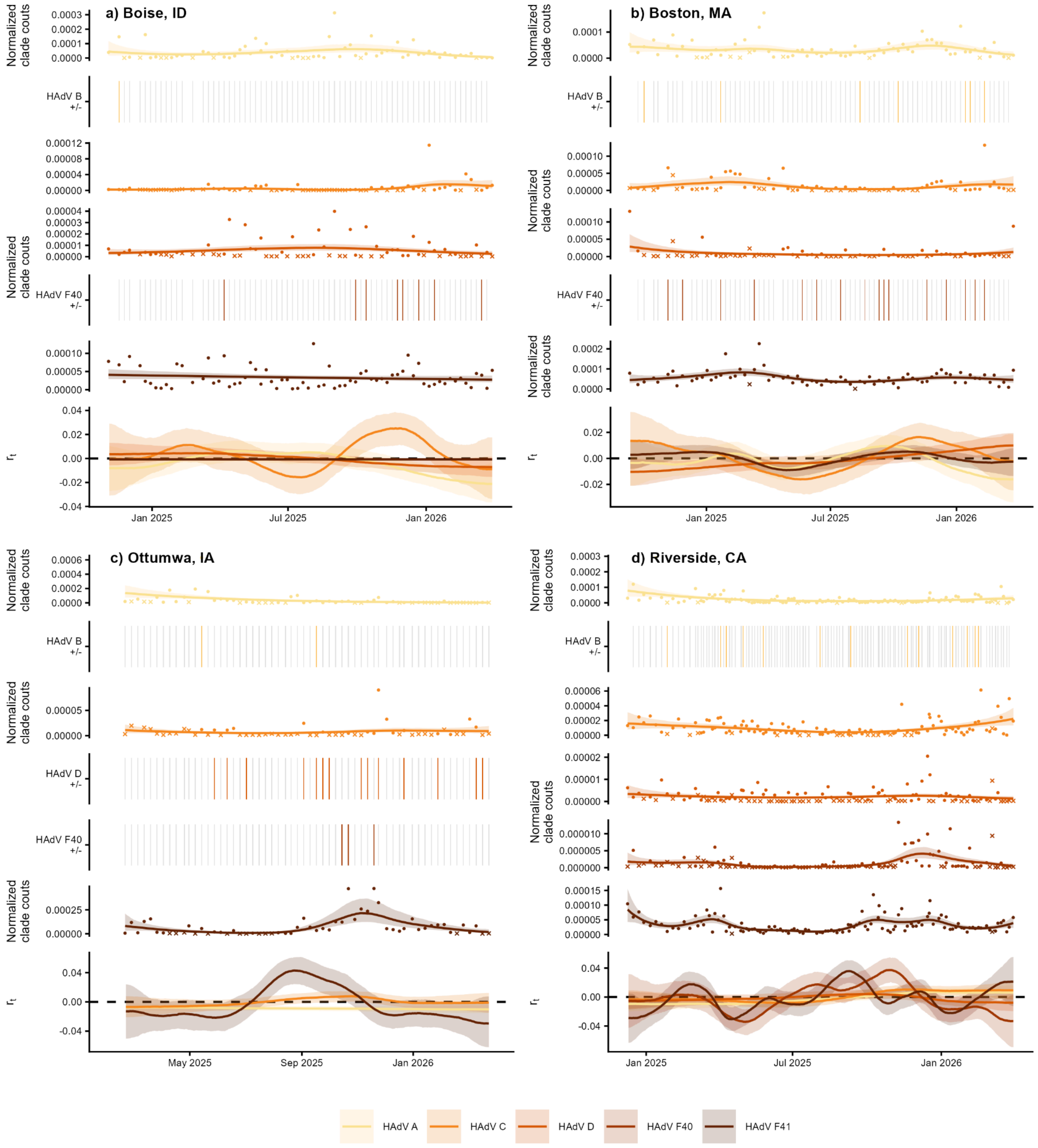

**Fig. S20. Untargeted HAdV wastewater monitoring data and $r_t$ estimates for all locations, differentiated by species/type.**

Untargeted wastewater monitoring time series data with GAM smoothing alongside $r_t$ estimates are shown for (a) Boise, Idaho; (b) Boston, Massachusetts; (c) Ottumwa, Iowa; and (d) Riverside, California. HAdV clade counts are normalized by PMMoV clade counts and scaled to account for sequencing depth; x's denote non-detect samples and outlier measurements are not shown for visualization. Lines represent median estimates with 90% CrIs shaded. Heatmaps showing presence/absence are used when >75% of samples did not result in a positive virus detection. Abbreviations: CrI = credible interval, GAM = generalized additive model, HAdV = human adenovirus, PMMoV = pepper mild mottle virus.

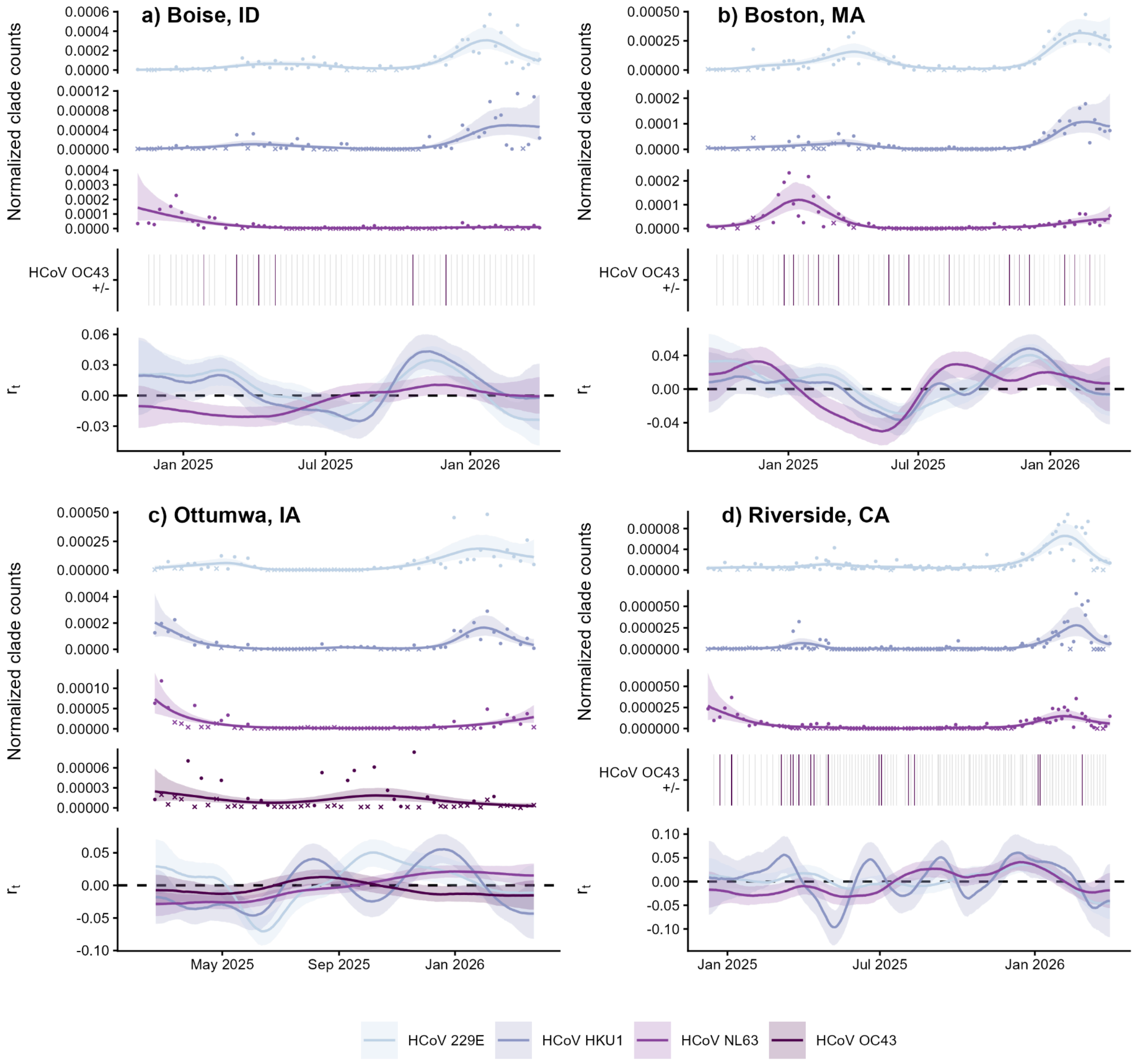

**Fig. S21. Untargeted HCoV wastewater monitoring data and $r_t$ estimates for all locations, differentiated by species.**

Untargeted wastewater monitoring time series data with GAM smoothing alongside $r_t$ estimates are shown for (a) Boise, Idaho; (b) Boston, Massachusetts; (c) Ottumwa, Iowa; and (d) Riverside, California. HCoV clade counts are normalized by PMMoV clade counts and scaled to account for sequencing depth; x's denote non-detect samples and outlier measurements are not shown for visualization. Lines represent median estimates with 90% CrIs shaded. Heatmaps showing presence/absence are used when >75% of samples did not result in a positive virus detection. Abbreviations: CrI = credible interval, GAM = generalized additive model, HCoV = human coronavirus, PMMoV = pepper mild mottle virus.

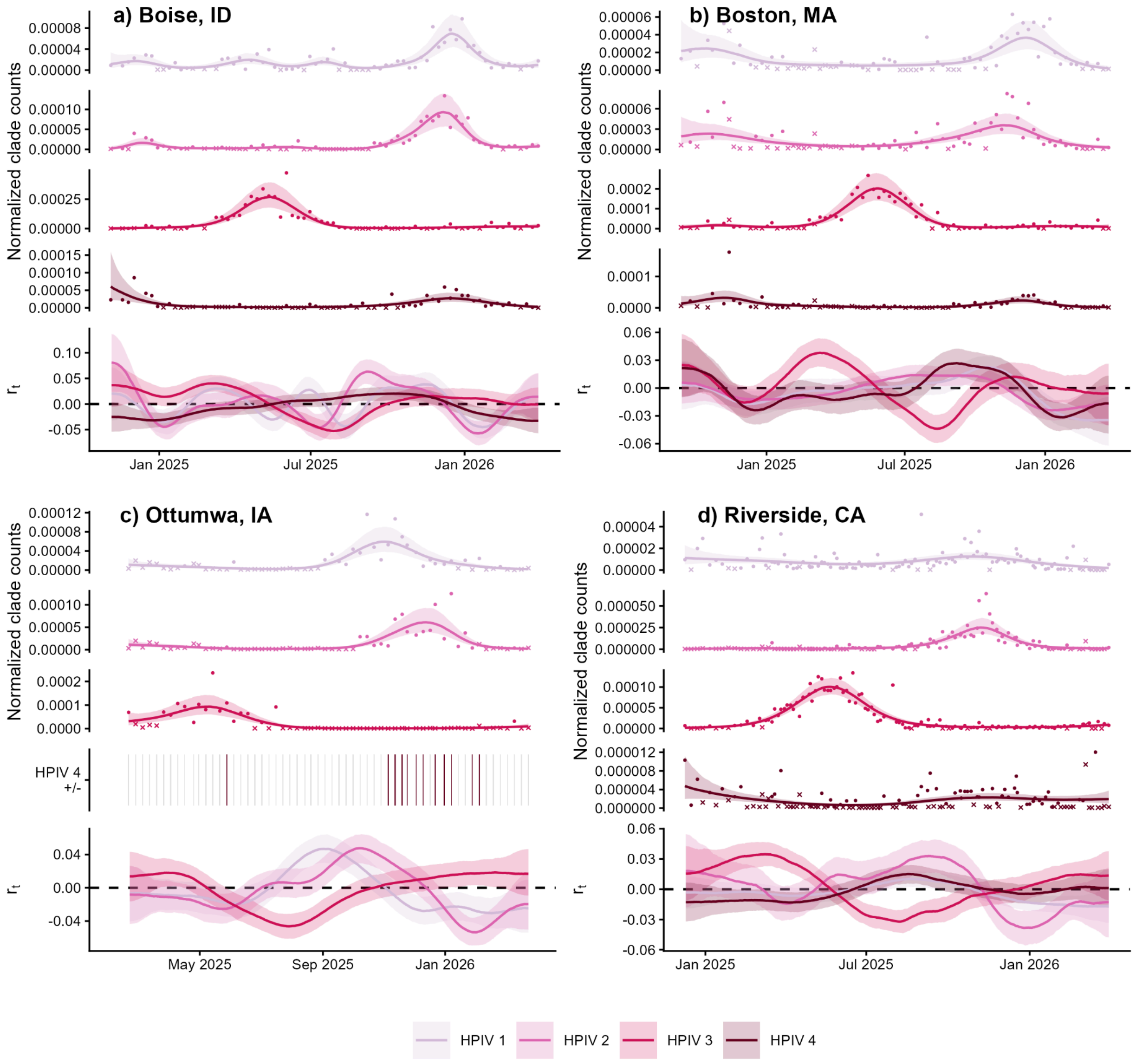

**Fig. S22. Untargeted HPIV wastewater monitoring data and $r_t$ estimates for all locations, differentiated by type.**

Untargeted wastewater monitoring time series data with GAM smoothing alongside $r_t$ estimates are shown for (a) Boise, Idaho; (b) Boston, Massachusetts; (c) Ottumwa, Iowa; and (d) Riverside, California. HPIV clade counts are normalized by PMMoV clade counts and scaled to account for sequencing depth; x's denote non-detect samples and outlier measurements are not shown for visualization. Lines represent median estimates with 90% CrIs shaded. Heatmaps showing presence/absence are used when >75% of samples did not result in a positive virus detection. Abbreviations: CrI = credible interval, GAM = generalized additive model, HPIV = human parainfluenza virus, PMMoV = pepper mild mottle virus.

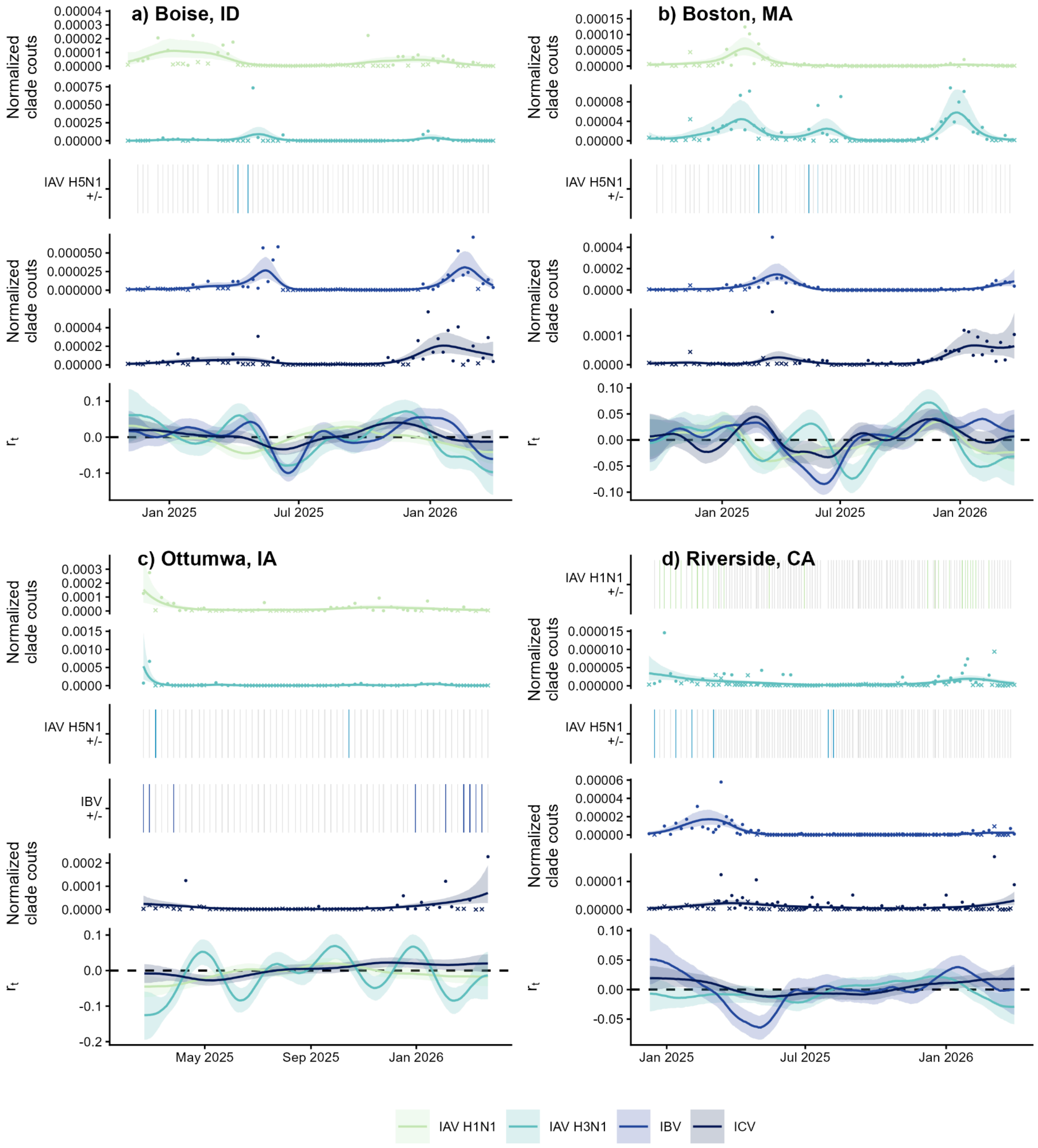

**Fig. S23. Untargeted influenza wastewater monitoring data and $r_t$ estimates for all locations, differentiated by type/subtype/lineage.**

Untargeted wastewater monitoring time series data with GAM smoothing alongside $r_t$ estimates are shown for (a) Boise, Idaho; (b) Boston, Massachusetts; (c) Ottumwa, Iowa; and (d) Riverside, California. Influenza clade counts are normalized by PMMoV clade counts and scaled to account for sequencing depth; x's denote non-detect samples and outlier measurements are not shown for visualization. Lines represent median estimates with 90% CrIs shaded. Heatmaps showing presence/absence are used when >75% of samples did not result in a positive virus detection. Abbreviations: CrI = credible interval, GAM = generalized additive model, IAV = influenza A virus, IBV = influenza B virus, ICV = influenza C virus, PMMoV = pepper mild mottle virus.

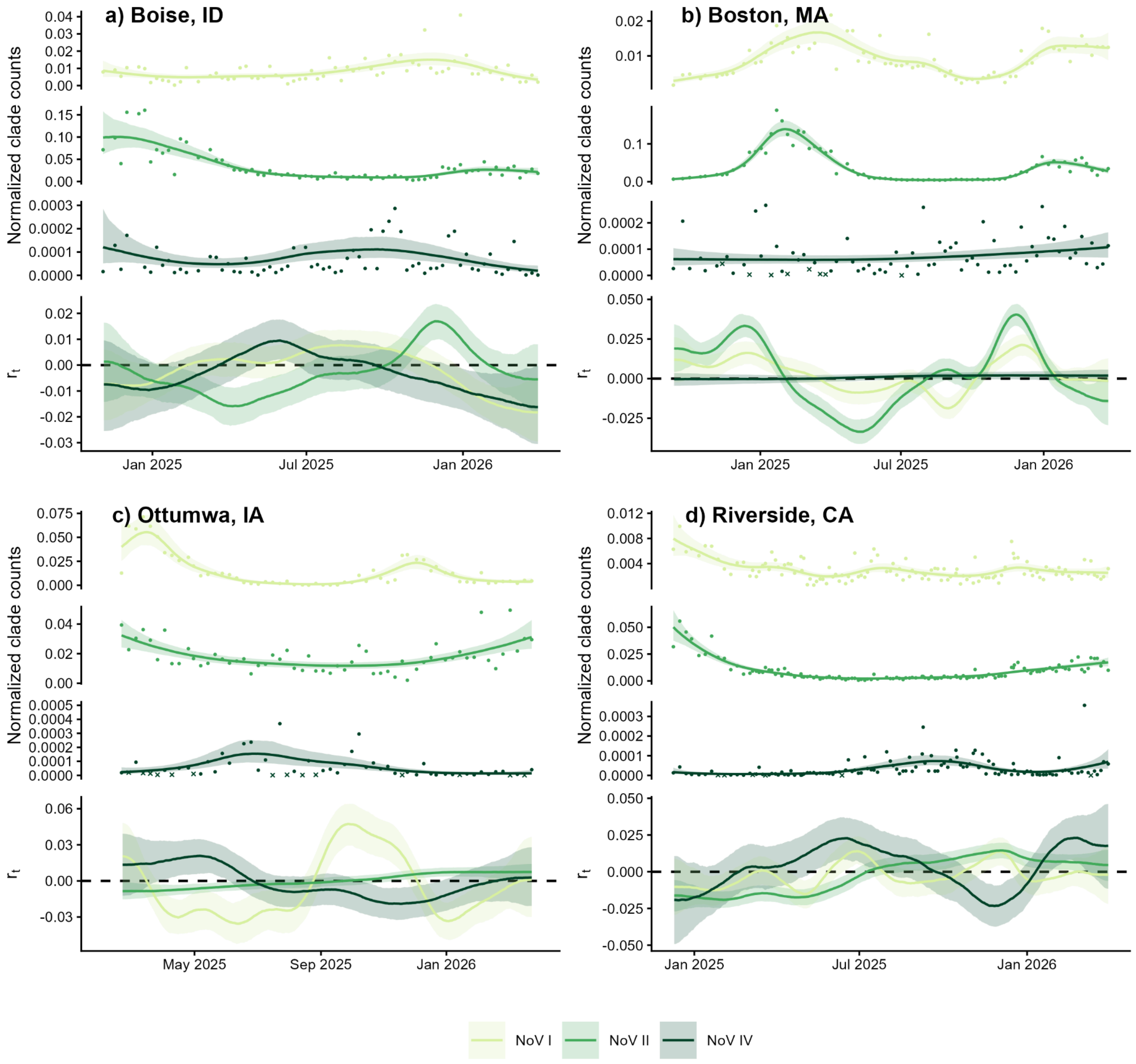

**Fig. S24. Untargeted norovirus wastewater monitoring data and $r_t$ estimates for all locations, differentiated by genogroup.**

Untargeted wastewater monitoring time series data with GAM smoothing alongside $r_t$ estimates are shown for (a) Boise, Idaho; (b) Boston, Massachusetts; (c) Ottumwa, Iowa; and (d) Riverside, California. NoV clade counts are normalized by PMMoV clade counts and scaled to account for sequencing depth; x's denote non-detect samples and outlier measurements are not shown for visualization. Lines represent median estimates with 90% CrIs shaded. Abbreviations: CrI = credible interval, GAM = generalized additive model, NoV = norovirus, PMMoV = pepper mild mottle virus.

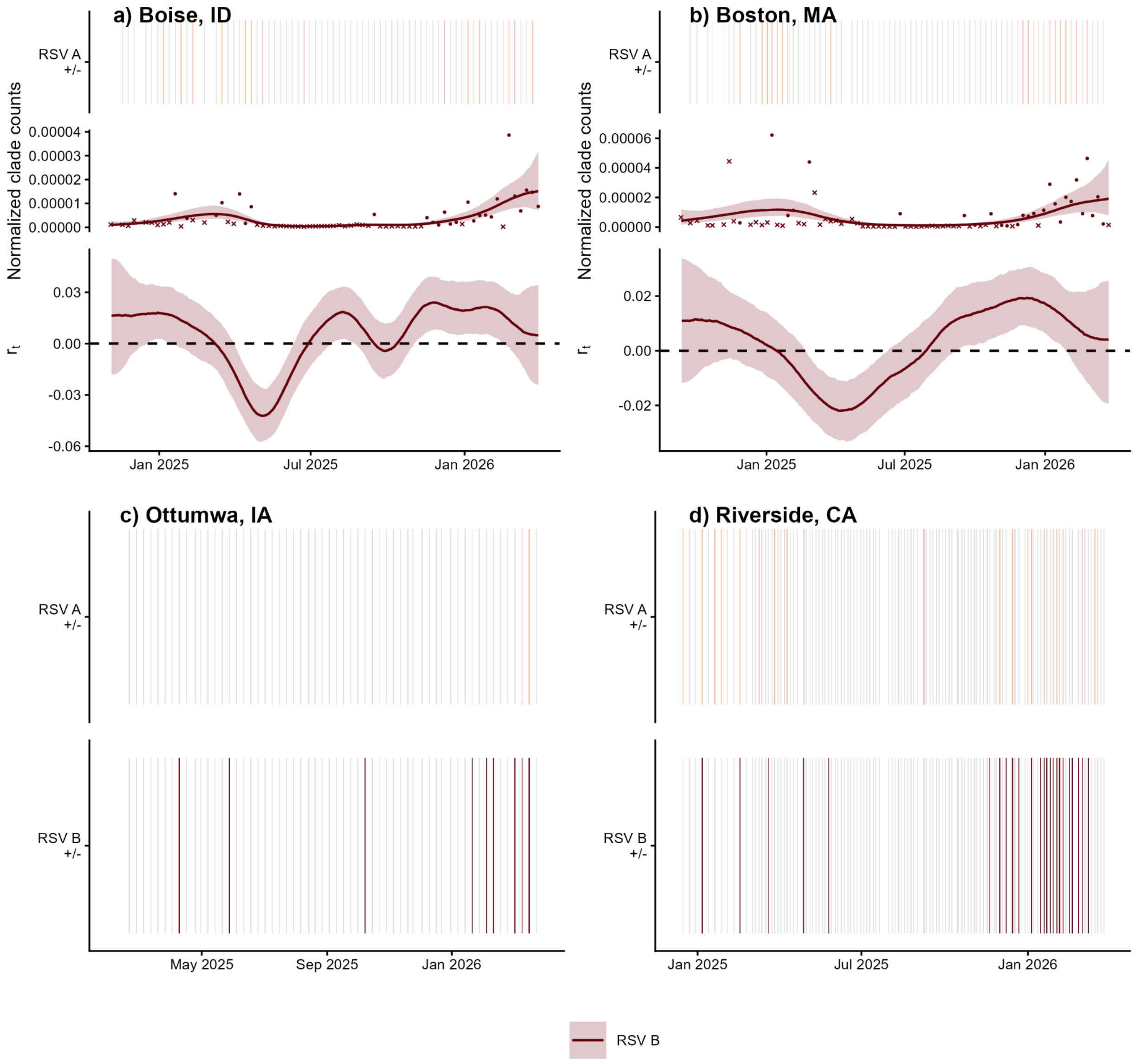

**Fig. S25. Untargeted RSV wastewater monitoring data and $r_t$ estimates for all locations, differentiated by group.**

Untargeted wastewater monitoring time series data with GAM smoothing alongside $r_t$ estimates are shown for (a) Boise, Idaho; (b) Boston, Massachusetts; (c) Ottumwa, Iowa; and (d) Riverside, California. RSV clade counts are normalized by PMMoV clade counts and scaled to account for sequencing depth; x's denote non-detect samples and outlier measurements are not shown for visualization. Lines represent median estimates with 90% CrIs shaded. Heatmaps showing presence/absence are used when >75% of samples did not result in a positive virus detection. Abbreviations: CrI = credible interval, GAM = generalized additive model, PMMoV = pepper mild mottle virus, RSV = respiratory syncytial virus.

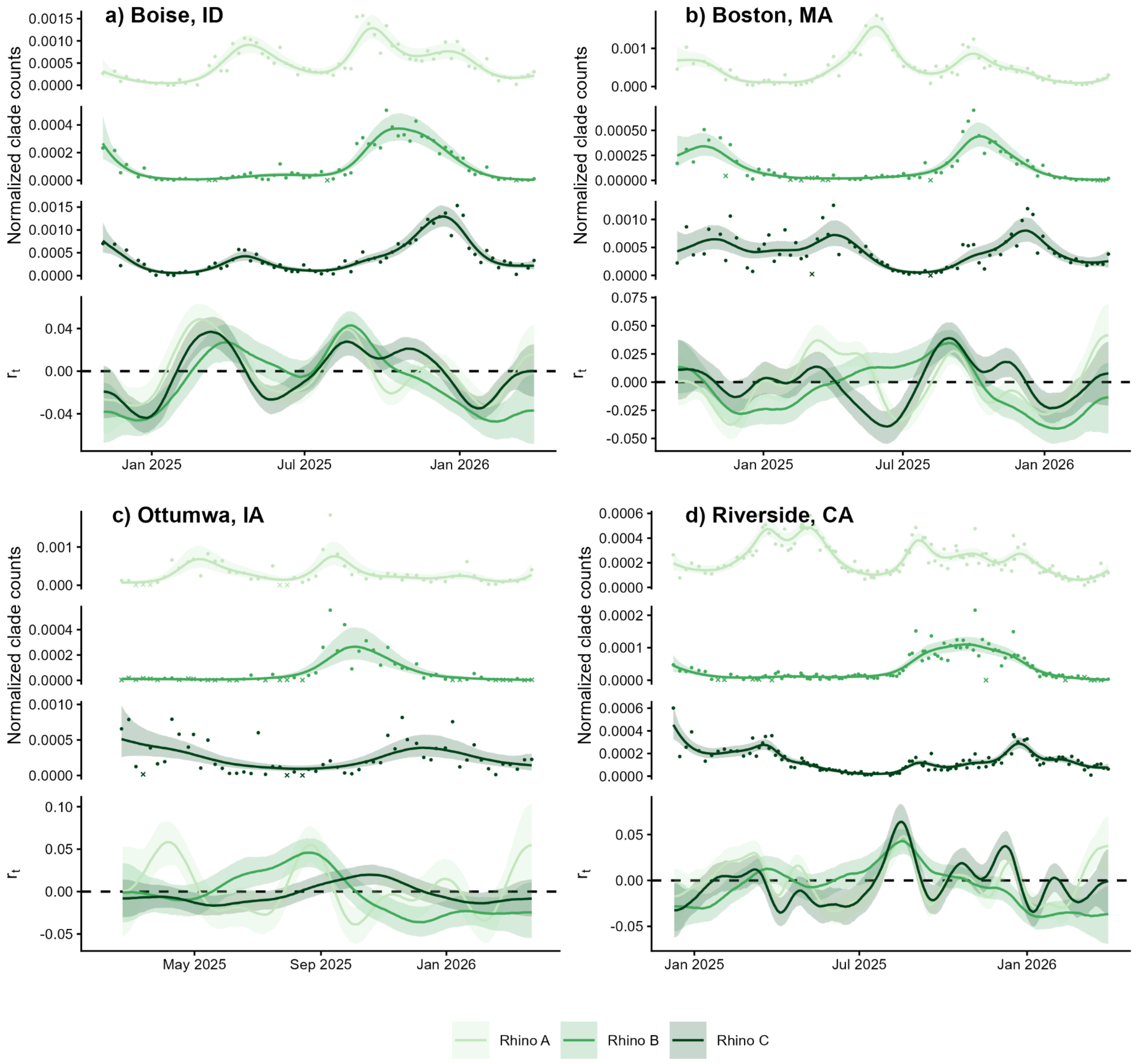

**Fig. S26. Untargeted rhinovirus wastewater monitoring data and $r_t$ estimates for all locations, differentiated by species.**

Untargeted wastewater monitoring time series data with GAM smoothing alongside $r_t$ estimates are shown for (a) Boise, Idaho; (b) Boston, Massachusetts; (c) Ottumwa, Iowa; and (d) Riverside, California. Rhinovirus ("Rhino") clade counts are normalized by PMMoV clade counts and scaled to account for sequencing depth; x's denote non-detect samples and outlier measurements are not shown for visualization. Lines represent median estimates with 90% CrIs shaded. Abbreviations: CrI = credible interval, GAM = generalized additive model, PMMoV = pepper mild mottle virus.

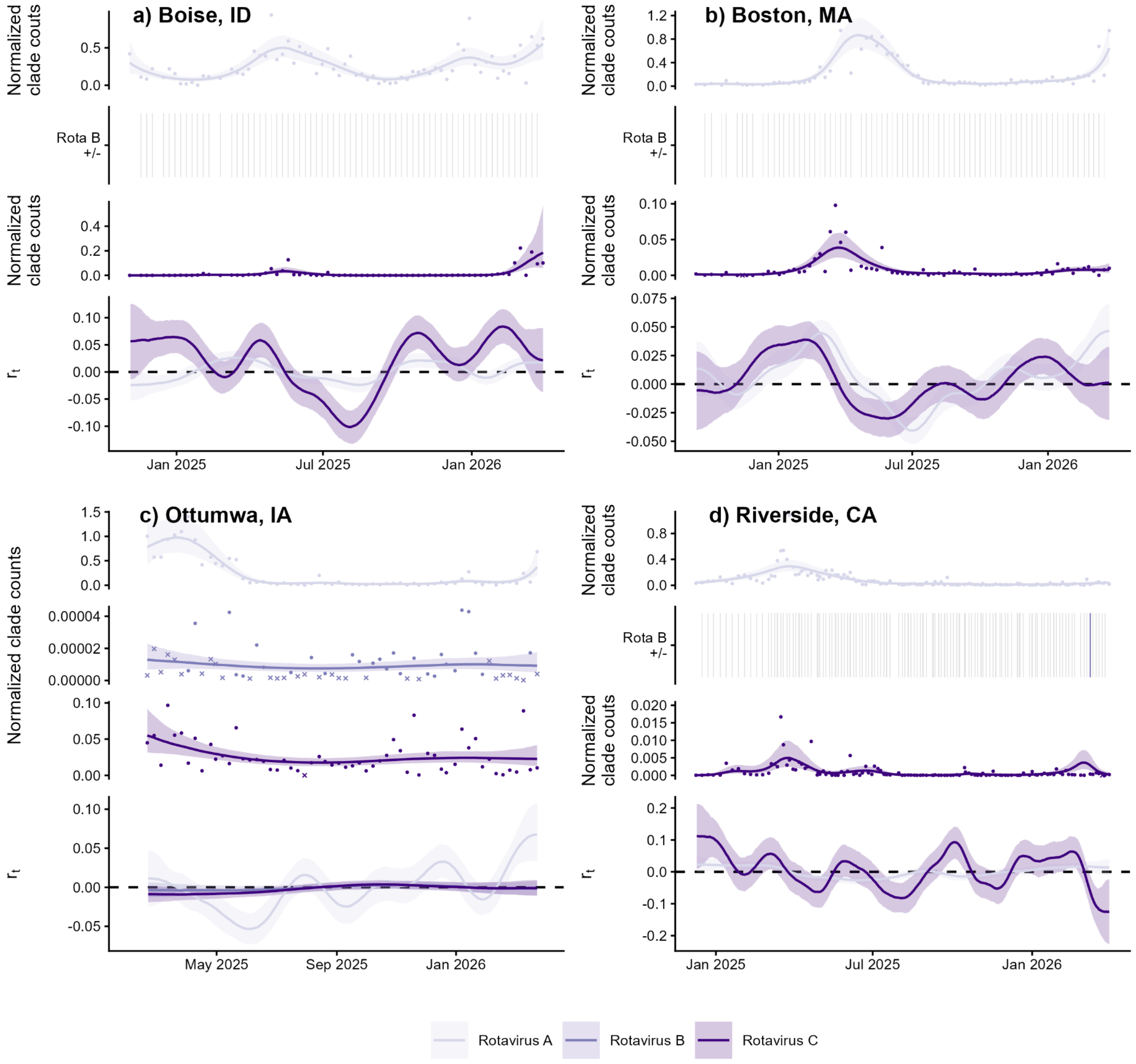

**Fig. S27. Untargeted rotavirus wastewater monitoring data and $r_t$ estimates for all locations, differentiated by group.**

Untargeted wastewater monitoring time series data with GAM smoothing alongside $r_t$ estimates are shown for (a) Boise, Idaho; (b) Boston, Massachusetts; (c) Ottumwa, Iowa; and (d) Riverside, California. Rotavirus clade counts are normalized by PMMoV clade counts and scaled to account for sequencing depth; x's denote non-detect samples and outlier measurements are not shown for visualization. Lines represent median estimates with 90% CrIs shaded. Heatmaps showing presence/absence are used when >75% of samples did not result in a positive virus detection. Abbreviations: CrI = credible interval, GAM = generalized additive model, PMMoV = pepper mild mottle virus.

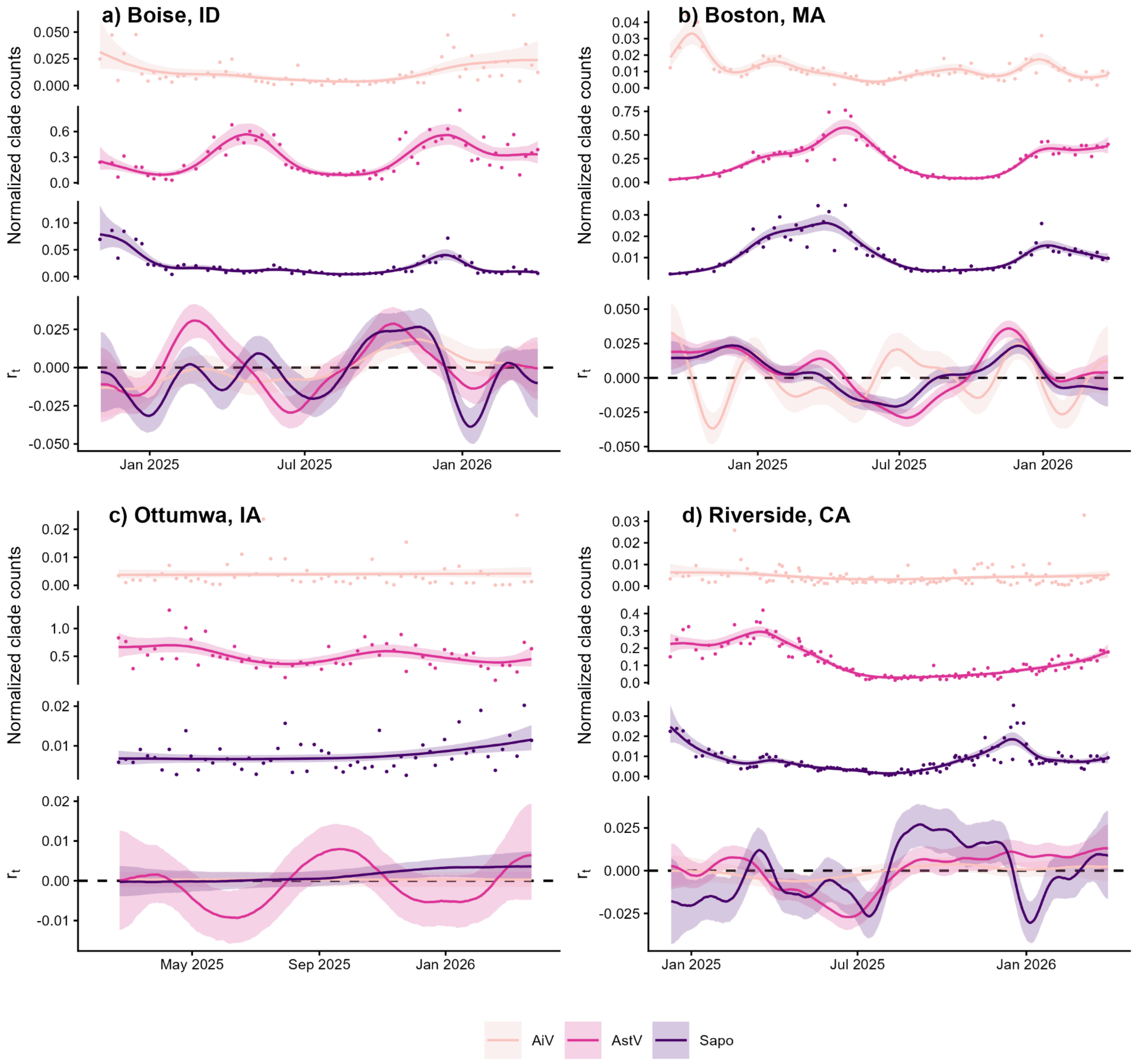

**Fig. S28. Untargeted AiV, AstV, and sapovirus wastewater monitoring data and $r_t$ estimates for all locations.**

Untargeted wastewater monitoring time series data with GAM smoothing alongside $r_t$ estimates are shown for (a) Boise, Idaho; (b) Boston, Massachusetts; (c) Ottumwa, Iowa; and (d) Riverside, California. Other enteric virus clade counts are normalized by PMMoV clade counts and scaled to account for sequencing depth; x's denote non-detect samples and outlier measurements are not shown for visualization. Lines represent median estimates with 90% CrIs shaded. Abbreviations: AiV = aichivirus, AstV = astrovirus, CrI = credible interval, GAM = generalized additive model, PMMoV = pepper mild mottle virus, sapo = sapovirus.

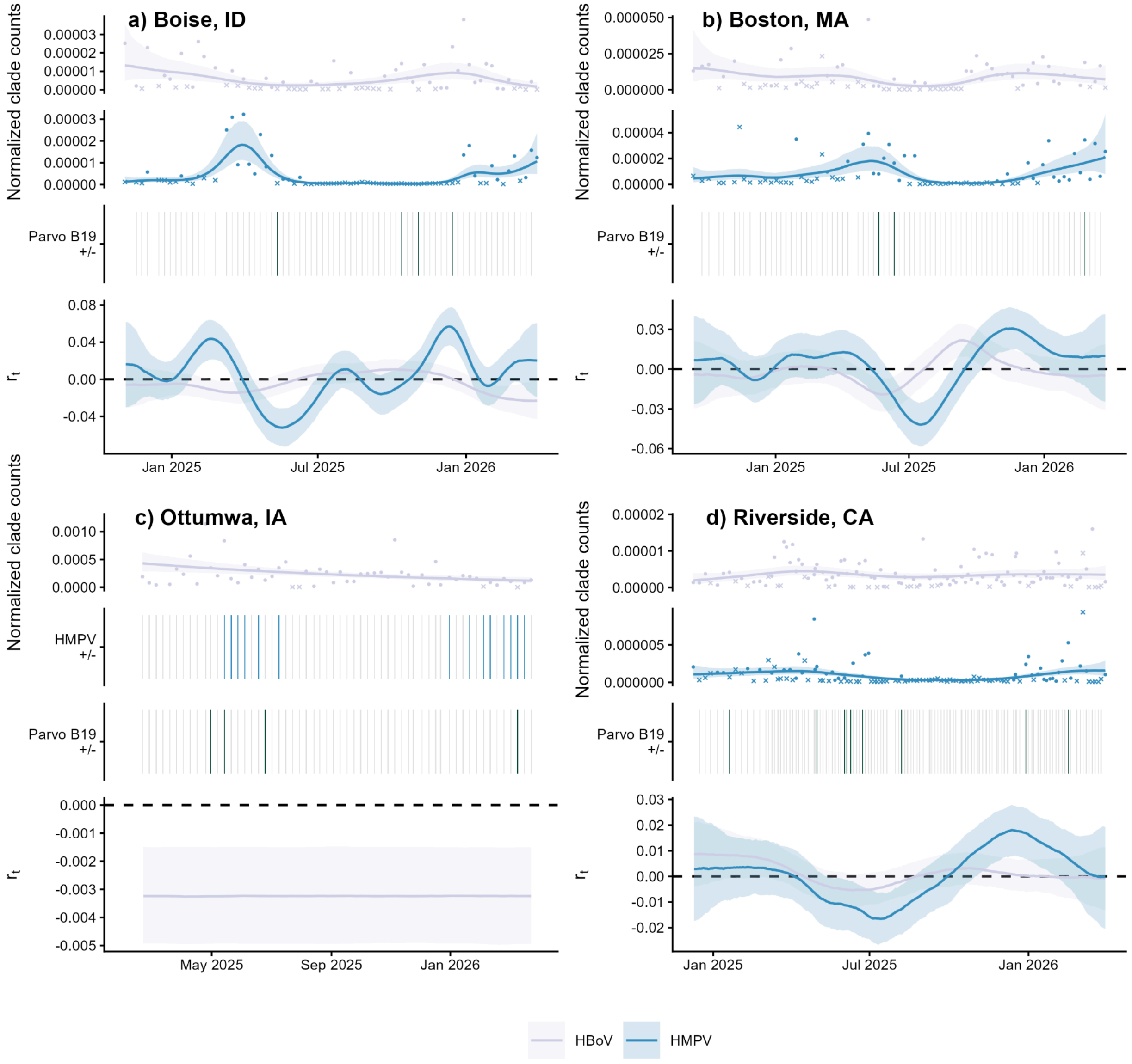

**Fig. S29. Untargeted HBoV, HMPV, and parvovirus B19 wastewater monitoring data and $r_t$ estimates for all locations.**

Untargeted wastewater monitoring time series data with GAM smoothing alongside $r_t$ estimates are shown for (a) Boise, Idaho; (b) Boston, Massachusetts; (c) Ottumwa, Iowa; and (d) Riverside, California. Other respiratory virus clade counts are normalized by PMMoV clade counts and scaled to account for sequencing depth; x's denote non-detect samples and outlier measurements are not shown for visualization. Lines represent median estimates with 90% CrIs shaded. Heatmaps showing presence/absence are used when >75% of samples did not result in a positive virus detection. Abbreviations: CrI = credible interval, GAM = generalized additive model, HBoV = human bocavirus, HMPV = human metapneumovirus, PMMoV = pepper mild mottle virus.

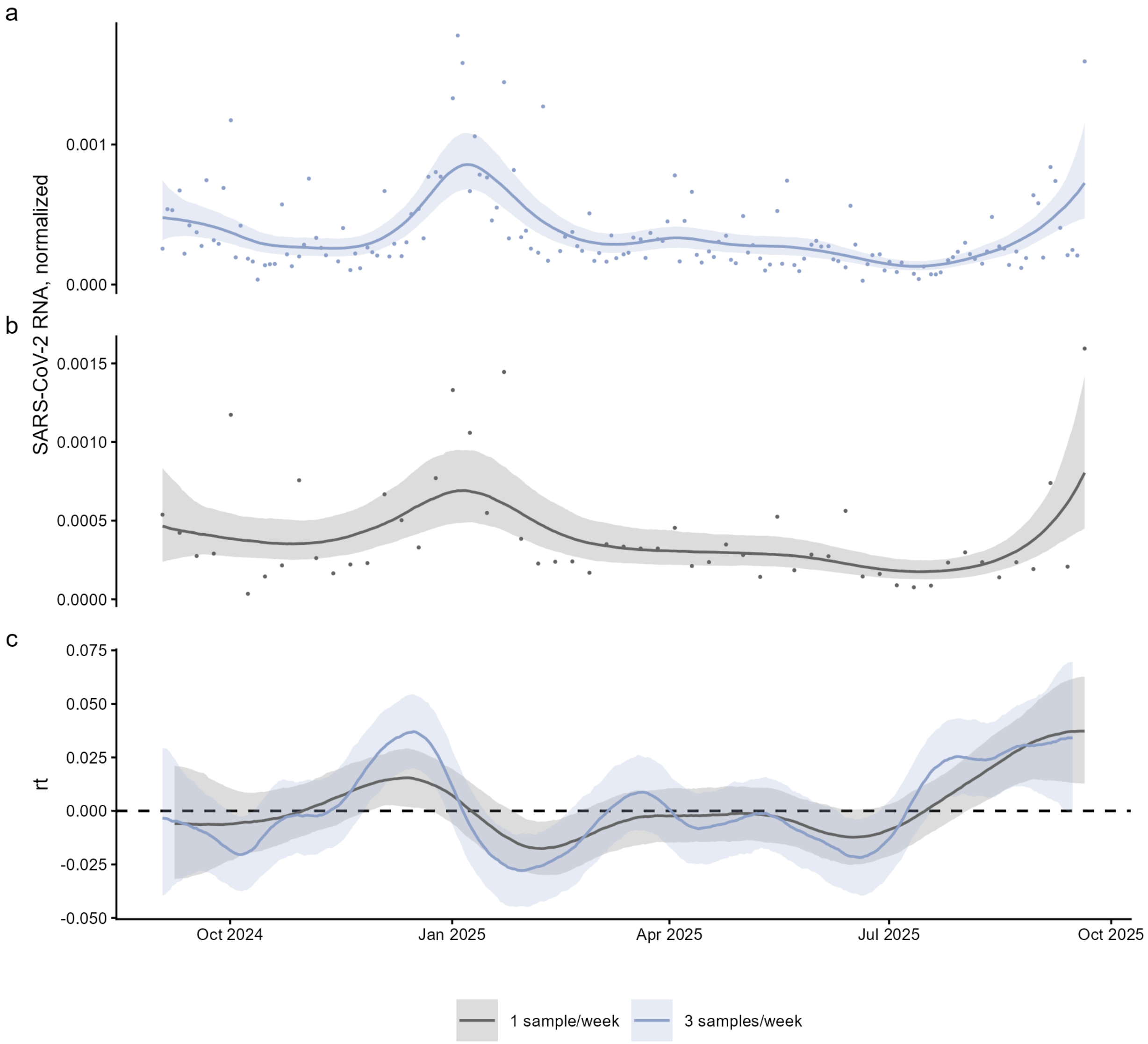

**Fig. S30. Example of $r_t$ estimates using different wastewater sampling frequencies.**
Targeted SARS-CoV-2 wastewater monitoring data for Boston, MA with GAM smoothing at (**a**) 3 samples per week (purple) and (**b**) 1 sample/week (gray). SARS-CoV-2 RNA concentrations are normalized by PMMoV RNA concentrations; outlier measurements are not shown for visualization. Downsampled data in **b** represent one random sample each week from **a**. (**c**) Derived $r_t$ estimates from each wastewater sampling frequency. Lines represent median estimates with 90% CrIs shaded. Abbreviations: CrI = credible interval, GAM = generalized additive model, PMMoV = pepper mild mottle virus.

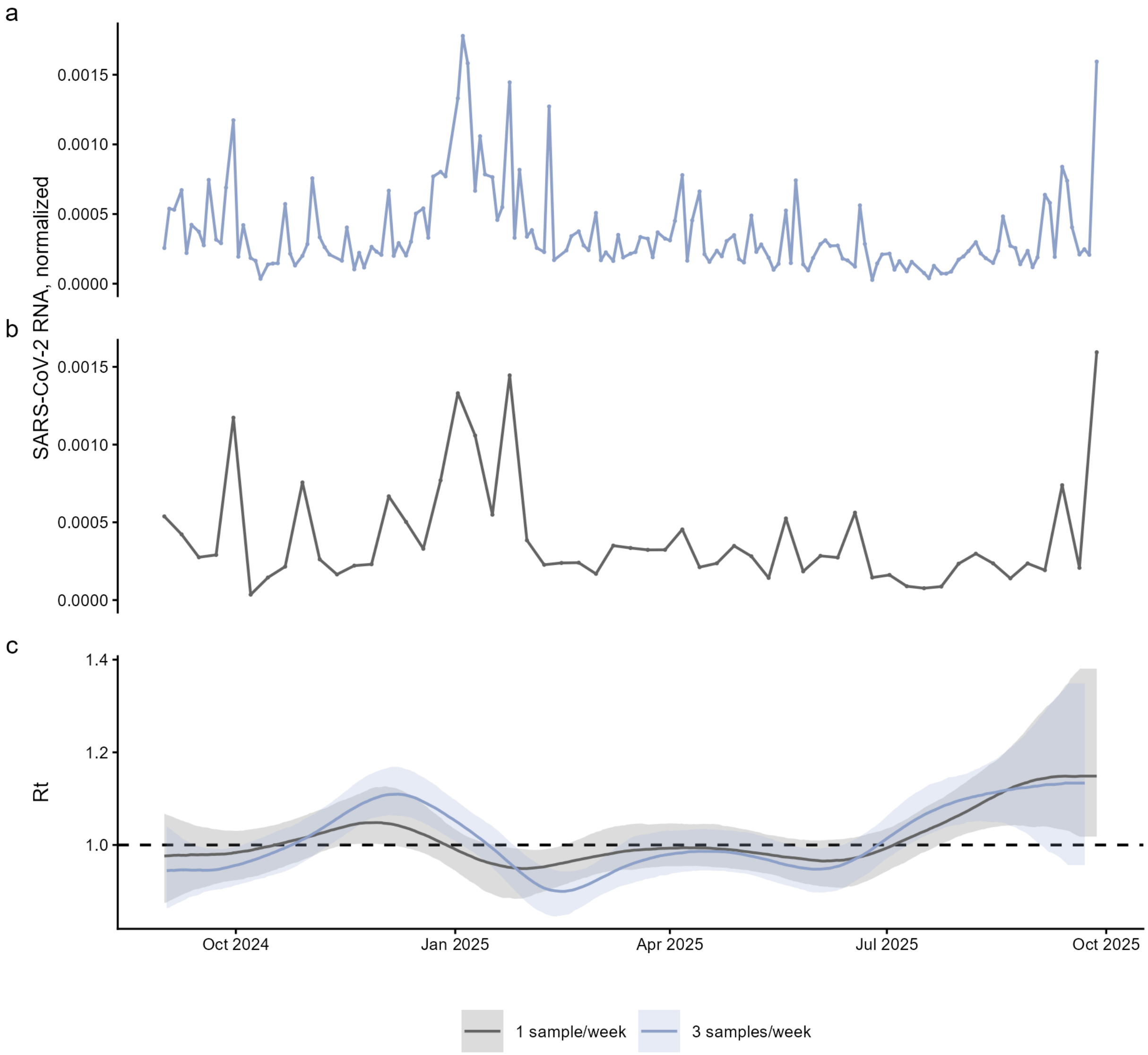

**Fig. S31. Example of $R_t$ estimates using different wastewater sampling frequencies.**
Targeted SARS-CoV-2 wastewater monitoring data for Boston, MA at (**a**) 3 samples per week (purple) and (**b**) 1 sample/week (gray). SARS-CoV-2 RNA concentrations are normalized by PMMoV RNA concentrations; outlier measurements are not shown for visualization. Downsampled data in **b** represent one random sample each week from **a**. (**c**) Derived $R_t$ estimates from each wastewater sampling frequency. Lines represent median estimates with 90% CrIs shaded. Abbreviations: CrI = credible interval, GAM = generalized additive model, PMMoV = pepper mild mottle virus.

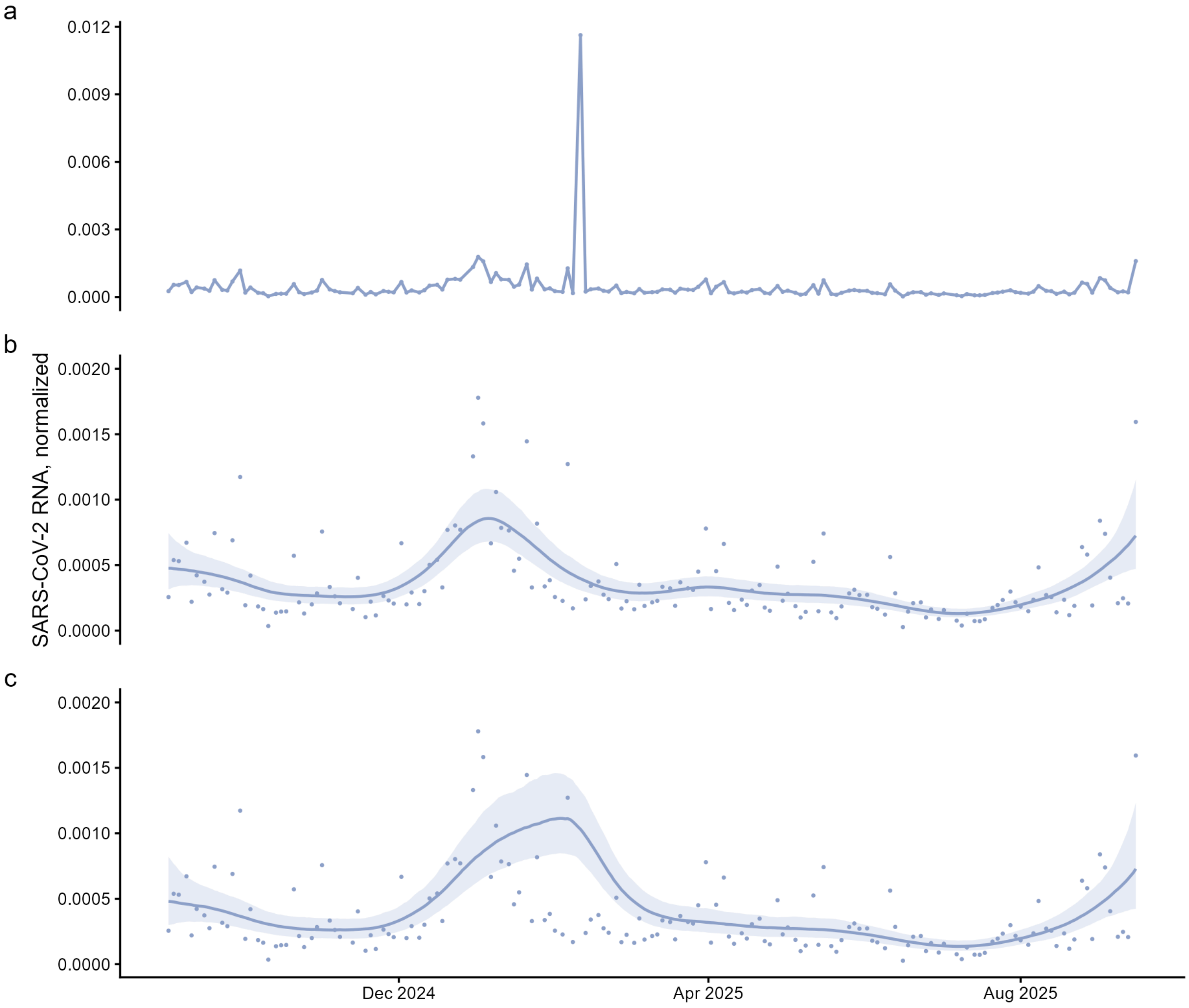

**Fig. S32. Example of GAM smoothing with and without outliers.**
(**a**) Targeted wastewater monitoring data (SARS-CoV-2 RNA concentrations normalized by PMMoV RNA concentrations) for Boston, Massachusetts. The normalized concentration above 0.003 is an outlier sample. Corresponding GAM-smoothed time series data without (**b**) and with (**c**) the outlier sample retained. Solid lines represent the median and shaded regions represent the 90% CrI. The y-axis in (**b**) and (**c**) is adjusted such that the outlier sample is not visible. Abbreviations: CrI = credible interval, GAM = generalized additive model, PMMoV = pepper mild mottle virus.

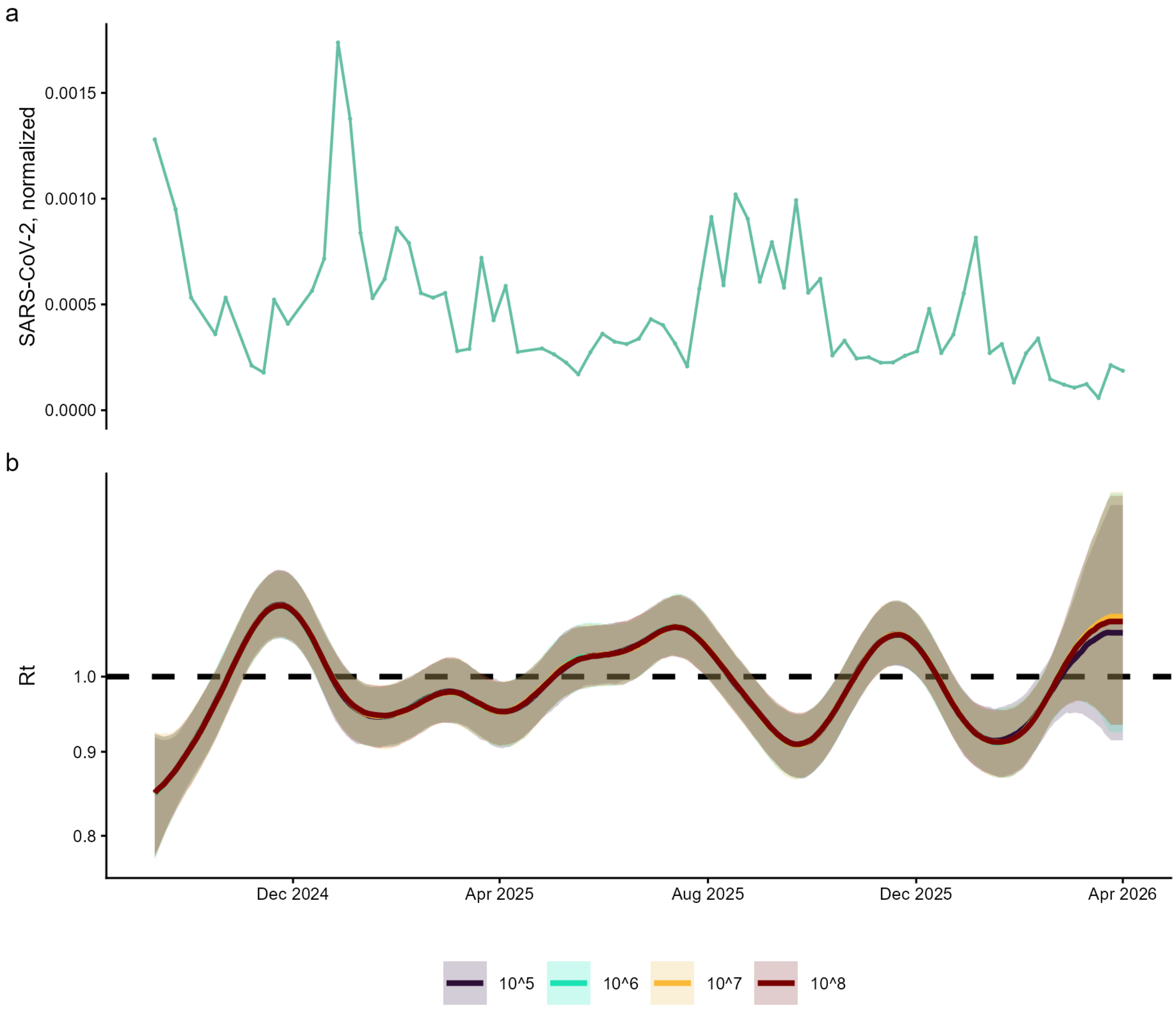

**Fig. S33. Example of $R_t$ estimations using different multiplication factors.**

(**a**) Untargeted wastewater monitoring data (SARS-CoV-2 clade counts normalized by PMMoV clade counts and scaled to account for sequencing depth) for Boston, Massachusetts. (**b**) Corresponding $R_t$ estimations applying various multiplication factors to the untargeted wastewater monitoring data ($10^5$ = navy blue, $10^6$ = teal, $10^7$ = yellow, $10^8$ = red). Solid lines represent median estimates with 90% CrIs shaded. Abbreviations: CrI = credible interval, GAM = generalized additive model, PMMoV = pepper mild mottle virus.
